# Predicting diagnostic progression to schizophrenia or bipolar disorder via machine learning applied to electronic health record data

**DOI:** 10.1101/2024.07.02.24309828

**Authors:** Lasse Hansen, Martin Bernstorff, Kenneth Enevoldsen, Sara Kolding, Jakob Grøhn Damgaard, Erik Perfalk, Kristoffer L. Nielbo, Andreas A. Danielsen, Søren D. Østergaard

## Abstract

**Importance:** The diagnosis of schizophrenia and bipolar disorder is often delayed several years despite illness typically emerging in late adolescence or early adulthood, which impedes initiation of targeted treatment.

**Objective:** To investigate whether machine learning models trained on routine clinical data from electronic health records (EHRs) can predict diagnostic progression to schizophrenia or bipolar disorder among patients undergoing treatment in psychiatric services for other mental illness.

**Design:** Cohort study based on data from EHRs.

**Setting:** The psychiatric services of the Central Denmark Region.

**Participants:** All patients between ≥15 and <60 years with at least one contact with the psychiatric services of the Central Denmark Region between 2011 and 2021. Patients with only a single contact were removed, leaving a total of 24,449 eligible patients with 398,922 outpatient contacts with the psychiatric services.

**Exposures:** Predictors based on EHR data, including medications, diagnoses, and clinical notes.

**Main Outcomes and Measures:** Diagnostic transition to schizophrenia or bipolar disorder within 5 years, predicted one day before outpatient contacts by means of regularized logistic regression and Extreme Gradient Boosting (XGBoost) models.

**Results:** Transition to the first occurrence of either schizophrenia or bipolar disorder was predicted by the XGBoost model with an area under the receiver operating characteristics curve (AUROC) of 0.70 on the training set, and 0.64 on the test set which consisted of two held-out hospital sites. At a predicted positive rate of 4%, the XGBoost model had a sensitivity of 9.3%, a specificity of 96.3%, and a positive predictive value of 13.0%. Predicting schizophrenia and bipolar disorder separately yielded AUROCs of 0.80 and 0.62, respectively, on the test set.

The clinical notes proved particularly informative for prediction.

**Conclusions and relevance:** It is possible to predict diagnostic transition to schizophrenia and bipolar disorder from routine clinical data extracted from EHRs, with schizophrenia being notably easier to predict than bipolar disorder.

**Key Points:** *Question:* Can diagnostic progression to schizophrenia or bipolar disorder be accurately predicted from routine clinical data extracted from electronic health records?

*Findings:* In this study, which included all patients aged between ≥15 and <60 years with contacts to the psychiatric services of the Central Denmark Region between 2011 and 2021, progression to schizophrenia was predicted with high accuracy, with bipolar disorder proving a more difficult target.

*Meaning:* **Detecting p**rogression to schizophrenia through machine learning based on routine clinical data is feasible. This may reduce diagnostic delay and duration of untreated illness.

## Introduction

Schizophrenia and bipolar disorder are severe mental disorders that often impair the ability to lead a normal life (1,2). Indeed, both disorders have severe negative consequences for social functioning, work ability, and lifespan (1–3). Despite typically emerging in late adolescence or early adulthood, diagnosis is often delayed several years (4,5). Timely and accurate diagnosis is crucial, as diagnostic delay impedes the initiation of targeted treatment.

Furthermore, the longer the duration of untreated illness, the worse the prognosis becomes (4,5). However, timely diagnosis of schizophrenia and bipolar disorder is challenging due to the prodromal phase, in which patients do not yet meet full diagnostic criteria, and due to symptom overlap with other disorders such as anxiety and depression (1,6). In fact, many patients who are eventually diagnosed with schizophrenia and bipolar disorder have previously received treatment for other and less severe mental disorders (7,8).

Machine learning applied to electronic health record (EHR) data likely holds great promise for assisting in the diagnosis of complex psychiatric conditions such as schizophrenia and bipolar disorder (9). Clinical notes in EHRs are presumably particularly valuable in this context, as they contain comprehensive descriptions of symptoms, treatment responses, and patient-clinician interactions. Due to the sheer amount of unstructured text in these notes, often covering several years, it is difficult for clinicians to harness and utilize the comprehensive information embedded within them efficiently. Using methods from natural language processing (NLP) and deep learning, it may be possible to extract and synthesize data from clinical notes, uncovering patterns that could indicate an impending progression from less severe conditions to schizophrenia or bipolar disorder (10).

This paper investigates whether machine learning models trained on routine clinical data from electronic health records can predict the risk of diagnostic progression to schizophrenia or bipolar disorder among patients undergoing treatment in psychiatric services. Early diagnosis enabled by machine learning models could potentially reduce the duration of untreated illness in schizophrenia and bipolar disorder, leading to better prognoses and improved illness trajectories.

## Methods

Reporting follows the guidelines set out in Transparent Reporting of multivariable prediction models for Individual Prognosis Or Diagnosis with Artificial Intelligence (TRIPOD+AI) (11).

### Data

The study included data from an updated version of the PSYchiatric Clinical Outcome Prediction (PSYCOP) cohort (12). The cohort contains routine electronic health record data from all individuals with at least one contact with the Psychiatric Services of the Central Denmark Region (catchment population of approximately 1.3 million) in the period from January 1, 2011, to November 22, 2021 (Figure 1A). The data covers all contacts with public hospitals in the Central Denmark Region (both psychiatric and somatic hospitals). As the Danish healthcare system is universal, public hospitals financially cover the vast majority of hospital contacts.

**Figure 1:**
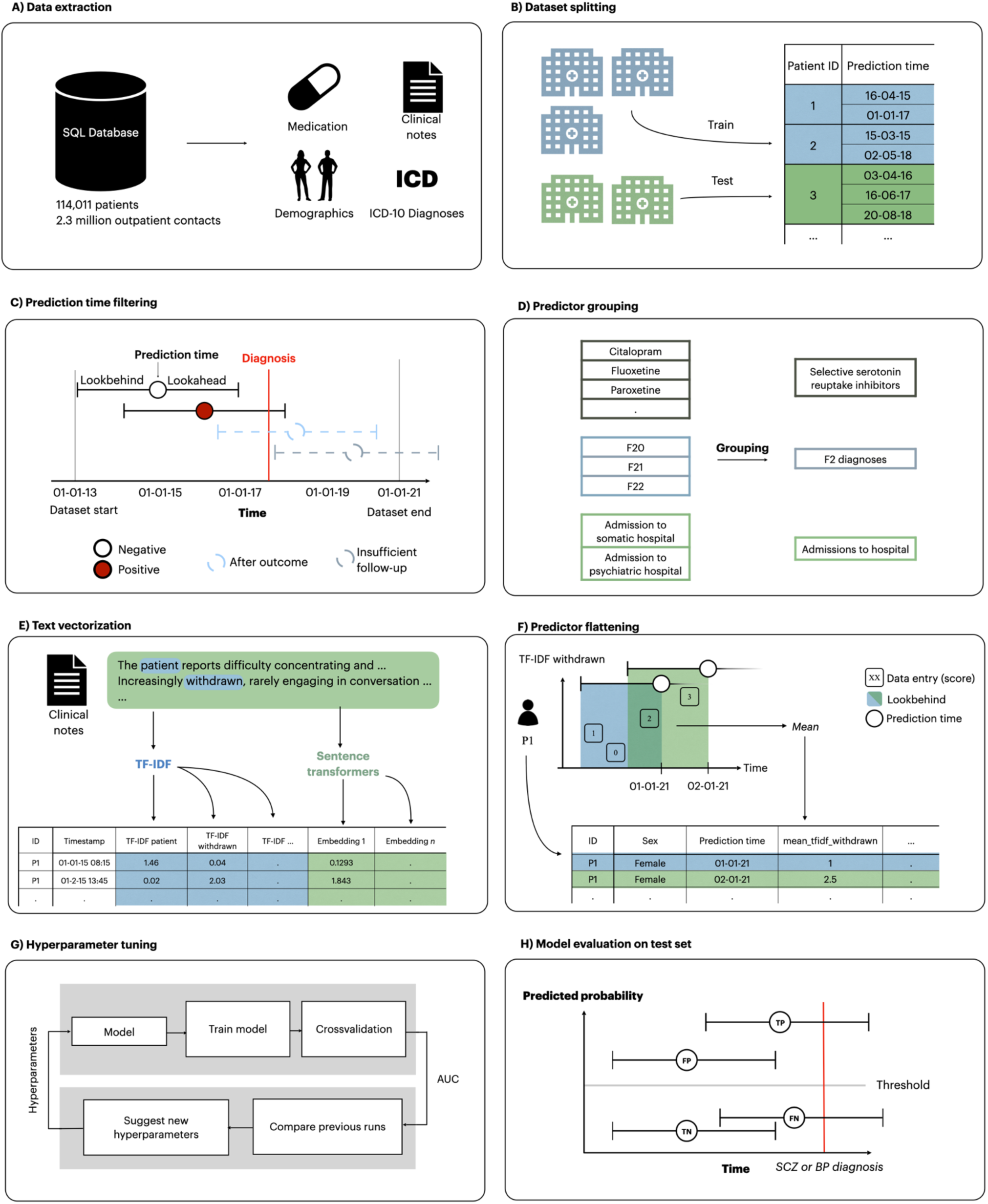
Data extraction and transformation, model training, and model testing pipeline. A) Data was extracted from the electronic health records. B) Data was split into a training and a test set based on hospital sites. C) Prediction times occurring after November 21, 2016 were removed due to lack of follow-up. Prediction times occurring after a diagnosis of schizophrenia or bipolar disorder were also removed. D) Certain predictors were grouped. E) Clinical notes were turned into vectors using TF-IDF or sentence transformer models. F) Predictors for each prediction time were extracted by aggregating the variables within the lookbehind with an aggregation function. As a result, each row in the dataset represents a specific prediction time with a column for each predictor. G) Models were trained and optimised on the training set using 5-fold cross-validation. Hyperparameters were tuned to optimise AUROC. H) The best candidate models were evaluated on the test set. Figure adapted from (34).

### Data Split

As the data in the PSYCOP cohort spans five psychiatric hospitals, we split the data into a training and a test set based on geographical location, in order to assess the external validity of the models. Specifically, patient contacts with the hospitals in the western and eastern part of the region (Aarhus, Herning, Holstebro, Randers, Horsens, and Gødstrup) were used for training, while patient contacts with the central part of the region (Silkeborg and Viborg) were used for testing (Fig 1B). As patients might first receive treatment in one of the geographical splits and then move or be transferred to the other split, we dropped prediction times occurring after a move to avoid having the same patient present in both splits, as this might introduce leakage (13).

### Cohort Definition

The cohort was limited to contacts occurring after January 1, 2013, due to inconsistencies in the data before 2013, stemming from the gradual implementation of a new electronic health record system in 2011 (14,15). Only patients aged 15 years or older were included due to the low prevalence of schizophrenia and bipolar disorder in younger individuals (see Supplementary Figure 1). Patients above 60 years of age were excluded because of the heterogeneous symptomatology observed in late-onset schizophrenia and bipolar disorder (16,17). Additionally, to avoid flagging non-informative cases, such as patients currently under assessment for one of the disorders, predictions were issued at the earliest three months after a patient’s first contact with the psychiatric services in the region. Contacts occurring after the diagnosis of schizophrenia or bipolar disorder were removed. If a patient moved out of the Central Denmark Region and later returned, no data was included from the interim period. No predictions were issued until three months after contact with the psychiatric services in the region had been re-established. The cohort definition and filtering are depicted in a flowchart found in Supplementary Figure 2.

### Outcome Definition

Diagnostic progression to schizophrenia was defined as the time of the first International Classification of Diseases, 10^th^ revision (ICD-10) diagnosis (codes in parentheses) of either schizophrenia (F20) or schizoaffective disorder (F25). Schizoaffective disorder was included as its ICD-10 definition is very close to that of schizophrenia. Diagnostic progression to bipolar disorder was defined as the time of the first ICD-10 diagnosis of either a manic episode (F30) or bipolar affective disorder (F31). Models were tested with three different outcomes: 1) diagnostic progression to schizophrenia or bipolar disorder (joint model), 2) diagnostic progression to schizophrenia, and 3) diagnostic progression to bipolar disorder.

For the joint outcome (1), the first occurring diagnosis was used. The joint outcome was motivated by the etiologic and phenomenological overlap between schizophrenia and bipolar disorder (18,19) and a desire to maximize the power of the analysis by including more patients with the outcome.

### Prediction Time Definition

To ensure that the developed models would have utility in clinical practice, we applied the “landmark model” framework for dynamic prediction (20,21). Landmark modelling involves selecting one or multiple time points of interest (the “landmark” or “prediction time”) – such as a certain type of clinical visit – from which to predict (future) outcomes. Data preceding the chosen timepoints is used to construct predictors, while predictions are made for pre-defined future periods, e.g., 6 months ahead. This approach offers several benefits for clinical prediction modelling, as it ensures that predictions are issued at relevant times and that the training and validation behaviour and performance mirrors the clinical setting (22).

We defined the prediction times as the day before a scheduled outpatient contact. Predictions issued a day before a contact allow practitioners to prepare possible interventions (e.g., a focus on symptoms compatible with progression to schizophrenia or bipolar disorder, or a Schedules for Clinical Assessment in Neuropsychiatry (SCAN) interview (23)). At each prediction time, separate models were trained to predict whether the three outcomes (1. schizophrenia or bipolar disorder, 2. schizophrenia, and 3. bipolar disorder) occurred within five years following the prediction time. Prediction times occurring after November 21, 2016 were removed as they did not have the required 5 years of follow-up (Figure 1C).

### Data Processing and Model Training

Figure 1 illustrates the model processing, training, and evaluation pipeline. Additional details are provided in the following sections.

### Predictor Construction

A full list of predictors is shown in Supplementary Table 1. Notably, only routine clinical data from the EHRs were considered for predictors. There was no data collection for the purpose of this study. The preprocessing of structured predictors from the EHR (e.g., diagnoses, medications, etc.) and text predictors (free-text clinical notes), respectively, are outlined below.

#### Structured predictors

Predictors from structured data were constructed by looking back a specified period of time (the lookbehind window) from each prediction time and extracting a single value for each predictor. When multiple values were present in the lookbehind window, we applied an appropriate aggregation function, such as the mean or count. If no values were present in the lookbehind window, a fallback value (e.g. 0 or NaN) was used. Predictors were created using lookbehind windows of 182 days, 365 days, and 730 days. Predictor construction was conducted using *timeseriesflattener* v2.2.0 (24). The structured predictors can be grouped into five categories: demographics, physical psychiatric hospital contacts, diagnoses, administered medications, and rating scales. Demographics included age and sex. Physical contacts included both inpatient and outpatient psychiatric hospital contacts, contacts with the somatic department, and admissions. Diagnoses included all psychiatric subchapters from the ICD-10 (F0-F9, see Figure 1D). Predictors derived from administered medication were based on ATC-codes at the group level, namely antidepressants, antipsychotics, first generation antipsychotics, second generation antipsychotics, benzodiazepines, lithium, clozapine, valproate, lamotrigine, pregabaline, selective serotonin reuptake inhibitors, serotonin-norepinephrine reuptake inhibitors, tricyclic antidepressants, and benzodiazepine related sleeping agents. ATC codes for the individual medications are specified in Supplementary Table 2. Rating scales included the Brøset Violence Checklist (25) and the 17-item Hamilton Depression Rating Scale (26).

#### Text predictors

Free-form clinical notes (the note types are specified in Supplementary Table 3) from the EHRs were embedded as numerical feature vectors using three different methods: 1) term-frequency of predefined words describing psychopathology, 2) term frequency-inverse document frequency (TF-IDF), and 3) sentence transformers (27), These three methods were chosen to cover a spectrum of approaches for analysing clinical notes, from highly specific psychopathological concepts to broader contextual information. Each method brings unique strengths: term-frequency of predefined words offers clinical relevance and interpretability, TF-IDF provides more coverage and allows for discovery of non-predefined important words, while sentence transformers capture semantic relationships and context, albeit with less interpretability.

The predictor set based on words describing psychopathology was constructed by counting the occurrence of a list of 365 words describing psychopathology derived by authors EP and AAD (both registrars in psychiatry), based on the Present State Examination (Danish Version). This simple approach is highly clinically relevant and easily interpretable. However, it is insensitive to the context and semantic relationships and has limited coverage.

In TF-IDF, each clinical note is represented as a vector, where dimensions correspond to unique words. The value in each dimension reflects the frequency of the term within the note balanced against its inverse frequency across all notes. This results in feature vectors for clinical notes that emphasize words that are distinctive of the particular note. While simple, TF-IDF is still widely used due to its interpretability and high performance both in terms of speed of computation and quality of results. Similarly to the approach using the term-frequency of words describing psychopathology, TF-IDF is insensitive to context and semantic relationships.

Sentence transformers use pre-trained deep neural networks to construct semantically meaningful text embeddings (27). Sentence transformers are trained using a triplet objective function or similar loss function which pulls semantically related sentences in the vector space closer, while pushing dissimilar ones apart. This ensures that clinical notes with comparable content (e.g., descriptions of hypomanic symptoms) have similar embeddings, despite variability in phrasing or style. Sentence transformers have achieved state-of-the-results on many text-based tasks (27–29), owing to their semantic understanding and tolerance for linguistic variations (e.g. spelling errors and different phrasings). However, sentence transformers are more computationally demanding than TF-IDF, and the embedding dimensions are not inherently interpretable.

TF-IDF models were trained on either all clinical notes or a single note type (“Subjective mental state”) in the training set, using *scikit-learn* v1.2.1 (30) with a dimensionality of 500 or 1000 features. Two different sentence transformer models were evaluated: *dfm-sentence-encoder-large* (31), and a version of *dfm-sentence-encoder-large* finetuned on the clinical notes in the training set. These models were chosen as they were the best-performing open-source encoder models for Danish on the ScandEval benchmark at the time of the analysis (32). Similar to the TF-IDF models, sentence transformer predictor sets were constructed based on either all clinical notes or “Subjective mental state” notes. The predictor set based on words describing psychopathology was constructed by counting the occurrence of a list of words describing psychopathology in all relevant notes.

The method used for aggregating structured predictors was adapted for embedded clinical notes. Each note embedding, comprising multiple dimensions (e.g. individual words for the predefined words and TF-IDF representations), was processed as follows: For each dimension, values within 730 days before the prediction time were averaged using the mean. This procedure was repeated for all dimensions, producing a series of time-averaged embeddings with the same dimensionality as before aggregation. Only a single lookbehind was used (730 days) to avoid very large feature sets. See Figure 1E-F for an illustration.

Models were trained using each text-based feature set to predict the outcomes in the training set. The text-based feature set achieving the highest 5-fold cross-validated area under the receiver operating characteristics curve (AUROC) was used for the analyses reported in the manuscript. For further details on text predictors and text models, see the Supplementary Methods.

### Model Training

Separate models were trained and optimized for each of the three outcomes separately (1. schizophrenia or bipolar disorder (joint model), 2. schizophrenia, and 3. bipolar disorder), each following the process outlined in the following sections.

#### Model types

Two common, state-of-the-art machine learning models, elastic net regularized logistic regression and Extreme Gradient Boosting (XGBoost) (33), were validated for prediction of diagnostic progression to schizophrenia and bipolar disorder from EHRs. Regularized logistic regression was chosen as it constitutes a strong baseline (34). XGBoost is a tree-based gradient-boosting algorithm that consistently achieves state-of-the-art results on numerous classification tasks (34,35). Neural network models were not included, as they tend to be outperformed by tree-based models on tabular classification tasks (35).

#### Hyperparameter tuning

Hyperparameter optimization was conducted for each model type to maximise the AUROC using the tree-structured parzen estimator algorithm in Optuna v3.4.0 (42) (Figure 1G).

Further details on the hyperparameters explored and their final values can be found in Supplementary Table 4. Hyperparameters were optimized by conducting 5-fold cross-validation on the training set.

#### Data augmentation

Data augmentation using synthetically generated data has been argued to improve performance on multiple classification tasks within healthcare (36,37). During training, experiments were therefore conducted to augment the training data with synthetic data generated using two methods, TabDDPM and SMOTE. TabDDPM (38) is the best-performing generative model for tabular data (39), while SMOTE (40) is a common method for generating synthetic samples of the minority class. Data augmentation with TabDDPM was conducted by first training TabDDPM on the training set and generating synthetic samples of the minority class (i.e. positive prediction times). Following hyperparameter tuning, models with the optimal hyperparameter configuration were trained with additional synthetic samples of the minority class corresponding to 1 to 10 times the number of real data points. Data augmentation with SMOTE was conducted within-fold, by training the model and adding 1 to 10 times the number of additional synthetic minority samples using imbalanced-learn v.0.12.2 (41). Models were not evaluated on synthetic data points during cross-validation, only on real data.

### Model Evaluation

The best-performing model following hyperparameter tuning was re-trained on the entire training set and applied to the test-set (Figure 1H). All evaluation metrics are based on the test-set unless otherwise stated. The AUROC was calculated for global performance.

Furthermore, we report sensitivity, specificity, positive predictive value (PPV), negative predictive value (NPV), and the median time from first positive prediction to the outcome at specific classification thresholds. These classification thresholds were based on a desired predicted positive rate, i.e. the proportion of highest-risk prediction times that are marked as positive. Predictor importance was estimated via information gain.

As a sensitivity analysis of the best-performing joint model, we tested how it performed in predicting schizophrenia or bipolar disorder separately.

### Robustness analyses

We performed stratified analyses of the stability of model predictions over time and demographics.

### Ethics

The use of EHRs from the Central Denmark Region for this study was approved by the Legal Office of the Central Denmark Region in accordance with the Danish Health Care act §46, Section 2. According to the Danish Committee Act, ethical review board approval is not required for studies based solely on data from EHRs (waiver for this project: 1-10-72-1-22). Data were processed and stored in accordance with the European Union General Data Protection Regulation and the project is registered on the internal list of research projects having the Central Denmark Region as data steward.

## Results

The cohort consisted of 25,805 unique patients with 403,424 outpatient visits eligible for prediction. Table 1 shows an overview of the number of patients and contacts in each split, along with demographic characteristics. The largest feature set contained 1,082 predictors, covering diagnoses, medications, admissions, and embeddings derived from clinical notes (Supplementary Table 1).

**Table 1:** Descriptive statistics for individual patients. (A) and outpatient contacts (B) that were eligible for prediction with a 5-year lookahead period. The train set includes the hospital units in Aarhus, Herning, Holstebro, Randers, Horsens, and Gødstrup, and the test set covers Silkeborg and Viborg.

**A. Patients**
|  | Overall | Train | Test |
| --- | --- | --- | --- |
| Number of patients | 24,449 | 20,224 | 4,225 |
| Female, n (%) | 13,843 (56.6) | 11,332 (56.0) | 2,511 (59.4) |
| Incident BP, n | 1,148 | 911 | 237 |
| Incident SCZ, n | 841 | 707 | 134 |
| Days from first contact to BP diagnosis, median [Q1,Q3] | 771.0<br>[355.8,1352.0] | 818.0<br>[385.0,1402.0] | 593.0<br>[250.0,1119.0] |
| Days from first contact to SCZ diagnosis, median [Q1,Q3] | 811.0<br>[387.0,1492.0] | 805.0<br>[386.0,1491.5] | 850.5<br>[420.2,1505.2] |
| Outpatient visits, median [Q1,Q3] | 9.0 [3.0,21.0] | 9.0 [3.0,21.0] | 10.0 [4.0,23.0] |
| Admissions, median [Q1,Q3] | 3.0 [1.0,6.0] | 3.0 [1.0,6.0] | 3.0 [1.0,5.0] |

**B. Outpatient visits**
|  |  | Overall | Train | Test |
| --- | --- | --- | --- | --- |
| Number of prediction times (outpatient visits) |  | 398,922 | 332,818 | 66,104 |
| Positive prediction times, n (%) |  | 19,505 (4.9) | 15,836 (4.8) | 3,669 (5.6) |
| Female, n (%) |  | 257,644 (64.6) | 212,579 (63.9) | 45,065 (68.2) |
| Age grouped, n (%) | 15-18 | 16,819 (4.2) | 15,270 (4.6) | 1,549 (2.3) |
|  | 19-20 | 22,558 (5.7) | 17,782 (5.3) | 4,776 (7.2) |
|  | 21-30 | 136,972 (34.3) | 112,024 (33.7) | 24,948 (37.7) |
|  | 31-40 | 100,437 (25.2) | 84,150 (25.3) | 16,287 (24.6) |
|  | 41-50 | 78,922 (19.8) | 67,336 (20.2) | 11,586 (17.5) |
|  | 51-60 | 43,214 (10.8) | 36,256 (10.9) | 6,958 (10.5) |
| Age, median [Q1,Q3] |  | 32.2<br>[24.2,42.5] | 32.4<br>[24.3,42.7] | 30.8<br>[23.4,41.7] |
| Incident BP within 5 years, n (%) |  | 11,624 (2.9) | 9,387 (2.8) | 2,237 (3.4) |

|  |  | **Overall** | **Train** | **Test** |
| --- | --- | --- | --- | --- |
| Incident SCZ within 5 years, n (%) |  | 8,319 (2.1) | 6,841 (2.1) | 1,478 (2.2) |
| N. admissions prior 2 years, median [Q1,Q3] |  | 0.0 [0.0,0.0] | 0.0 [0.0,0.0] | 0.0 [0.0,0.0] |
| F0 disorders prior 2 years, n (%) |  | 7,017 (1.8) | 5,693 (1.7) | 1,324 (2.0) |
| F1 disorders prior 2 years, n (%) |  | 28,536 (7.2) | 23,602 (7.1) | 4,934 (7.5) |
| F2 disorders prior 2 years, n (%) |  | 13,802 (3.5) | 12,099 (3.6) | 1,703 (2.6) |
| F3 disorders prior 2 years, n (%) |  | 139,808 (35.0) | 117,277 (35.2) | 22,531 (34.1) |
| F4 disorders prior 2 years, n (%) |  | 142,101 (35.6) | 122,426 (36.8) | 19,675 (29.8) |
| F5 disorders prior 2 years, n (%) |  | 23,294 (5.8) | 20,404 (6.1) | 2,890 (4.4) |
| F6 disorders prior 2 years, n (%) |  | 75,525 (18.9) | 64,022 (19.2) | 11,503 (17.4) |
| F7 disorders prior 2 years, n (%) |  | 5,976 (1.5) | 5,598 (1.7) | 378 (0.6) |
| F8 disorders prior 2 years, n (%) |  | 15,268 (3.8) | 13,373 (4.0) | 1,895 (2.9) |
| F9 disorders prior 2 years, n (%) |  | 67,306 (16.9) | 52,891 (15.9) | 14,415 (21.8) |

### Predictor selection

The text feature set that provided the best predictive performance on the training set was TF-IDF with 1000 features, trained on all note types (see Supplementary Table 5). Consequently, this feature set was used for all subsequent analyses.

The data augmentation method that provided the best predictive performance on the training set was TabDDPM with a 2x multiplier for the minority class (see Supplementary Table 6). That is, adding synthetic data equivalent to twice the number of positive outcomes (onset of schizophrenia or bipolar disorder within 5 years) yielded the greatest benefits. This configuration was used for all subsequent analyses. See the Supplementary Methods for further details.

### Model training

As shown in Figure 2A, the performance of the joint model, i.e. the model trained to predict the first occurring onset of either schizophrenia or bipolar disorder, approached the performance of the separate models when evaluated on each outcome separately on the training set. The performance of the joint model in the training phase is shown in Figure 2B. XGBoost was superior to logistic regression in all cases (see Supplementary Table 7). Figure 2B shows that the feature set including structured data, text, and synthetic data performed slightly better than the other feature sets on the training set. At a threshold of the 4% highest risk predictions marked as positive, the median lead time for the model to flag patients who will develop schizophrenia or bipolar disorder was 0.7 years.

**Figure 2:**
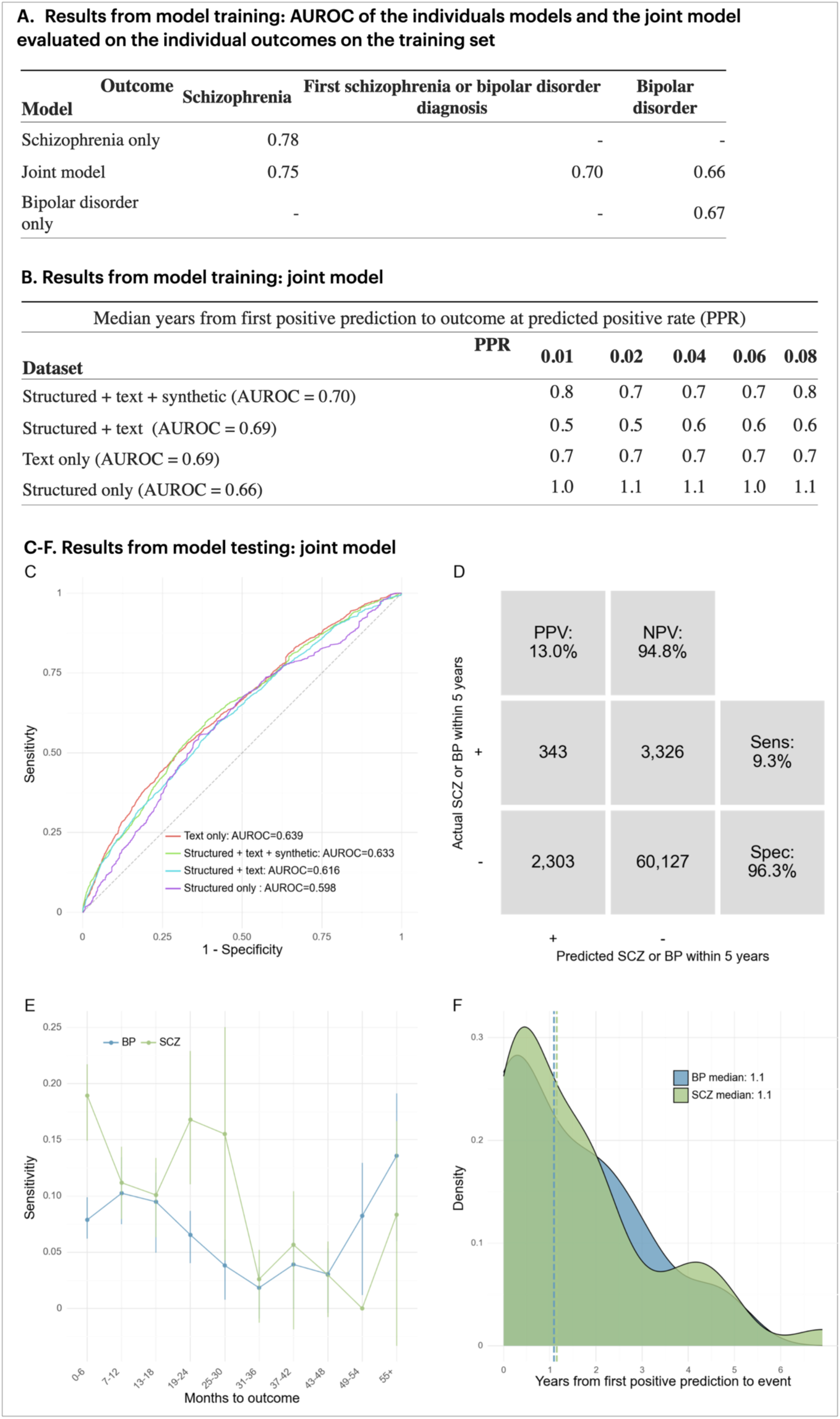
A) Out-of-fold performance (AUROC) of models trained on individual outcomes and the joint model (*structured + text + synthetic* feature set), on the outcomes for each model. The diagonals show the performance of the outcome the respective model was trained on**. B)** AUROC and median years to first positive prediction for the best model for each dataset on out-of-fold predictions on the training set. The best model was XGBoost in all cases. **C)** Receiver operating characteristics (ROC) curve of the best-performing models for each feature set on the test set. The model with the highest AUROC on the test (*text only)* was used in panels C-E with a classification threshold corresponding to 4% positives. **D)** Confusion matrix. PPV: Positive predictive value. NPV: Negative predictive value. **E)** Sensitivity by months from prediction time to event, stratified by outcome (BP=bipolar disorder, SCZ=schizophrenia). **F)** Time (years) from the first positive prediction to the patient receiving a diagnosis of bipolar disorder or schizophrenia. The dotted lines indicate the median time for each group.

### Joint model (predicting schizophrenia or bipolar disorder) testing

When applied to the test set (Figure 2C-F), the XGBoost joint model using only text features performed best, achieving an AUROC of 0.63. Figure 2D shows the confusion matrix using this model and a threshold based on a 4% predicted positive rate. The PPV was 13.0%, indicating that for every 7.7 positive predictions, one prediction time was followed by a diagnosis of schizophrenia or bipolar disorder within 5 years. The sensitivity at the level of prediction times was 9.3%, and 13.5% of all patients who received a diagnosis of schizophrenia or bipolar disorder were predicted positive at least once (Table 2A). The median time from the first positive prediction to the outcome was 1.1 years (see Figure 2F). As shown in Figure 2E, for the joint model, sensitivity was generally higher for predicting schizophrenia compared to bipolar disorder.

**Table 2A:** Performance by predicted positive rate for the best-performing model on the test set (XGBoost using only TFIDF-1000 text features).

| Predicted positive rate | True prevalence | PPV | NPV | Sens | Spec | FPR | FNR | Acc | TP | TN | FP | FN | F1 | MCC | % of all patients with BP or SCZ captured | Median years from first positive to SCZ diagnosis | Median years from first positive to BP diagnosis | Median years from first positive to first SCZ or BP diagnosis |
| --- | --- | --- | --- | --- | --- | --- | --- | --- | --- | --- | --- | --- | --- | --- | --- | --- | --- | --- |
| 8.0% | 5.6% | 13.2% | 95.1% | 19.1% | 92.6% | 7.4% | 80.9% | 88.6% | 699 | 57,841 | 4,589 | 2,970 | 15.6% | 9.9% | 22.8% | 1.4 | 0.8 | 0.9 |
| 6.0% | 5.6% | 13.9% | 95.0% | 15.0% | 94.5% | 5.5% | 85.0% | 90.1% | 552 | 59,016 | 3,414 | 3,117 | 14.5% | 9.2% | 19.9% | 1.3 | 0.8 | 0.9 |
| 4.0% | 5.6% | 13.0% | 94.8% | 9.3% | 96.3% | 3.7% | 90.7% | 91.5% | 343 | 60,127 | 2,303 | 3,326 | 10.9% | 6.6% | 13.5% | 1.1 | 1.1 | 0.9 |
| 2.0% | 5.6% | 14.2% | 94.6% | 5.1% | 98.2% | 1.8% | 94.9% | 93.0% | 188 | 61,296 | 1,134 | 3,481 | 7.5% | 5.4% | 7.8% | 0.6 | 0.2 | 0.5 |
| 1.0% | 5.6% | 20.4% | 94.6% | 3.7% | 99.2% | 0.8% | 96.3% | 93.9% | 135 | 61,902 | 528 | 3,534 | 6.2% | 6.5% | 4.3% | 0.4 | 0.5 | 0.3 |

Abbreviations: % of all patients with BP or SCZ captured, percentage of all patients who received a diagnosis of either bipolar disorder or schizophrenia, who had at least one positive prediction; F1, the harmonic mean of the PPV and sensitivity; FN, false negatives. Numbers are prediction times (outpatient contacts); FNR, false negative rate; FP, false positives. Numbers are prediction times; FPR, false positive rate; MCC, Matthew's correlation coefficient; Median years from first positive to SCZ/BP diagnosis, for all patients with at least one true positive, the number of years from their first positive prediction to having developed schizophrenia/bipolar disorder; NPV, negative predictive value; PPV, positive predictive value; Predicted positive rate, the proportion of contacts predicted positive by the model. Since the model outputs a predicted probability, this is a threshold set during evaluation; TN, true negatives. Numbers are prediction times; TP, true positives. Numbers are prediction times; True prevalence, the proportion of contacts that qualified for the schizophrenia or bipolar disorder outcome within the lookahead window.

A list of the 10 most important features for the joint model according to information gain is shown in Table 2B. Notably, text embeddings of words, including “discharge”, “voices”, and “admission” were found to be highly influential for the model.

**Table 2B:**
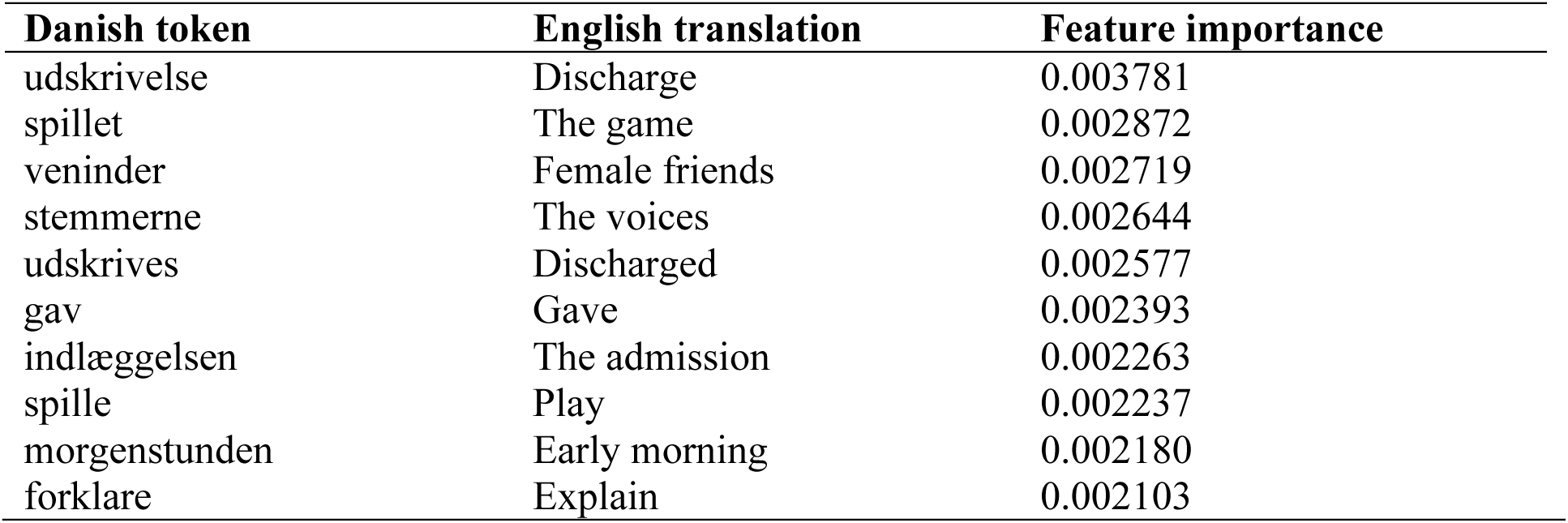
Top 10 most important features by information gain in the best-performing (text-only) joint model. All features are TF-IDF with a 2-year lookbehind and mean aggregation function.

| Danish token | English translation | Feature importance |
| --- | --- | --- |
| udskrivelse | Discharge | 0.003781 |
| spillet | The game | 0.002872 |
| veninder | Female friends | 0.002719 |
| stemmerne | The voices | 0.002644 |
| udskrives | Discharged | 0.002577 |
| gav | Gave | 0.002393 |
| indlæggelsen | The admission | 0.002263 |
| spille | Play | 0.002237 |
| morgenstunden | Early morning | 0.002180 |
| forklare | Explain | 0.002103 |

### Models trained to predict either schizophrenia or bipolar disorder

The results for the models trained to predict either schizophrenia or bipolar disorder separately are shown in Supplementary Figures 3-4 and Supplementary Tables 8-9. The models predicting schizophrenia obtained the best performance, with an AUROC on the test set of 0.80 for the best model. At a 4% predicted positive rate, sensitivity was 19.4% and the PPV was 10.8% on the test set. Bipolar disorder proved more difficult, with the best model achieving an AUROC of 0.62 on the test set. At a 4% predicted positive rate, sensitivity was 9.9% and the PPV was 8.4% on the test set. As shown in Supplementary Table 10, the best models on both the training and set for the separate outcomes used the text-only feature set or the feature set including synthetic data.

The joint model achieved an AUROC of 0.74 for the schizophrenia-only outcome and 0.57 for the bipolar disorder-only outcome on the test set (Supplementary Table 11).

### Robustness analyses

Figure 3 shows that the performance of the joint model is stable across sex and age. The model performs slightly better on relatively young and old patients. Performance is quite stable across levels of time from first visit, with some instability at the extremes, likely partially owing to lack of data. No noticeable trends are observed in the performance across calendar time. Supplementary Figure 5 shows the schizophrenia model to be highly robust across stratifications, with slightly better performance for older patients. As shown in Supplementary Figure 6, the bipolar model is less robust, particularly across calendar time, with a noticeable dip in performance around Q3 2015.

**Figure 3:**
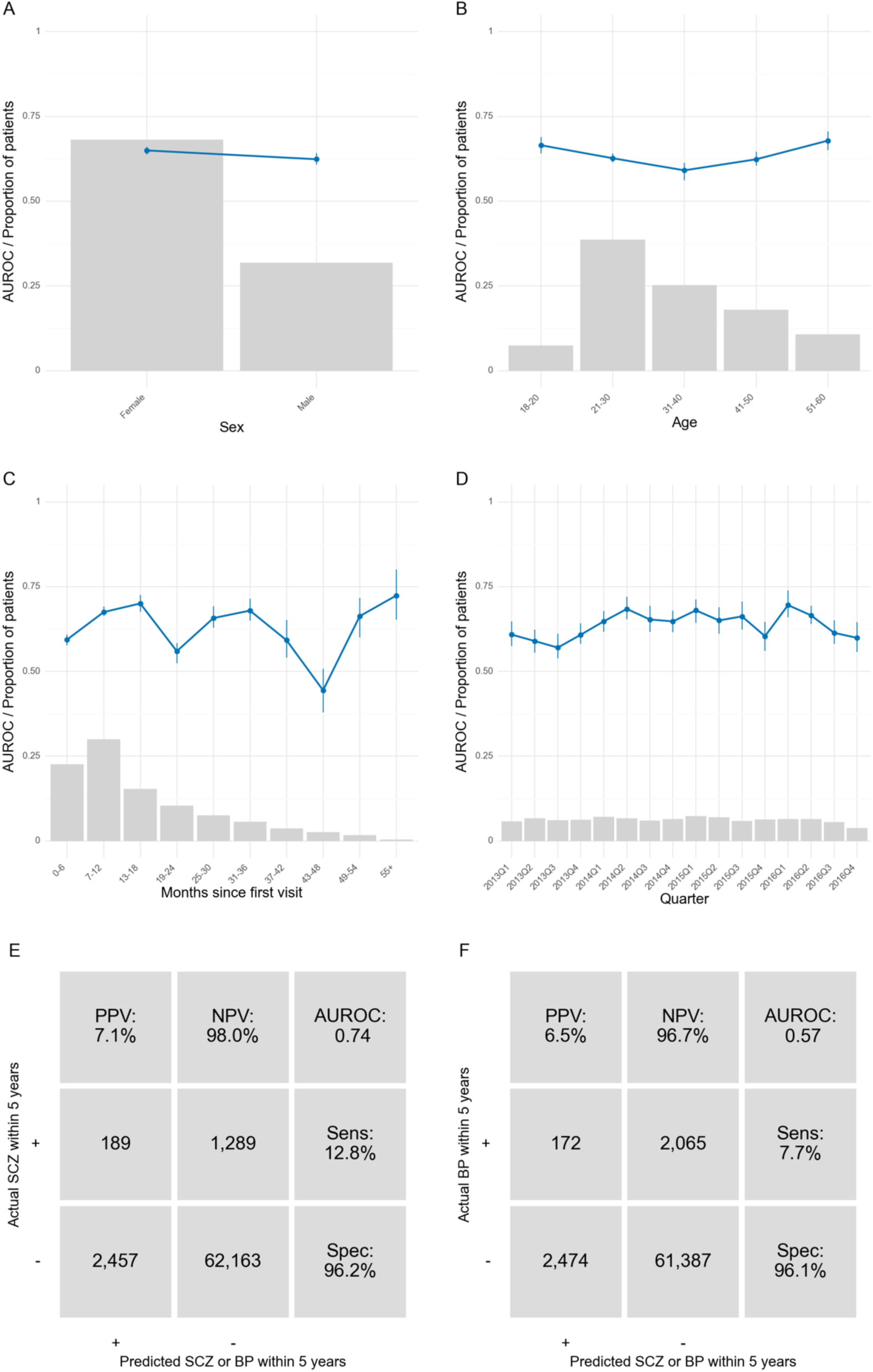
Robustness of the joint model across stratifications on the test set. Blue line is the area under the receiver operating characteristics curve. Grey bars represent the proportion of prediction times in each bin. Error bars are 95%-confidence intervals from 100-fold bootstrap. Due to the low n in some of the bins, some bootstrap folds contained only one class. This resulted in missing error bars for those bins. Panels E and F show the performance when evaluating the joint model on the schizophrenia and bipolar disorder outcome, respectively, using a 4% PPR.

## Discussion

This study investigated the feasibility of predicting diagnostic progression to schizophrenia or bipolar disorder within 5 years, among patients with pre-existing mental illness. A model predicting the progression to either of the two disorders achieved an AUROC of 0.64 on the test set, with a notable disparity in predictive performance between the two disorders. While the best model for predicting progression to schizophrenia achieved an AUROC of 0.80 on the test set, predicting bipolar disorder proved more challenging with the best model achieving an AUROC of 0.62. This discrepancy may be attributed to the relatively larger heterogeneity within bipolar disorder compared to schizophrenia, and the distinctiveness of the psychotic symptoms of schizophrenia. Bipolar disorder covers a wide spectrum of symptoms, with some individuals presenting with mania (Bipolar I), and some with no manic episodes but rather frequent depressive episodes (Bipolar II). Additionally, Bipolar II is very similar to major depressive disorder, often making it challenging to distinguish between the two conditions (43). The distinction between bipolar I and II is not made in ICD-10 but both types are present in the patient population analysed in this study. In contrast, most individuals with schizophrenia have the paranoid schizophrenia subtype (ICD-10: F20), accounting for approximately 72% in the Central Denmark Region (44), which is relatively more homogenous in its presentation.

A substantial drop in performance was observed when moving from the training set to the test set, particularly for the joint model and the bipolar disorder model. The training and test sets contained data from different hospitals in the Central Denmark region. This indicates that significant distribution shifts can occur even in a relatively homogenous population in close geographical proximity using the same healthcare system and clinical guidelines. These shifts might be caused by slightly different patient populations and/or variations in diagnostic practices. Indeed, a recent study indicated that diagnosing of schizotypal disorder (F21) varies markedly across regions in Denmark with the relative incidence being much higher in the Capital region compared to the rest of Denmark (44). These findings provide indications that mental disorders, which typically have large within-disorder variability in their expression, might pose difficult targets for predictions, particularly across sites. Additionally, the change in performance across sites supports the argument that external validation should not be a strict requirement for scientific publication or model evaluation (45). Rather, models should be tested in the specific context where they will be applied.

Model performance was mainly driven by the inclusion of text-based predictors extracted from clinical notes. Indeed, models trained with both structured and text-based predictors performed practically equivalently to models trained with only text-based features. This underscores the importance of text in clinical prediction modelling within psychiatry (46,47). A representation of text based on TF-IDF was found to slightly outperform a general-purpose Transformer encoder model as well as a Transformer encoder model continuously pre-trained on the training data. This suggests that the main predictive signal of the clinical notes likely comes from specific words or short phrases. The transparency and high efficiency in computational terms make TF-IDF models valuable in a clinical context, as they are fast to use and inherently interpretable. Inspection of the most important words by feature importance and the context they appeared in within the EHRs, revealed that many were related to hospital admission or psychiatric symptoms. Specifically, “Admission” and “Discharge” directly pertained to hospitalization. “Play” and “The game” often described patients’ interactions with staff or other patients (playing board games etc.) during their stay. “Early morning” frequently appeared at the beginning of notes detailing a day during an admission. Symptom-related terms included “The voices,” typically referring to auditory hallucinations, while “female friends” was often used to describe social interactions or lack thereof (i.e., social withdrawal). “Explain” commonly appeared when patients struggled to articulate the reasons for their actions or experiences, potentially indicating delusions or derealization.

Augmenting the data with synthetic samples of the minority class yielded little to no performance gain on the test set. This may be caused by the sample being too small or too difficult to learn generalizable patterns from. For instance, the data contained a considerable proportion of missing values which had to be imputed before training the generative model, as TabDDPM does not support it. This might impair learning, as many values would be repeated due to imputation.

Performance from our models predicting schizophrenia is in line with the literature, with e.g. Irving et al. (46) achieving a Harrell’s C of 0.79-0.86 for ten-year survival prediction of onset of psychosis from an index date. Harrell’s C and AUROC are equivalent in binary outcomes, but direct comparisons cannot be made with censored data as those used by Irving and colleagues (46). Irving et al. made a single prediction at an index date and only provide aggregate performance metrics such as Harrell’s C, Brier score, and the calibration slope. Wang et al. (51) achieved an AUROC of 0.80 in predicting early-onset bipolar disorder 3 years into the future from a single randomly sampled time between age 10-25 per patient. The performance discrepancy in bipolar disorder prediction likely stems from the more age-restricted cohort in Wang et al., and major differences in the definition of the outcome with Wang et al. requiring 1) at least two ICD codes for bipolar disorder and 2) predominance of bipolar disorder diagnoses, and 3) treatment with at least two medications commonly used for bipolar disorder. In summary, direct comparison is difficult, as most studies only make a single prediction at an index date, or only report aggregated measures such as AUROC or the C-index. In contrast, we issue predictions dynamically at clinically relevant times (before an outpatient contact) and report performance at multiple decision thresholds to facilitate maximal clinical utility and critical scrutiny.

If applied within the Psychiatric Services of the Central Denmark Region, the model’s positive predictions should be automatically presented to the staff through the EHR system, enabling intervention at the level of the individual patient. Specifically, increased focus on symptoms compatible with schizophrenia or bipolar disorder, e.g., via a focused diagnostic interview at the next outpatient consultation would seem reasonable. If no symptoms of bipolar disorder or schizophrenia are present, the model should be disabled for the specific patient for a substantial period to reduce alarm-fatigue among the staff. Models predicting schizophrenia might be more suitable for implementation than those predicting bipolar disorder due to substantially better predictive performance. Joint models, predicting either of the two disorders, show potential, but separate models perform better.

### Limitations

The study should be interpreted considering the following limitations. First, the data is restricted to patients under psychiatric treatment and does not contain information from primary care. Consequently, the prediction models are primarily useful for patients who are progressing from another mental disorder to schizophrenia or bipolar disorder. Patients whose initial contact to the psychiatric services is due to clinical suspicion of schizophrenia or bipolar disorder will not see additional benefits from the model. Second, text models might be at high risk of fitting to already present clinical suspicion and thereby provide less value. As shown in Figure 2B, the median time from the first positive prediction to the outcome is highest for the model only using structured predictors, despite having the lowest overall performance in terms of AUROC. This indicates that while text models might lead to more cases being identified correctly, this might be at the cost of less lead time.

## Conclusion

The present study developed and validated models for predicting progression to schizophrenia or bipolar disorder using electronic health record data. The model predicting schizophrenia performed substantially better than the model predicting bipolar disorder, likely due to heterogenic clinical manifestations of the latter. Lastly, text-based features from clinical notes show great promise for improving the prediction of psychiatric outcomes and should be explored further.

## Data Availability

Due to the sensitive nature of the data it can not be made available. All code is freely available online.

https://github.com/Aarhus-Psychiatry-Research/psycop-common

## Acknowledgement Section

Author contributions

The study was conceptualized and designed by all authors. The coding and statistical analyses were carried out by LH relying on a codebase jointly developed between LH, MB, KE, SK, and JD. All authors contributed to the interpretation of the results. LH wrote the first draft of the manuscript, which was subsequently revised for important intellectual content by the remaining authors. All authors approved the final version of the manuscript prior to submission. LH had full access to all the data in the study and takes responsibility for the integrity of the data and the accuracy of the data analysis.

The authors thank Bettina Nørremark from Aarhus University Hospital – Psychiatry for assistance with extraction of data.

