## Supplementary Materials for "Predicting diagnostic progression to schizophrenia or bipolar disorder via machine learning applied to electronic health record data"

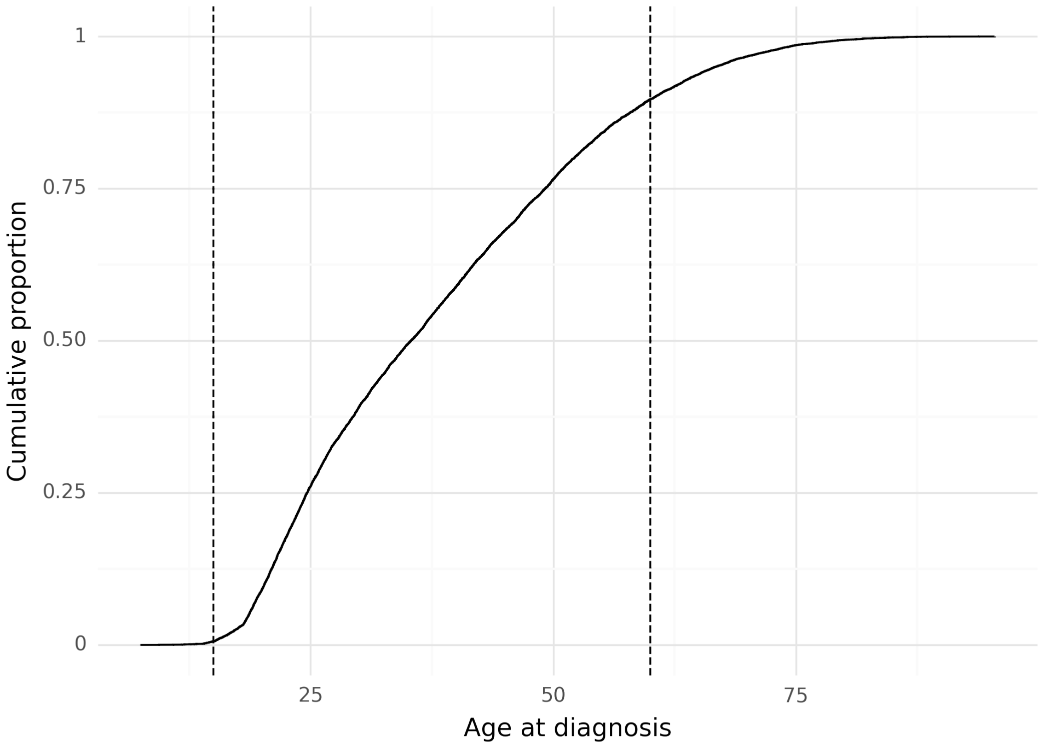


***Supplementary Figure 1****: Cumulative proportion of age at diagnosis of schizophrenia or bipolar disorder. Dashed lines indicate the minimum and maximum ages (15 and 60).*

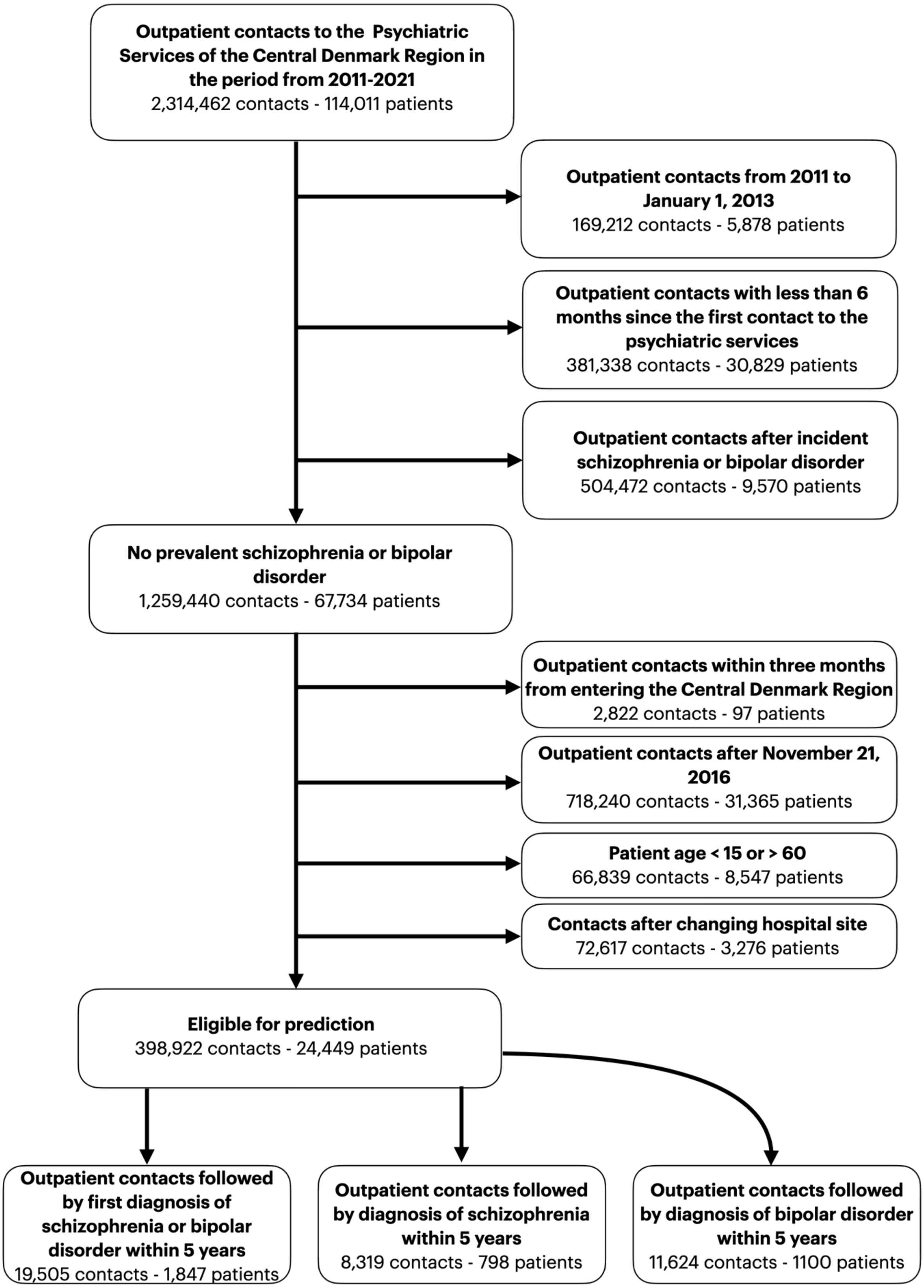


*Supplementary Figure 2: Filtering of eligible prediction times before model training.*

***Supplementary Table 1:*** *Predictor description. Moved to the end of the Supplementary Materials (after References) for readability purposes.*

***Supplementary Table 2****: ATC codes for medication grouping*

| **Medication group** | **ATC codes** |
|---|---|
| Antipsychotics | N05A* (exclude: N05AN01 (lithium)) |
| Antidepressants | N06A* |
| First generation antipsychotics | N05AF01, N05AG02, N05AD01, N05AF05, N05AD03, N05AD05, N05AF03 |
| Second generation antipsychotics | N05AL05, N05AX12, N05AH05, N05AX16, N05AX15, N05AE02, N05AE05, N05AH03, N05AX13, N05AH04, N05AX08, N05AE04, N05AE03 |
| Clozapine | N05AH02 |
| Lithium | N05AN01 |
| Valproate | N03AG01 |
| Lamotrigine | N03AX09 |
| Benzodiazepines | N05BA |
| Pregabaline | N03AX16 |
| SSRIs | N06AB* |
| SNRIs | N06AX21, N06AX16 |
| TCAs | N06AA* |
| Selected NASSA | N06AX11, N06AX03 |
| Benzodiazepine related sleeping agents | N05CF01, N05CF02 |

***Supplementary Table 3:*** *Description of the content of each of the note types.*

| Danish name | English name | Description |
|---|---|---|
| Aftaler, Psykiatri | Appointments, Psychiatry | Description of concrete care- and treatment-related appointments and agreements with and about the patient. |
| Aktuelt socialt, Psykiatri | Current social functioning | Description of the patient's current social situation: relationship to the family, civil status, residential-, occupational-, and economic conditions, and contact with the social services. Documentation of ongoing treatment plans in relation to the patient's social relationships. |
| Aktuelt psykisk | Subjective mental state | Description of the development, progress, and current status of the patient's mental illness. |
| Aktuelt somatisk, Psykiatri | Subjective physical state | Description of the patient's current and chronic somatic illnesses and symptoms. Information on current treatment in relation to the patient's physical condition. |
| Konklusion/vurdering | Conclusion/evaluation | Aggregation and interpretation of all findings and decisions based on an interview (e.g in relation to an outpatient visit or during an inpatient stay). |
| Kontaktårsag | Reason of contact | The reason for the patient's in- or outpatient treatment course. |
| Objektivt psykisk | Current objective mental state | Objective assessment of the patient's mental state, including state of consciousness, orientation, intelligence, psychomotor function, mood, delusions, psychotic symptoms, etc. |
| Observation af patient, Psykiati | Observation of patient, Psychiatry | Description of an inpatient's mental symptoms, behaviour, reactions towards relatives, other patients, staff, etc. |

| | | |
|---|---|---|
| Samtale med behandlingssigte | Conversation with treatment aim | Documentation of the conversation’s purpose and attendees. |
| Semistruktureret diagnostisk interview | Semi-structured diagnostic interview | Registration that a semi-structured diagnostic interview has been conducted. Description of the method and documentation of result and conclusion. |
| Telefonnotat | Telephone note | Documentation of telephone conversations with a patient or the patient’s guardian of non-clinical character. |

***Supplementary Table 4****: The search spaces and optimal values for each model type on the feature set including both structured variables and text on the training set (joint model). The imputation method in bold was the best performing.*

| | |
|---|---|
| **XGBoost** (structured + text) | |
| Imputation method | Mean, median, **most frequent**, no imputation |
| N. estimators | [100; 1200], 315 |
| Alpha | [$1^{-8}$; 0.1], 0.0039071871 |
| Lambda | [$1^{-8}$; 1], 0.0006726613 |
| Learning rate | [$1^{-8}$; 0.1], 0.0357272581 |
| Gamma | [$1^{-8}$; 0.001], 0.0000050752 |
| Max depth | [3; 8], 6 |
| **Logistic regression** (structured + text) | |
| Imputation method | **Mean**, median, most frequent |
| Penalty | Elastic net |
| Normalization | Z-score |
| C | [$1^{-8}$; 10], 0.0021408992 |
| L1 ratio | [0; 1], 0.8843476177 |
| Max iterations | [500;1000], 884 |

### Supplementary Methods

#### Text models

**Words describing psychopathology**

To assess the performance of a clinically informed embedding, 365 terms deemed important for describing psychopathology were chosen by authors EP and AAD (both registrars in psychiatry) and used in Hansen et al. [1]. The terms were based on the Present State Examination (Danish version) [2] and the International Statistical Classification of Diseases and Related Health Problems 10th Revision (ICD-10) [3]. The embedding was constructed by counting the term frequency of each term in each note.

**Term frequency-inverse document frequency embeddings**

Term frequency-inverse document frequency (TF-IDF) is a widely used measure of the importance of a term in a specific document (a clinical note) within a corpus (all clinical notes. TF-IDF is calculated by counting the frequency of all terms within all documents and scaling by how many documents the term appears in. As a result, the higher the TF-IDF score, the more important and distinct the term is for the particular document. This has the added benefit of down-weighting common terms, as they will appear in most documents.

We trained TF-IDF models using the *scikit-learn* v1.2.1 package [4], with *min_df* set to 2 and *max_df* set to 0.9, and the remaining parameters to their default values. *Min_df* refers to the minimum number of documents a term must occur in to be included in the analysis, and *max_df* refers to the maximum proportion of documents a term is allowed to occur in to be included in the analysis. A grid of 2x2 TF-IDF models were trained by varying the maximum number of features (500 or 1.000) and the type of clinical notes to consider (only "Subjective mental state" or all available clinical notes). The models were trained on the training set and used to embed the notes from all splits.

**Embeddings from large language models**

Using embeddings of text from large language models (LLMs) has gained increased traction for a wide variety of use cases from classification tasks to determining semantic similarity of documents [5]. In brief, LLMs are neural networks trained on vast text corpora on a self-supervised task such as masked language modelling or causal language modelling. Through this pretraining, the models learn to encode the semantics of texts in a vector representation. To use this for downstream tasks, such as classification, the models are then either fine-tuned for the particular task, or the embeddings are extracted and passed to a different model. As we want to combine text with other structured predictors (such as diagnoses), we use the latter option. To extract the embeddings for each clinical note, we summarize the embeddings from the last hidden layer of the models for each token in the note by mean pooling.

*Dfm-encoder-large*

To embed the clinical notes, we employed the *danish-foundation-models/encoder-large-v1 (*dfm-encoder*)* model [6], which is a continued pretraining of *NbAiLab/nb-bert-large* [7] on several large-scale Danish corpora [8]. The dfm-encoder was chosen as it, at the time of writing, is the best-performing encoder model for Danish tasks on the ScandEval benchmark [9]. All notes were embedded using the frozen model.

*Finetuned dfm-encoder*

Medical language is highly specialized and idiosyncratic. As a consequence, language models, which are often trained primarily on web data, might find medical language and clinical notes to be out of domain, and therefore see reduced performance. To alleviate this, we finetuned the dfm-encoder on the clinical notes in the training set using unsupervised SimCSE [10]. SimCSE uses a simple contrastive learning objective for training language models. Contrastive learning involves training a model to distinguish between positive pairs, i.e., similar text instances, and negative pairs, i.e., dissimilar text instances, by maximising the similarity of positive pairs and minimizing the similarity of negative pairs. In unsupervised SimCSE, the positive pairs are created by feeding the same sentence through the model encoder twice, relying on the noise from dropout [11] to produce slightly different embeddings. Negative pairs are randomly sampled from the minibatch, which represents a subset of the training data utilized for updating the parameters of the model. During each training iteration, an anchor text instance is chosen and the model is tasked to predict the positive example among multiple negatives.

Similarly to the procedure for the TF-IDF models, we finetuned the dfm-encoder separately on “Subjective mental state” only or on all available clinical notes and used the respective model to embed the notes.

### Text embedding experiment

Training large language models and using them to embed clinical notes is extremely computationally demanding. Therefore, it was deemed infeasible to test all combinations of text predictors alongside the structured predictors. Instead, an experiment was conducted to find which text embedding model to use in the main analysis described in the methods section of the manuscript. Additionally, we investigated whether embedding only a single note type ("Current mental state") would be competitive with embedding all available note types (see Supplementary Table 3 for an overview), as this note type was found to be the most predictive in a previous study on the same cohort [12].

The experiment was constructed to mirror the conditions of the main analysis. That is, predictor sets were constructed using the same procedure as explained in the methods section of the manuscript regarding the definition of prediction times, inclusion criteria, wash-in periods, etc. A predictor set was constructed for each set of embedded text using a 730-day lookbehind window with the mean as aggregation function, except for the pre-defined clinical words which used count as aggregation function. An XGBoost model with default hyperparameters was trained on each of the predictor sets to predict whether the patient will be diagnosed with schizophrenia or bipolar disorder within the next five years from each prediction time using 5-fold cross-validation on the training set. Supplementary Table 5 shows the mean out-of-fold AUROC for each model.

***Supplementary Table 5:*** *Mean (standard deviation) out-of-fold AUROC of models using only the embedded text from the specified note type and model on predicting 5-year transition to either schizophrenia or bipolar disorder on the training set. The best model is highlighted in bold, and the second best is in italics.*

| Notes/model | TF-IDF-500 | TF-IDF-1000 | Dfm-encoder | Finetuned dfm-encoder | PSE keywords |
|---|---|---|---|---|---|
| Current mental state | 0.614 ±0.018 | 0.644 ±0.029 | *0.654 ±0.032* | 0.645 ±0.02 | |
| All notes | 0.649 ±0.031 | **0.671 ±0.026** | 0.648 ±0.027 | 0.642 ±0.029 | 0.644 ±0.026 |

### Model training and hyperparameter tuning

After identifying which text embedding to use for the main experiment (TF-IDF 1000 on all notes), three predictor sets were constructed. One using only structured features, one using only text features (TF-IDF-1000), and one using both structured and text features. On each feature set, for each model, hyperparameter tuning was conducted for 150 trials by optimizing AUROC. Supplementary Table 4 shows the search spaces for the two models, and Supplementary Table 7 shows the performance of the best model for each feature set.

### Augmentation with synthetic data

Synthetic data generation (SDG) is the process of generating artificial data with similar characteristics as the original data. SDG is useful for a multitude of purposes such as improving the fairness of models [13] and improving the performance and generalisability of downstream models [14]. When used to add additional artificial samples to a dataset, SDG can be viewed as a type of data augmentation, which is particularly useful in data scarce situations [15] or in cases with pronounced class imbalance [16]. Although the majority of work has been in the field of computer vision or natural language processing, synthetic tabular data generation has been shown to improve performance within the tabular domain as well [17].

Since the present study deals with a case of severe class imbalance, we sought to investigate whether augmentation with synthetic data could improve the performance of our classifiers. We tested two models for this purpose: SMOTE [18], a widely used method for generating synthetic data in the minority class by interpolating between existing samples, and TabDDPM [19], a recently introduced diffusion model for generating synthetic tabular data.

### Data augmentation experiment

Training and generating data with both TabDDPM and SMOTE is computationally expensive. In particular, TabDDPM requires fitting a neural network whereas the K-nearest neighbour step of the SMOTE algorithm is slow with large sample sizes. To keep the computational demands feasible, we only applied the two methods to the best-performing model and predictor set configuration, i.e., after hyperparameter optimization. As TabDDPM performs best with normalized values, we additionally z-score normalized all predictors before feeding to TabDDPM.

Three TabDDPM model were fit to only the positive prediction times in the training set using the largest predictor set, and trained for 4.000 iterations with a learning rate of 0.0001, 0.0005, and 0.001, respectively. The model with a learning rate of 0.0005 achieved the lowest loss, which was used for the following TabDDPM models. Afterwards, separate TabDDPM models were trained for each outcome (joint, schizophrenia only, bipolar disorder) and were used to generate synthetic samples equal to 10x the number of positive samples in the training data for the minority class with 500 diffusion timesteps.

Classification models with the same hyperparameters as the best-performing model on the training set were trained with varying ratios of added synthetic samples of the minority class from TabDPPM and SMOTE. The models were evaluated only on real data during cross-validation to not artificially inflate performance estimates if synthetic data was easier to predict than real data. Supplementary Table 6 shows the results. SMOTE was applied at each fold during cross-validation, and synthetic data from TabDDPM was loaded before cross-validation to reduce computational demands. The best-performing model (TabDDPM with a sample multiplier of 2) was used for the main analysis in Figure 2.

***Supplementary Table 6:*** *Mean (standard deviation) out-of-fold AUROC of the best performing model (XGBoost) with additional synthetic samples added for the minority class. The top row shows the ratio of real data to synthetic data, i.e., 1 indicates equal proportions, 10 indicates 10 times as many synthetic samples as real samples. The best performing model is in bold, the second best in italics.*

| **Method** | **1** | **2** | **3** | **4** | **5** | **6** | **7** | **8** | **9** | **10** |
|---|---|---|---|---|---|---|---|---|---|---|
| TabDDPM | 0.692 ±0.018 | **0.7 ±0.017** | *0.698 ±0.041* | 0.696 ±0.02 | 0.689 ±0.021 | 0.692 ±0.04 | 0.691 ±0.016 | 0.685 ±0.026 | 0.69 ±0.026 | 0.684 ±0.023 |
| SMOTE | 0.691 ±0.031 | 0.686 ±0.035 | 0.689 ±0.034 | 0.688 ±0.03 | 0.686 ±0.029 | 0.684 ±0.029 | 0.684 ±0.032 | 0.684 ±0.03 | 0.681 ±0.03 | 0.682 ±0.026 |

***Supplementary Table 7:*** *Mean (standard deviation) out-of-fold AUROC of the best performing model for each model type on each data set.*

| **Model** | **Structured only** | **Text only** | **Structured + text** |
|---|---|---|---|
| XGBoost | 0.662 ±0.018 | 0.690 ±0.02 | 0.688 ±0.034 |
| Logistic Regression | 0.659 ±0.019 | 0.679 ±0.028 | 0.681 ±0.022 |

### Results for models stratified by outcome

**A. Results from model training: schizophrenia model.**

| Median years from first positive prediction to outcome at predicted positive rate (PPR) | | | | | |
|---|---|---|---|---|---|
| **Dataset** / **PPR** | **0.01** | **0.02** | **0.04** | **0.06** | **0.08** |
| Structured + text + synthetic (AUROC = 0.77) | 0.7 | 0.5 | 0.7 | 0.8 | 0.9 |
| Structured + text (AUROC = 0.78) | 0.5 | 0.5 | 0.6 | 0.7 | 0.9 |
| Text only (AUROC = 0.77) | 0.6 | 0.5 | 0.7 | 0.7 | 0.7 |
| Structured only (AUROC = 0.72) | 0.9 | 0.9 | 0.8 | 0.9 | 1.0 |

**B-E. Results from model testing: schizophrenia model**

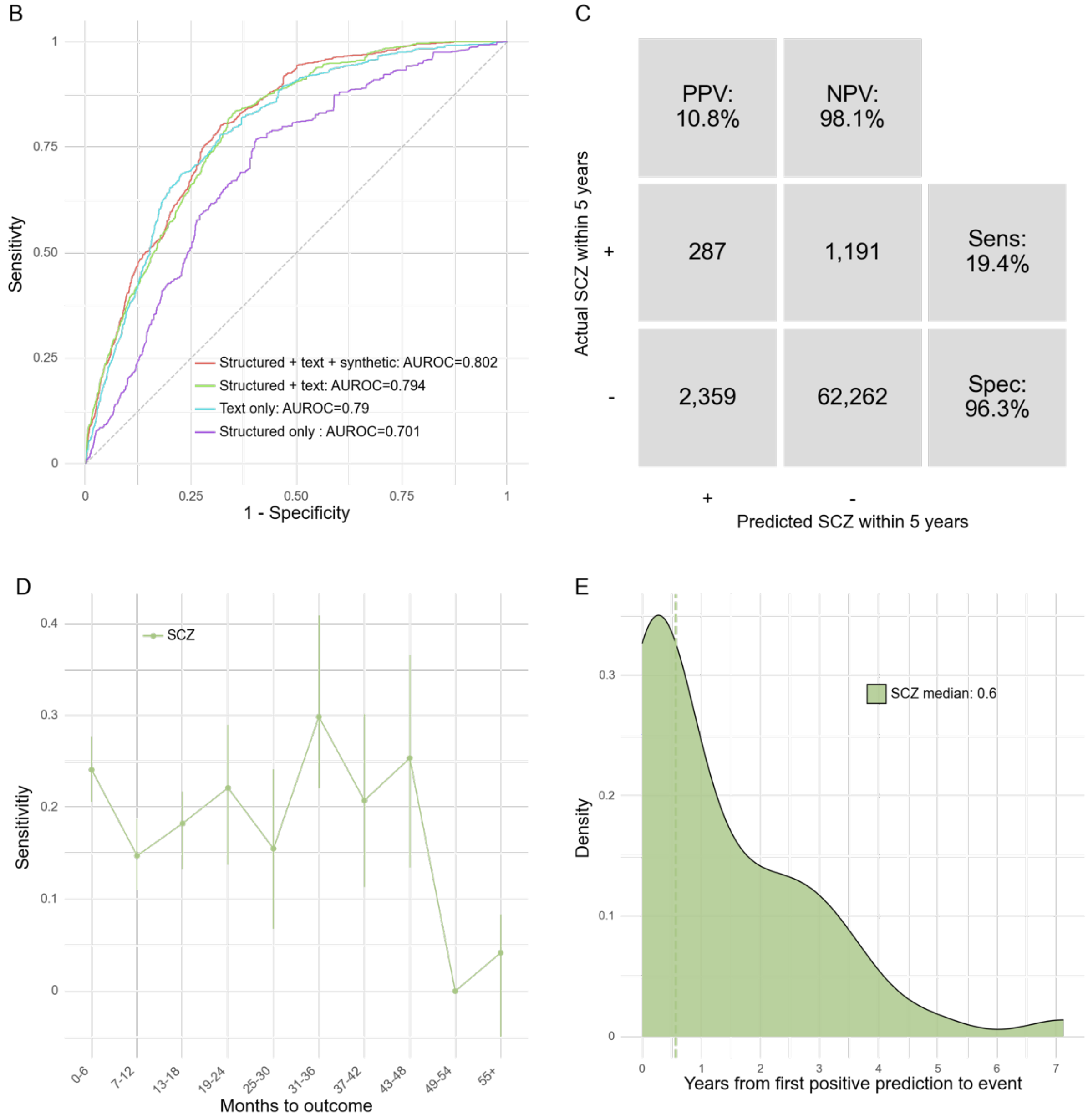

***Supplementary Figure 3**: Performance of the model trained and evaluated on the schizophrenia outcome.* *A) AUROC and median years to first positive prediction for the best model for each dataset on out-of-fold predictions on the training set. The best model was XGBoost in all cases.* ***B)*** *Receiver operating characteristics (ROC) curve of the best-performing models for each feature set on the test set. The model with the highest AUROC on the test (structured + text+ synthetic) was used in panels C-E with a classification threshold corresponding to 4% positives.* ***C)*** *Confusion matrix. PPV: Positive predictive value. NPV: Negative predictive value.* ***D)*** *Sensitivity by months from prediction time to event, stratified by outcome (BP=bipolar disorder, SCZ=schizophrenia).* ***E)*** *Time (years) from the first positive prediction to the patient receiving a diagnosis of bipolar disorder or schizophrenia. The dotted lines indicate the median time for each group.*

***Supplementary Table 8****: Performance by predicted positive rate for the best-performing schizophrenia model on the test set (structured + text + synthetic).*

| Predicted positive rate | True prevalence | PPV | NPV | Sens | Spec | FPR | FNR | Acc | TP | TN | FP | FN | F1 | MCC | % of all SCZ captured | Median years from first positive to first SCZ diagnosis |
|---|---|---|---|---|---|---|---|---|---|---|---|---|---|---|---|---|
| 8.0% | 2.2% | 8.7% | 98.3% | 31.1% | 92.5% | 7.5% | 68.9% | 91.2% | 460 | 59,793 | 4,828 | 1,018 | 13.6% | 12.9% | 30.4% | 0.9 |
| 6.0% | 2.2% | 9.3% | 98.2% | 24.8% | 94.4% | 5.6% | 75.2% | 92.9% | 367 | 61,022 | 3,599 | 1,111 | 13.5% | 12.0% | 23.2% | 0.6 |
| 4.0% | 2.2% | 10.8% | 98.1% | 19.4% | 96.3% | 3.7% | 80.6% | 94.6% | 287 | 62,262 | 2,359 | 1,191 | 13.9% | 11.9% | 16.0% | 0.6 |
| 2.0% | 2.2% | 11.8% | 98.0% | 10.6% | 98.2% | 1.8% | 89.4% | 96.2% | 156 | 63,455 | 1,166 | 1,322 | 11.1% | 9.2% | 11.2% | 0.6 |
| 1.0% | 2.2% | 16.3% | 97.9% | 7.3% | 99.1% | 0.9% | 92.7% | 97.1% | 108 | 64,068 | 553 | 1,370 | 10.1% | 9.6% | 8.0% | 0.8 |

**A. Results from model training: bipolar disorder model.**

| Median years from first positive prediction to outcome at predicted positive rate (PPR) | | | | | |
|---|---|---|---|---|---|
| **Dataset** / **PPR** | **0.01** | **0.02** | **0.04** | **0.06** | **0.08** |
| Structured + text + synthetic (AUROC = 0.66) | 0.3 | 0.3 | 0.5 | 0.6 | 0.6 |
| Structured + text (AUROC = 0.67) | 0.3 | 0.5 | 0.5 | 0.6 | 0.7 |
| Text only (AUROC = 0.68) | 0.3 | 0.3 | 0.5 | 0.7 | 0.7 |
| Structured only (AUROC = 0.63) | 1.4 | 2.6 | 1.5 | 1.0 | 1.1 |

**B-E. Results from model testing: bipolar disorder model**

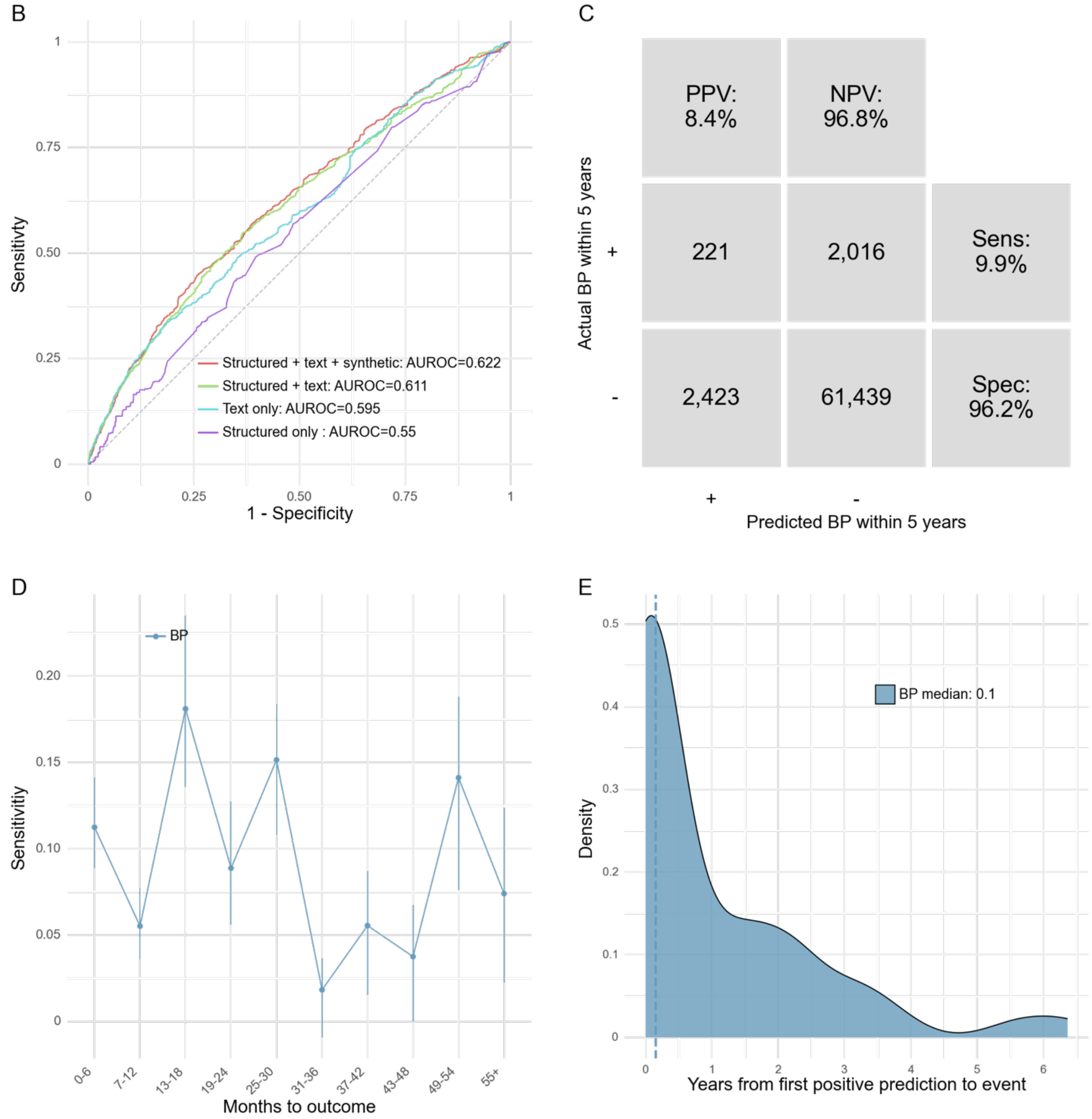


***Supplementary Figure 4**: Performance of the model trained and evaluated on the bipolar disorder outcome. **A)** AUROC and median years to first positive prediction for the best model for each dataset on out-of-fold predictions on the training set. The best model was XGBoost in all cases. **B)** Receiver operating characteristics (ROC) curve of the best-performing models for each feature set on the test set. The model with the highest AUROC on the*

*test (structured + text+ synthetic) was used in panels C-E with a classification threshold corresponding to 4% positives.* ***C)*** *Confusion matrix. PPV: Positive predictive value. NPV: Negative predictive value.* ***D)*** *Sensitivity by months from prediction time to event, stratified by outcome (BP=bipolar disorder, SCZ=schizophrenia).* ***E)*** *Time (years) from the first positive prediction to the patient receiving a diagnosis of bipolar disorder or schizophrenia. The dotted lines indicate the median time for each group.*

***Supplementary Table 9***: *Performance by predicted positive rate for the best-performing bipolar disorder model on the test set (structured + text + synthetic).*

| Predicted positive rate | True prevalence | PPV | NPV | Sens | Spec | FPR | FNR | Acc | TP | TN | FP | FN | F1 | MCC | % of all BP captured | Median years from first positive to first BP diagnosis |
|---|---|---|---|---|---|---|---|---|---|---|---|---|---|---|---|---|
| 8.0% | 3.4% | 7.6% | 97.0% | 18.0% | 92.3% | 7.7% | 82.0% | 89.8% | 402 | 58,972 | 4,890 | 1,835 | 10.7% | 6.9% | 18.5% | 0.4 |
| 6.0% | 3.4% | 7.6% | 96.9% | 13.5% | 94.3% | 5.7% | 86.5% | 91.5% | 301 | 60,196 | 3,666 | 1,936 | 9.7% | 5.9% | 14.2% | 0.4 |
| 4.0% | 3.4% | 8.4% | 96.8% | 9.9% | 96.2% | 3.8% | 90.1% | 93.3% | 221 | 61,439 | 2,423 | 2,016 | 9.1% | 5.6% | 9.9% | 0.1 |
| 2.0% | 3.4% | 9.4% | 96.7% | 5.5% | 98.1% | 1.9% | 94.5% | 95.0% | 124 | 62,664 | 1,198 | 2,113 | 7.0% | 4.7% | 7.3% | 0.1 |
| 1.0% | 3.4% | 9.6% | 96.7% | 2.9% | 99.0% | 1.0% | 97.1% | 95.8% | 65 | 63,248 | 614 | 2,172 | 4.5% | 3.5% | 5.2% | 0.0 |

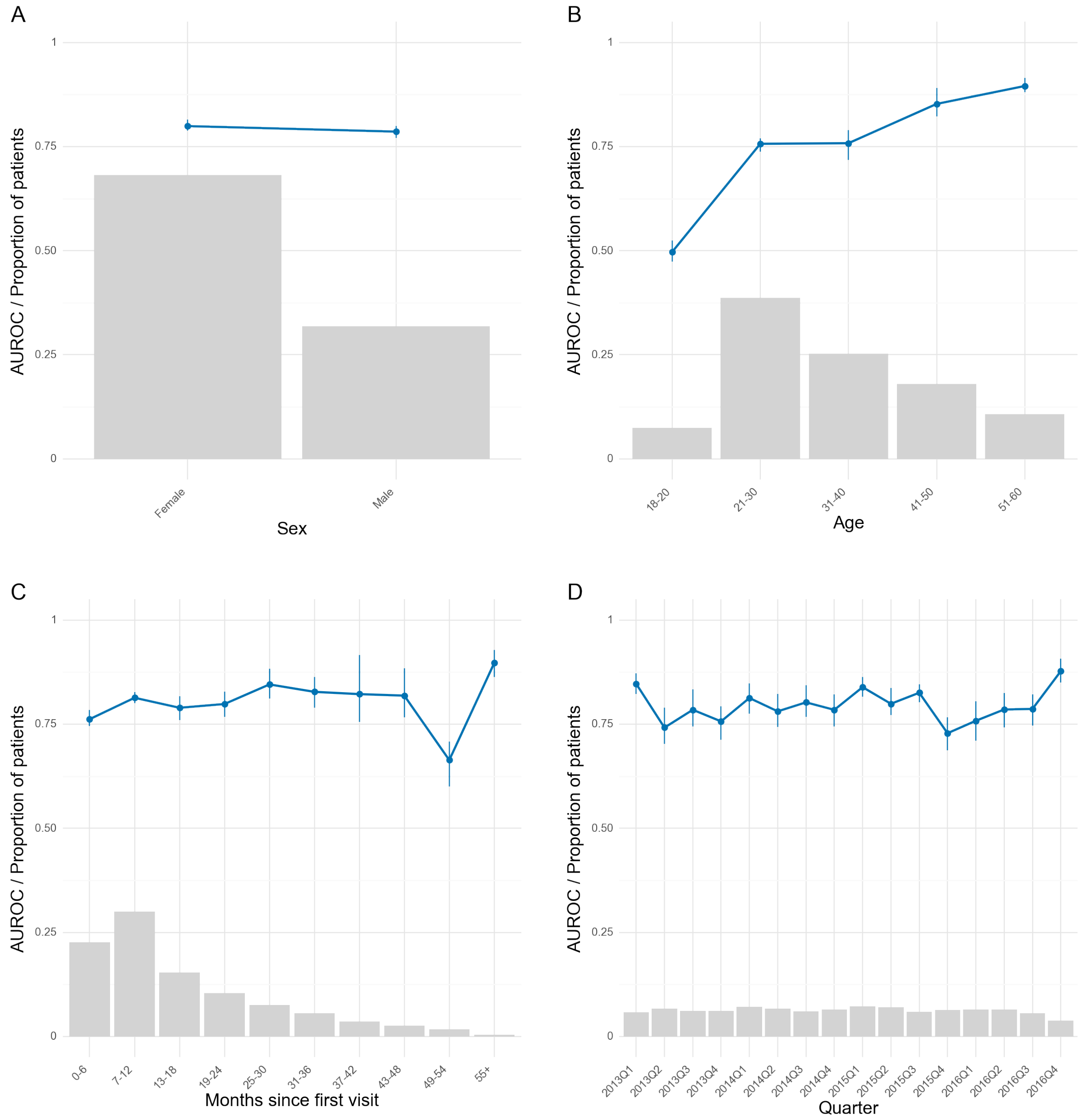


***Supplementary Figure 5****: Robustness of the best schizophrenia model (structured + text) across stratifications on the test set. Blue line is the area under the receiver operating characteristics curve. Grey bars represent the proportion of prediction times in each bin. Error bars are 95%-confidence intervals from 100-fold bootstrap. Due to the low n in some of the bins, some bootstrap folds contained only one class. This resulted in missing error bars for those bins.*

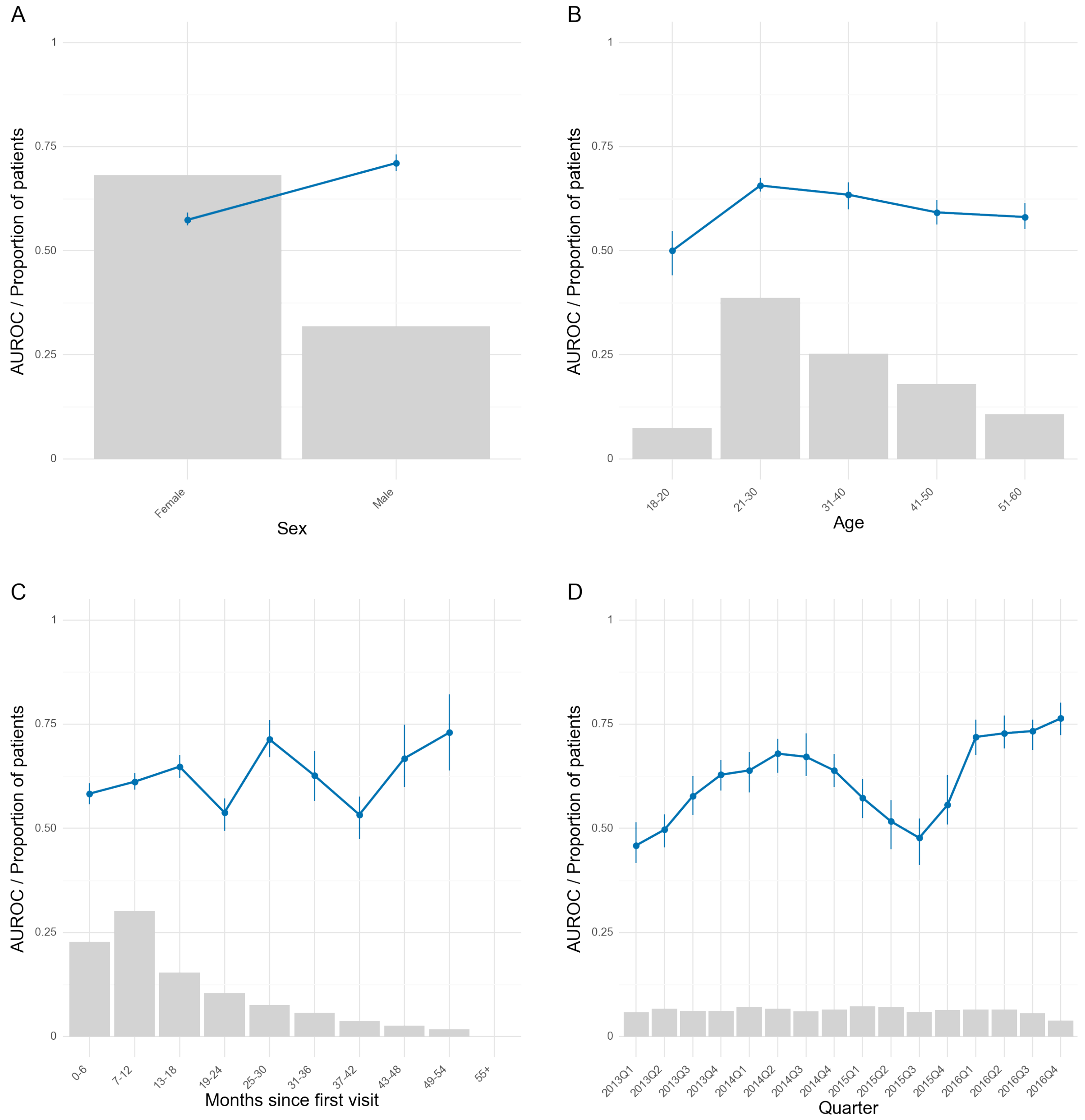


***Supplementary Figure 6**: Robustness of the best bipolar disorder model (structured + text) across stratifications on the test set. Blue line is the area under the receiver operating characteristics curve. Grey bars represent the proportion of prediction times in each bin. Error bars are 95%-confidence intervals from 100-fold bootstrap. Due to the low n in some of the bins, some bootstrap folds contained only one class. This resulted in missing error bars for those bins.*

***Supplementary Table 10**: Performance (AUROC) of models trained on the individual outcomes. Train denotes the mean and standard deviation of the out-of-fold predictions during 5-fold cross-validation on the training set. Test denotes AUROC on the test set.*

| Outcome | Feature set | Train | Test |
|---|---|---|---|
| Schizophrenia | Structured + text + synthetic | 0.774 ±0.02 | 0.80 |
| | Structured + text | 0.778 ±0.055 | 0.79 |
| | Text only | 0.774 ±0.064 | 0.79 |
| | Structured only | 0.725 ±0.053 | 0.70 |
| Bipolar disorder | Structured + text + synthetic | 0.658 ±0.038 | 0.62 |
| | Structured + text | 0.674 ±0.038 | 0.61 |
| | Text only | 0.669 ±0.047 | 0.59 |
| | Structured only | 0.632 ±0.039 | 0.55 |

***Supplementary Table 11***: *Results from model testing. AUROC of joint and individual models evaluated on each outcome on the test set.*

| Model \ Outcome | Schizophrenia | First schizophrenia or bipolar disorder diagnosis | Bipolar disorder |
|---|---|---|---|
| Schizophrenia only | 0.80 | - | - |
| Joint model | 0.74 | 0.64 | 0.57 |
| Bipolar disorder only | - | - | 0.62 |

***Supplementary Table 1:*** *Predictor description. Lookbehind period refers to how far back in time from the prediction time to look for values, resolve multiple indicates the aggregation function, i.e. how to handle multiple values in the lookbehind, fallback strategy indicates which value set if no values exists in the lookbehind for the predictor, static indicates whether the value can change over time or is static (e.g. sex), mean indicates the mean value, N. unique represents the number of unique values, and proportion using fallback refers to how large a proportion of the values in the column consists of the fallback value.*

### Predictors descriptive stats

| | Feature name | Lookbehind period | Resolve multiple | Fallback strategy | Static | Mean | N. unique | Proportion using fallback |
|---|---|---|---|---|---|---|---|---|
| **0** | admissions | 0 to 183 days | count | 0 | False | 0.20 | 32 | 0.90 |
| **1** | admissions | 0 to 365 days | count | 0 | False | 0.34 | 53 | 0.85 |
| **2** | admissions | 0 to 730 days | count | 0 | False | 0.54 | 81 | 0.81 |
| **3** | age_years | 0 | N/A | nan | False | 33.80 | 4501 | 0.00 |
| **4** | tfidf-1000_10 | 0 to 730 days | mean | nan | False | 0.01 | 126649 | 0.00 |
| **5** | tfidf-1000_100 | 0 to 730 days | mean | nan | False | 0.00 | 47122 | 0.00 |
| **6** | tfidf-1000_1000 | 0 to 730 days | mean | nan | False | 0.00 | 50965 | 0.00 |
| **7** | tfidf-1000_11 | 0 to 730 days | mean | nan | False | 0.00 | 71421 | 0.00 |
| **8** | tfidf-1000_1100 | 0 to 730 days | mean | nan | False | 0.00 | 48927 | 0.00 |
| **9** | tfidf-1000_12 | 0 to 730 days | mean | nan | False | 0.00 | 103731 | 0.00 |
| **10** | tfidf-1000_13 | 0 to 730 days | mean | nan | False | 0.00 | 75289 | 0.00 |
| **11** | tfidf-1000_1300 | 0 to 730 days | mean | nan | False | 0.00 | 52393 | 0.00 |
| **12** | tfidf-1000_14 | 0 to 730 days | mean | nan | False | 0.01 | 113788 | 0.00 |

| | Feature name | Lookbehind period | Resolve multiple | Fallback strategy | Static | Mean | N. unique | Proportion using fallback |
|---|---|---|---|---|---|---|---|---|
| **13** | tfidf-1000_14 dage | 0 to 730 days | mean | nan | False | 0.00 | 68634 | 0.00 |
| **14** | tfidf-1000_15 | 0 to 730 days | mean | nan | False | 0.00 | 91006 | 0.00 |
| **15** | tfidf-1000_16 | 0 to 730 days | mean | nan | False | 0.00 | 56219 | 0.00 |
| **16** | tfidf-1000_17 | 0 to 730 days | mean | nan | False | 0.00 | 46617 | 0.00 |
| **17** | tfidf-1000_18 | 0 to 730 days | mean | nan | False | 0.00 | 53780 | 0.00 |
| **18** | tfidf-1000_20 | 0 to 730 days | mean | nan | False | 0.00 | 79766 | 0.00 |
| **19** | tfidf-1000_22 | 0 to 730 days | mean | nan | False | 0.00 | 39264 | 0.00 |
| **20** | tfidf-1000_23 | 0 to 730 days | mean | nan | False | 0.00 | 78596 | 0.00 |
| **21** | tfidf-1000_25 | 0 to 730 days | mean | nan | False | 0.00 | 63369 | 0.00 |
| **22** | tfidf-1000_30 | 0 to 730 days | mean | nan | False | 0.00 | 60772 | 0.00 |
| **23** | tfidf-1000_34 | 0 to 730 days | mean | nan | False | 0.00 | 59701 | 0.00 |
| **24** | tfidf-1000_50 | 0 to 730 days | mean | nan | False | 0.00 | 49557 | 0.00 |
| **25** | tfidf-1000_abilify | 0 to 730 days | mean | nan | False | 0.00 | 9031 | 0.00 |
| **26** | tfidf-1000_acceptere | 0 to 730 days | mean | nan | False | 0.00 | 44536 | 0.00 |
| **27** | tfidf-1000_accepterer | 0 to 730 days | mean | nan | False | 0.00 | 48019 | 0.00 |
| **28** | tfidf-1000_adfærd | 0 to 730 days | mean | nan | False | 0.00 | 85414 | 0.00 |
| **29** | tfidf-1000_adhd | 0 to 730 days | mean | nan | False | 0.01 | 58371 | 0.00 |
| **30** | tfidf-1000_adspurgt | 0 to 730 days | mean | nan | False | 0.00 | 98493 | 0.00 |

| | Feature name | Lookbehind period | Resolve multiple | Fallback strategy | Static | Mean | N. unique | Proportion using fallback |
|---|---|---|---|---|---|---|---|---|
| **31** | tfidf-1000_afd | 0 to 730 days | mean | nan | False | 0.00 | 53701 | 0.00 |
| **32** | tfidf-1000_afdeling | 0 to 730 days | mean | nan | False | 0.00 | 35315 | 0.00 |
| **33** | tfidf-1000_afdelingen | 0 to 730 days | mean | nan | False | 0.00 | 41741 | 0.00 |
| **34** | tfidf-1000_afdæmpet | 0 to 730 days | mean | nan | False | 0.00 | 14361 | 0.00 |
| **35** | tfidf-1000_aflede | 0 to 730 days | mean | nan | False | 0.00 | 41455 | 0.00 |
| **36** | tfidf-1000_afslutning | 0 to 730 days | mean | nan | False | 0.00 | 45414 | 0.00 |
| **37** | tfidf-1000_afsluttes | 0 to 730 days | mean | nan | False | 0.00 | 57428 | 0.00 |
| **38** | tfidf-1000_afsluttet | 0 to 730 days | mean | nan | False | 0.00 | 56217 | 0.00 |
| **39** | tfidf-1000_afsnittet | 0 to 730 days | mean | nan | False | 0.00 | 15051 | 0.00 |
| **40** | tfidf-1000_afstand | 0 to 730 days | mean | nan | False | 0.00 | 71552 | 0.00 |
| **41** | tfidf-1000_afsted | 0 to 730 days | mean | nan | False | 0.00 | 72436 | 0.00 |
| **42** | tfidf-1000_aftale | 0 to 730 days | mean | nan | False | 0.01 | 133899 | 0.00 |
| **43** | tfidf-1000_aftaler | 0 to 730 days | mean | nan | False | 0.01 | 150468 | 0.00 |
| **44** | tfidf-1000_aftales | 0 to 730 days | mean | nan | False | 0.01 | 126236 | 0.00 |
| **45** | tfidf-1000_aftales pt | 0 to 730 days | mean | nan | False | 0.00 | 51074 | 0.00 |
| **46** | tfidf-1000_aftalt | 0 to 730 days | mean | nan | False | 0.01 | 134593 | 0.00 |
| **47** | tfidf-1000_aftalt pt | 0 to 730 days | mean | nan | False | 0.00 | 41088 | 0.00 |
| **48** | tfidf-1000_aften | 0 to 730 days | mean | nan | False | 0.00 | 76265 | 0.00 |

| | Feature name | Lookbehind period | Resolve multiple | Fallback strategy | Static | Mean | N. unique | Proportion using fallback |
|---|---|---|---|---|---|---|---|---|
| **49** | tfidf-1000_aftenen | 0 to 730 days | mean | nan | False | 0.00 | 86811 | 0.00 |
| **50** | tfidf-1000_aftensmad | 0 to 730 days | mean | nan | False | 0.00 | 38916 | 0.00 |
| **51** | tfidf-1000_aftensmaden | 0 to 730 days | mean | nan | False | 0.00 | 29484 | 0.00 |
| **52** | tfidf-1000_afventer | 0 to 730 days | mean | nan | False | 0.00 | 55095 | 0.00 |
| **53** | tfidf-1000_afvisende | 0 to 730 days | mean | nan | False | 0.00 | 41265 | 0.00 |
| **54** | tfidf-1000_afviser | 0 to 730 days | mean | nan | False | 0.00 | 62495 | 0.00 |
| **55** | tfidf-1000_aktiv | 0 to 730 days | mean | nan | False | 0.00 | 55159 | 0.00 |
| **56** | tfidf-1000_aktivitet | 0 to 730 days | mean | nan | False | 0.00 | 54067 | 0.00 |
| **57** | tfidf-1000_aktiviteter | 0 to 730 days | mean | nan | False | 0.00 | 89064 | 0.00 |
| **58** | tfidf-1000_aktuelle | 0 to 730 days | mean | nan | False | 0.00 | 100656 | 0.00 |
| **59** | tfidf-1000_aktuelt | 0 to 730 days | mean | nan | False | 0.01 | 155736 | 0.00 |
| **60** | tfidf-1000_akut | 0 to 730 days | mean | nan | False | 0.00 | 60489 | 0.00 |
| **61** | tfidf-1000_alkohol | 0 to 730 days | mean | nan | False | 0.00 | 53266 | 0.00 |
| **62** | tfidf-1000_amb | 0 to 730 days | mean | nan | False | 0.00 | 45851 | 0.00 |
| **63** | tfidf-1000_ambulant | 0 to 730 days | mean | nan | False | 0.00 | 46213 | 0.00 |
| **64** | tfidf-1000_anderledes | 0 to 730 days | mean | nan | False | 0.00 | 57233 | 0.00 |
| **65** | tfidf-1000_ang | 0 to 730 days | mean | nan | False | 0.00 | 66706 | 0.00 |
| **66** | tfidf-1000_angiveligt | 0 to 730 days | mean | nan | False | 0.00 | 74624 | 0.00 |

| | Feature name | Lookbehind period | Resolve multiple | Fallback strategy | Static | Mean | N. unique | Proportion using fallback |
|---|---|---|---|---|---|---|---|---|
| **67** | tfidf-1000_angiver | 0 to 730 days | mean | nan | False | 0.01 | 132562 | 0.00 |
| **68** | tfidf-1000_angst | 0 to 730 days | mean | nan | False | 0.01 | 154696 | 0.00 |
| **69** | tfidf-1000_angstanfald | 0 to 730 days | mean | nan | False | 0.00 | 46303 | 0.00 |
| **70** | tfidf-1000_angsten | 0 to 730 days | mean | nan | False | 0.00 | 64596 | 0.00 |
| **71** | tfidf-1000_angstsymptomer | 0 to 730 days | mean | nan | False | 0.00 | 58820 | 0.00 |
| **72** | tfidf-1000_anne | 0 to 730 days | mean | nan | False | 0.00 | 23840 | 0.00 |
| **73** | tfidf-1000_appetit | 0 to 730 days | mean | nan | False | 0.00 | 73703 | 0.00 |
| **74** | tfidf-1000_apsykotisk | 0 to 730 days | mean | nan | False | 0.00 | 56187 | 0.00 |
| **75** | tfidf-1000_arbejde | 0 to 730 days | mean | nan | False | 0.01 | 158216 | 0.00 |
| **76** | tfidf-1000_arbejder | 0 to 730 days | mean | nan | False | 0.00 | 97048 | 0.00 |
| **77** | tfidf-1000_arbejdet | 0 to 730 days | mean | nan | False | 0.00 | 86560 | 0.00 |
| **78** | tfidf-1000_av | 0 to 730 days | mean | nan | False | 0.00 | 18849 | 0.00 |
| **79** | tfidf-1000_bad | 0 to 730 days | mean | nan | False | 0.00 | 56879 | 0.00 |
| **80** | tfidf-1000_baggrund | 0 to 730 days | mean | nan | False | 0.00 | 76783 | 0.00 |
| **81** | tfidf-1000_bange | 0 to 730 days | mean | nan | False | 0.01 | 140476 | 0.00 |
| **82** | tfidf-1000_barn | 0 to 730 days | mean | nan | False | 0.00 | 72580 | 0.00 |
| **83** | tfidf-1000_beder | 0 to 730 days | mean | nan | False | 0.00 | 63538 | 0.00 |
| **84** | tfidf-1000_bedre | 0 to 730 days | mean | nan | False | 0.01 | 171103 | 0.00 |

| | Feature name | Lookbehind period | Resolve multiple | Fallback strategy | Static | Mean | N. unique | Proportion using fallback |
|---|---|---|---|---|---|---|---|---|
| **85** | tfidf-1000_bedring | 0 to 730 days | mean | nan | False | 0.00 | 83140 | 0.00 |
| **86** | tfidf-1000_bedst | 0 to 730 days | mean | nan | False | 0.00 | 77190 | 0.00 |
| **87** | tfidf-1000_bedt | 0 to 730 days | mean | nan | False | 0.00 | 56009 | 0.00 |
| **88** | tfidf-1000_begynder | 0 to 730 days | mean | nan | False | 0.00 | 76530 | 0.00 |
| **89** | tfidf-1000_begyndt | 0 to 730 days | mean | nan | False | 0.00 | 104650 | 0.00 |
| **90** | tfidf-1000_beh | 0 to 730 days | mean | nan | False | 0.00 | 53993 | 0.00 |
| **91** | tfidf-1000_behandler | 0 to 730 days | mean | nan | False | 0.00 | 66353 | 0.00 |
| **92** | tfidf-1000_behandling | 0 to 730 days | mean | nan | False | 0.02 | 180644 | 0.00 |
| **93** | tfidf-1000_behandlingen | 0 to 730 days | mean | nan | False | 0.00 | 86540 | 0.00 |
| **94** | tfidf-1000_behov | 0 to 730 days | mean | nan | False | 0.01 | 137322 | 0.00 |
| **95** | tfidf-1000_bekræfter | 0 to 730 days | mean | nan | False | 0.00 | 57954 | 0.00 |
| **96** | tfidf-1000_bekymret | 0 to 730 days | mean | nan | False | 0.00 | 108282 | 0.00 |
| **97** | tfidf-1000_bekymring | 0 to 730 days | mean | nan | False | 0.00 | 60050 | 0.00 |
| **98** | tfidf-1000_bekymringer | 0 to 730 days | mean | nan | False | 0.00 | 62132 | 0.00 |
| **99** | tfidf-1000_belastet | 0 to 730 days | mean | nan | False | 0.00 | 59668 | 0.00 |
| **100** | tfidf-1000_ben | 0 to 730 days | mean | nan | False | 0.00 | 45612 | 0.00 |
| **101** | tfidf-1000_benægter | 0 to 730 days | mean | nan | False | 0.00 | 58525 | 0.00 |
| **102** | tfidf-1000_besked | 0 to 730 days | mean | nan | False | 0.00 | 82208 | 0.00 |

| | Feature name | Lookbehind period | Resolve multiple | Fallback strategy | Static | Mean | N. unique | Proportion using fallback |
|---|---|---|---|---|---|---|---|---|
| **103** | tfidf-1000_beskrevet | 0 to 730 days | mean | nan | False | 0.00 | 60945 | 0.00 |
| **104** | tfidf-1000_beskrive | 0 to 730 days | mean | nan | False | 0.00 | 60991 | 0.00 |
| **105** | tfidf-1000_beskriver | 0 to 730 days | mean | nan | False | 0.01 | 172763 | 0.00 |
| **106** | tfidf-1000_beskrives | 0 to 730 days | mean | nan | False | 0.00 | 54387 | 0.00 |
| **107** | tfidf-1000_bestemt | 0 to 730 days | mean | nan | False | 0.00 | 51576 | 0.00 |
| **108** | tfidf-1000_bestilt | 0 to 730 days | mean | nan | False | 0.00 | 38241 | 0.00 |
| **109** | tfidf-1000_besøg | 0 to 730 days | mean | nan | False | 0.00 | 94441 | 0.00 |
| **110** | tfidf-1000_besøge | 0 to 730 days | mean | nan | False | 0.00 | 41257 | 0.00 |
| **111** | tfidf-1000_bil | 0 to 730 days | mean | nan | False | 0.00 | 50202 | 0.00 |
| **112** | tfidf-1000_bipolar | 0 to 730 days | mean | nan | False | 0.00 | 14056 | 0.00 |
| **113** | tfidf-1000_bivirkninger | 0 to 730 days | mean | nan | False | 0.00 | 67186 | 0.00 |
| **114** | tfidf-1000_bla | 0 to 730 days | mean | nan | False | 0.00 | 99821 | 0.00 |
| **115** | tfidf-1000_blevet | 0 to 730 days | mean | nan | False | 0.01 | 170001 | 0.00 |
| **116** | tfidf-1000_blodprøver | 0 to 730 days | mean | nan | False | 0.00 | 61225 | 0.00 |
| **117** | tfidf-1000_bo | 0 to 730 days | mean | nan | False | 0.00 | 56128 | 0.00 |
| **118** | tfidf-1000_bolig | 0 to 730 days | mean | nan | False | 0.00 | 37363 | 0.00 |
| **119** | tfidf-1000_bor | 0 to 730 days | mean | nan | False | 0.01 | 160961 | 0.00 |
| **120** | tfidf-1000_bosted | 0 to 730 days | mean | nan | False | 0.00 | 13678 | 0.00 |

| | Feature name | Lookbehind period | Resolve multiple | Fallback strategy | Static | Mean | N. unique | Proportion using fallback |
|---|---|---|---|---|---|---|---|---|
| **121** | tfidf-1000_bostedet | 0 to 730 days | mean | nan | False | 0.00 | 7682 | 0.00 |
| **122** | tfidf-1000_bostøtte | 0 to 730 days | mean | nan | False | 0.00 | 43701 | 0.00 |
| **123** | tfidf-1000_brev | 0 to 730 days | mean | nan | False | 0.00 | 63306 | 0.00 |
| **124** | tfidf-1000_bror | 0 to 730 days | mean | nan | False | 0.00 | 43866 | 0.00 |
| **125** | tfidf-1000_bruge | 0 to 730 days | mean | nan | False | 0.00 | 94516 | 0.00 |
| **126** | tfidf-1000_bruger | 0 to 730 days | mean | nan | False | 0.00 | 109072 | 0.00 |
| **127** | tfidf-1000_brugt | 0 to 730 days | mean | nan | False | 0.00 | 60754 | 0.00 |
| **128** | tfidf-1000_byen | 0 to 730 days | mean | nan | False | 0.00 | 54822 | 0.00 |
| **129** | tfidf-1000_børn | 0 to 730 days | mean | nan | False | 0.01 | 101659 | 0.00 |
| **130** | tfidf-1000_børnene | 0 to 730 days | mean | nan | False | 0.00 | 56195 | 0.00 |
| **131** | tfidf-1000_cigaretter | 0 to 730 days | mean | nan | False | 0.00 | 13992 | 0.00 |
| **132** | tfidf-1000_computer | 0 to 730 days | mean | nan | False | 0.00 | 32551 | 0.00 |
| **133** | tfidf-1000_dage | 0 to 730 days | mean | nan | False | 0.01 | 162307 | 0.00 |
| **134** | tfidf-1000_dagen | 0 to 730 days | mean | nan | False | 0.00 | 134214 | 0.00 |
| **135** | tfidf-1000_dagens | 0 to 730 days | mean | nan | False | 0.00 | 83615 | 0.00 |
| **136** | tfidf-1000_dagens samtale | 0 to 730 days | mean | nan | False | 0.00 | 64111 | 0.00 |
| **137** | tfidf-1000_daglige | 0 to 730 days | mean | nan | False | 0.00 | 69772 | 0.00 |
| **138** | tfidf-1000_dagligt | 0 to 730 days | mean | nan | False | 0.00 | 106178 | 0.00 |

| | Feature name | Lookbehind period | Resolve multiple | Fallback strategy | Static | Mean | N. unique | Proportion using fallback |
|---|---|---|---|---|---|---|---|---|
| **139** | tfidf-1000_data | 0 to 730 days | mean | nan | False | 0.00 | 46041 | 0.00 |
| **140** | tfidf-1000_datter | 0 to 730 days | mean | nan | False | 0.00 | 54754 | 0.00 |
| **141** | tfidf-1000_datteren | 0 to 730 days | mean | nan | False | 0.00 | 37513 | 0.00 |
| **142** | tfidf-1000_dd | 0 to 730 days | mean | nan | False | 0.01 | 97201 | 0.00 |
| **143** | tfidf-1000_dels | 0 to 730 days | mean | nan | False | 0.00 | 62589 | 0.00 |
| **144** | tfidf-1000_deltage | 0 to 730 days | mean | nan | False | 0.00 | 87043 | 0.00 |
| **145** | tfidf-1000_deltagelse | 0 to 730 days | mean | nan | False | 0.00 | 61129 | 0.00 |
| **146** | tfidf-1000_deltager | 0 to 730 days | mean | nan | False | 0.01 | 100589 | 0.00 |
| **147** | tfidf-1000_deltaget | 0 to 730 days | mean | nan | False | 0.00 | 38889 | 0.00 |
| **148** | tfidf-1000_depression | 0 to 730 days | mean | nan | False | 0.01 | 127556 | 0.00 |
| **149** | tfidf-1000_depressiv | 0 to 730 days | mean | nan | False | 0.00 | 94669 | 0.00 |
| **150** | tfidf-1000_depressive | 0 to 730 days | mean | nan | False | 0.01 | 119615 | 0.00 |
| **151** | tfidf-1000_depressive symptomer | 0 to 730 days | mean | nan | False | 0.00 | 92731 | 0.00 |
| **152** | tfidf-1000_deprimeret | 0 to 730 days | mean | nan | False | 0.00 | 72093 | 0.00 |
| **153** | tfidf-1000_derhjemme | 0 to 730 days | mean | nan | False | 0.00 | 88658 | 0.00 |
| **154** | tfidf-1000_derudover | 0 to 730 days | mean | nan | False | 0.00 | 73518 | 0.00 |
| **155** | tfidf-1000_desuden | 0 to 730 days | mean | nan | False | 0.00 | 113989 | 0.00 |
| **156** | tfidf-1000_dgl | 0 to 730 days | mean | nan | False | 0.00 | 44117 | 0.00 |

| | Feature name | Lookbehind period | Resolve multiple | Fallback strategy | Static | Mean | N. unique | Proportion using fallback |
|---|---|---|---|---|---|---|---|---|
| **157** | tfidf-1000_diagnose | 0 to 730 days | mean | nan | False | 0.00 | 55726 | 0.00 |
| **158** | tfidf-1000_diagnosen | 0 to 730 days | mean | nan | False | 0.00 | 60567 | 0.00 |
| **159** | tfidf-1000_direkte | 0 to 730 days | mean | nan | False | 0.00 | 56833 | 0.00 |
| **160** | tfidf-1000_dosis | 0 to 730 days | mean | nan | False | 0.00 | 58736 | 0.00 |
| **161** | tfidf-1000_drikke | 0 to 730 days | mean | nan | False | 0.00 | 46759 | 0.00 |
| **162** | tfidf-1000_drikker | 0 to 730 days | mean | nan | False | 0.00 | 42512 | 0.00 |
| **163** | tfidf-1000_drukket | 0 to 730 days | mean | nan | False | 0.00 | 33468 | 0.00 |
| **164** | tfidf-1000_drøfter | 0 to 730 days | mean | nan | False | 0.00 | 57215 | 0.00 |
| **165** | tfidf-1000_drøftes | 0 to 730 days | mean | nan | False | 0.00 | 56619 | 0.00 |
| **166** | tfidf-1000_dv | 0 to 730 days | mean | nan | False | 0.00 | 16929 | 0.00 |
| **167** | tfidf-1000_dårlig | 0 to 730 days | mean | nan | False | 0.00 | 131722 | 0.00 |
| **168** | tfidf-1000_dårlige | 0 to 730 days | mean | nan | False | 0.00 | 73676 | 0.00 |
| **169** | tfidf-1000_dårligt | 0 to 730 days | mean | nan | False | 0.01 | 140599 | 0.00 |
| **170** | tfidf-1000_dø | 0 to 730 days | mean | nan | False | 0.00 | 47774 | 0.00 |
| **171** | tfidf-1000_død | 0 to 730 days | mean | nan | False | 0.00 | 54210 | 0.00 |
| **172** | tfidf-1000_døren | 0 to 730 days | mean | nan | False | 0.00 | 53438 | 0.00 |
| **173** | tfidf-1000_ect | 0 to 730 days | mean | nan | False | 0.00 | 15546 | 0.00 |
| **174** | tfidf-1000_effekt | 0 to 730 days | mean | nan | False | 0.01 | 120782 | 0.00 |

| | Feature name | Lookbehind period | Resolve multiple | Fallback strategy | Static | Mean | N. unique | Proportion using fallback |
|---|---|---|---|---|---|---|---|---|
| **175** | tfidf-1000_efterfølgende | 0 to 730 days | mean | nan | False | 0.00 | 121099 | 0.00 |
| **176** | tfidf-1000_eftermiddag | 0 to 730 days | mean | nan | False | 0.00 | 43146 | 0.00 |
| **177** | tfidf-1000_eftermiddagen | 0 to 730 days | mean | nan | False | 0.00 | 41132 | 0.00 |
| **178** | tfidf-1000_efterspørger | 0 to 730 days | mean | nan | False | 0.00 | 27780 | 0.00 |
| **179** | tfidf-1000_egentlig | 0 to 730 days | mean | nan | False | 0.00 | 94771 | 0.00 |
| **180** | tfidf-1000_eget | 0 to 730 days | mean | nan | False | 0.00 | 88860 | 0.00 |
| **181** | tfidf-1000_egne | 0 to 730 days | mean | nan | False | 0.00 | 103913 | 0.00 |
| **182** | tfidf-1000_ekg | 0 to 730 days | mean | nan | False | 0.00 | 48393 | 0.00 |
| **183** | tfidf-1000_eks | 0 to 730 days | mean | nan | False | 0.00 | 44187 | 0.00 |
| **184** | tfidf-1000_ekstra | 0 to 730 days | mean | nan | False | 0.00 | 53086 | 0.00 |
| **185** | tfidf-1000_el | 0 to 730 days | mean | nan | False | 0.01 | 87673 | 0.00 |
| **186** | tfidf-1000_emotionel | 0 to 730 days | mean | nan | False | 0.01 | 132209 | 0.00 |
| **187** | tfidf-1000_emotionel kontakt | 0 to 730 days | mean | nan | False | 0.01 | 117394 | 0.00 |
| **188** | tfidf-1000_emotionelt | 0 to 730 days | mean | nan | False | 0.00 | 54000 | 0.00 |
| **189** | tfidf-1000_endvidere | 0 to 730 days | mean | nan | False | 0.00 | 88870 | 0.00 |
| **190** | tfidf-1000_energi | 0 to 730 days | mean | nan | False | 0.01 | 134645 | 0.00 |
| **191** | tfidf-1000_enkelt | 0 to 730 days | mean | nan | False | 0.00 | 70783 | 0.00 |
| **192** | tfidf-1000_enkelte | 0 to 730 days | mean | nan | False | 0.00 | 63537 | 0.00 |

| | Feature name | Lookbehind period | Resolve multiple | Fallback strategy | Static | Mean | N. unique | Proportion using fallback |
|---|---|---|---|---|---|---|---|---|
| **193** | tfidf-1000_episode | 0 to 730 days | mean | nan | False | 0.00 | 55831 | 0.00 |
| **194** | tfidf-1000_episoder | 0 to 730 days | mean | nan | False | 0.00 | 62266 | 0.00 |
| **195** | tfidf-1000_evt | 0 to 730 days | mean | nan | False | 0.01 | 141889 | 0.00 |
| **196** | tfidf-1000_faderen | 0 to 730 days | mean | nan | False | 0.00 | 55624 | 0.00 |
| **197** | tfidf-1000_fald | 0 to 730 days | mean | nan | False | 0.00 | 51913 | 0.00 |
| **198** | tfidf-1000_falde | 0 to 730 days | mean | nan | False | 0.00 | 93098 | 0.00 |
| **199** | tfidf-1000_falde søvn | 0 to 730 days | mean | nan | False | 0.00 | 71477 | 0.00 |
| **200** | tfidf-1000_falder | 0 to 730 days | mean | nan | False | 0.00 | 94324 | 0.00 |
| **201** | tfidf-1000_faldet | 0 to 730 days | mean | nan | False | 0.00 | 44033 | 0.00 |
| **202** | tfidf-1000_familie | 0 to 730 days | mean | nan | False | 0.00 | 110541 | 0.00 |
| **203** | tfidf-1000_familien | 0 to 730 days | mean | nan | False | 0.01 | 115073 | 0.00 |
| **204** | tfidf-1000_fast | 0 to 730 days | mean | nan | False | 0.00 | 96430 | 0.00 |
| **205** | tfidf-1000_faste | 0 to 730 days | mean | nan | False | 0.00 | 46744 | 0.00 |
| **206** | tfidf-1000_feks | 0 to 730 days | mean | nan | False | 0.00 | 94278 | 0.00 |
| **207** | tfidf-1000_ferie | 0 to 730 days | mean | nan | False | 0.00 | 78877 | 0.00 |
| **208** | tfidf-1000_film | 0 to 730 days | mean | nan | False | 0.00 | 55615 | 0.00 |
| **209** | tfidf-1000_fin | 0 to 730 days | mean | nan | False | 0.00 | 42130 | 0.00 |
| **210** | tfidf-1000_finder | 0 to 730 days | mean | nan | False | 0.00 | 91344 | 0.00 |

| | Feature name | Lookbehind period | Resolve multiple | Fallback strategy | Static | Mean | N. unique | Proportion using fallback |
|---|---|---|---|---|---|---|---|---|
| **211** | tfidf-1000_findes | 0 to 730 days | mean | nan | False | 0.00 | 52234 | 0.00 |
| **212** | tfidf-1000_fint | 0 to 730 days | mean | nan | False | 0.00 | 89927 | 0.00 |
| **213** | tfidf-1000_flytte | 0 to 730 days | mean | nan | False | 0.00 | 75497 | 0.00 |
| **214** | tfidf-1000_flyttet | 0 to 730 days | mean | nan | False | 0.00 | 73769 | 0.00 |
| **215** | tfidf-1000_fokus | 0 to 730 days | mean | nan | False | 0.01 | 104113 | 0.00 |
| **216** | tfidf-1000_folk | 0 to 730 days | mean | nan | False | 0.00 | 73557 | 0.00 |
| **217** | tfidf-1000_forbindelse | 0 to 730 days | mean | nan | False | 0.01 | 136954 | 0.00 |
| **218** | tfidf-1000_forhold | 0 to 730 days | mean | nan | False | 0.01 | 163035 | 0.00 |
| **219** | tfidf-1000_forklare | 0 to 730 days | mean | nan | False | 0.00 | 54864 | 0.00 |
| **220** | tfidf-1000_forklarer | 0 to 730 days | mean | nan | False | 0.00 | 63856 | 0.00 |
| **221** | tfidf-1000_forløb | 0 to 730 days | mean | nan | False | 0.01 | 131807 | 0.00 |
| **222** | tfidf-1000_forløbet | 0 to 730 days | mean | nan | False | 0.00 | 85824 | 0.00 |
| **223** | tfidf-1000_form | 0 to 730 days | mean | nan | False | 0.00 | 128638 | 0.00 |
| **224** | tfidf-1000_formel | 0 to 730 days | mean | nan | False | 0.01 | 135214 | 0.00 |
| **225** | tfidf-1000_formel emotionel | 0 to 730 days | mean | nan | False | 0.01 | 99041 | 0.00 |
| **226** | tfidf-1000_formel kontakt | 0 to 730 days | mean | nan | False | 0.00 | 68309 | 0.00 |
| **227** | tfidf-1000_formentlig | 0 to 730 days | mean | nan | False | 0.00 | 58059 | 0.00 |
| **228** | tfidf-1000_formiddag | 0 to 730 days | mean | nan | False | 0.00 | 35806 | 0.00 |

| | Feature name | Lookbehind period | Resolve multiple | Fallback strategy | Static | Mean | N. unique | Proportion using fallback |
|---|---|---|---|---|---|---|---|---|
| **229** | tfidf-1000_formiddagen | 0 to 730 days | mean | nan | False | 0.00 | 37407 | 0.00 |
| **230** | tfidf-1000_formår | 0 to 730 days | mean | nan | False | 0.00 | 52073 | 0.00 |
| **231** | tfidf-1000_fornemmelse | 0 to 730 days | mean | nan | False | 0.00 | 66144 | 0.00 |
| **232** | tfidf-1000_forpint | 0 to 730 days | mean | nan | False | 0.00 | 71783 | 0.00 |
| **233** | tfidf-1000_forsat | 0 to 730 days | mean | nan | False | 0.00 | 36777 | 0.00 |
| **234** | tfidf-1000_forskellige | 0 to 730 days | mean | nan | False | 0.00 | 106628 | 0.00 |
| **235** | tfidf-1000_forstå | 0 to 730 days | mean | nan | False | 0.00 | 72960 | 0.00 |
| **236** | tfidf-1000_forståelse | 0 to 730 days | mean | nan | False | 0.00 | 58125 | 0.00 |
| **237** | tfidf-1000_forstår | 0 to 730 days | mean | nan | False | 0.00 | 58680 | 0.00 |
| **238** | tfidf-1000_forsænket | 0 to 730 days | mean | nan | False | 0.00 | 79982 | 0.00 |
| **239** | tfidf-1000_forsænket stemningsleje | 0 to 730 days | mean | nan | False | 0.00 | 50542 | 0.00 |
| **240** | tfidf-1000_forsøg | 0 to 730 days | mean | nan | False | 0.00 | 43326 | 0.00 |
| **241** | tfidf-1000_forsøge | 0 to 730 days | mean | nan | False | 0.00 | 83019 | 0.00 |
| **242** | tfidf-1000_forsøger | 0 to 730 days | mean | nan | False | 0.00 | 99958 | 0.00 |
| **243** | tfidf-1000_forsøges | 0 to 730 days | mean | nan | False | 0.00 | 30867 | 0.00 |
| **244** | tfidf-1000_forsøgt | 0 to 730 days | mean | nan | False | 0.00 | 98256 | 0.00 |
| **245** | tfidf-1000_fortalt | 0 to 730 days | mean | nan | False | 0.00 | 65801 | 0.00 |
| **246** | tfidf-1000_fortalte | 0 to 730 days | mean | nan | False | 0.00 | 36632 | 0.00 |

| | Feature name | Lookbehind period | Resolve multiple | Fallback strategy | Static | Mean | N. unique | Proportion using fallback |
|---|---|---|---|---|---|---|---|---|
| **247** | tfidf-1000_fortsætte | 0 to 730 days | mean | nan | False | 0.00 | 78777 | 0.00 |
| **248** | tfidf-1000_fortsætter | 0 to 730 days | mean | nan | False | 0.00 | 61148 | 0.00 |
| **249** | tfidf-1000_fortælle | 0 to 730 days | mean | nan | False | 0.00 | 93133 | 0.00 |
| **250** | tfidf-1000_fortæller | 0 to 730 days | mean | nan | False | 0.02 | 191501 | 0.00 |
| **251** | tfidf-1000_fortæller pt | 0 to 730 days | mean | nan | False | 0.00 | 60548 | 0.00 |
| **252** | tfidf-1000_forvirret | 0 to 730 days | mean | nan | False | 0.00 | 38959 | 0.00 |
| **253** | tfidf-1000_forværring | 0 to 730 days | mean | nan | False | 0.00 | 73935 | 0.00 |
| **254** | tfidf-1000_forældre | 0 to 730 days | mean | nan | False | 0.01 | 87203 | 0.00 |
| **255** | tfidf-1000_forældrene | 0 to 730 days | mean | nan | False | 0.00 | 68647 | 0.00 |
| **256** | tfidf-1000_fraset | 0 to 730 days | mean | nan | False | 0.00 | 36656 | 0.00 |
| **257** | tfidf-1000_fredag | 0 to 730 days | mean | nan | False | 0.00 | 79395 | 0.00 |
| **258** | tfidf-1000_frem | 0 to 730 days | mean | nan | False | 0.00 | 124720 | 0.00 |
| **259** | tfidf-1000_fremstår | 0 to 730 days | mean | nan | False | 0.01 | 137473 | 0.00 |
| **260** | tfidf-1000_fremtiden | 0 to 730 days | mean | nan | False | 0.00 | 81727 | 0.00 |
| **261** | tfidf-1000_fremtræder | 0 to 730 days | mean | nan | False | 0.01 | 90612 | 0.00 |
| **262** | tfidf-1000_frokost | 0 to 730 days | mean | nan | False | 0.00 | 31687 | 0.00 |
| **263** | tfidf-1000_frustreret | 0 to 730 days | mean | nan | False | 0.00 | 56136 | 0.00 |
| **264** | tfidf-1000_frygt | 0 to 730 days | mean | nan | False | 0.00 | 52198 | 0.00 |

| | Feature name | Lookbehind period | Resolve multiple | Fallback strategy | Static | Mean | N. unique | Proportion using fallback |
|---|---|---|---|---|---|---|---|---|
| **265** | tfidf-1000_frygter | 0 to 730 days | mean | nan | False | 0.00 | 50306 | 0.00 |
| **266** | tfidf-1000_fulgt | 0 to 730 days | mean | nan | False | 0.00 | 54695 | 0.00 |
| **267** | tfidf-1000_fundet | 0 to 730 days | mean | nan | False | 0.00 | 75968 | 0.00 |
| **268** | tfidf-1000_fylder | 0 to 730 days | mean | nan | False | 0.00 | 85819 | 0.00 |
| **269** | tfidf-1000_fysisk | 0 to 730 days | mean | nan | False | 0.00 | 95471 | 0.00 |
| **270** | tfidf-1000_fysiske | 0 to 730 days | mean | nan | False | 0.00 | 56694 | 0.00 |
| **271** | tfidf-1000_fået | 0 to 730 days | mean | nan | False | 0.01 | 185867 | 0.00 |
| **272** | tfidf-1000_fået bedre | 0 to 730 days | mean | nan | False | 0.00 | 54014 | 0.00 |
| **273** | tfidf-1000_fælles | 0 to 730 days | mean | nan | False | 0.00 | 53227 | 0.00 |
| **274** | tfidf-1000_fællesmiljøet | 0 to 730 days | mean | nan | False | 0.00 | 14533 | 0.00 |
| **275** | tfidf-1000_føle | 0 to 730 days | mean | nan | False | 0.00 | 85304 | 0.00 |
| **276** | tfidf-1000_følelse | 0 to 730 days | mean | nan | False | 0.00 | 74818 | 0.00 |
| **277** | tfidf-1000_følelser | 0 to 730 days | mean | nan | False | 0.00 | 87616 | 0.00 |
| **278** | tfidf-1000_føler | 0 to 730 days | mean | nan | False | 0.01 | 184533 | 0.00 |
| **279** | tfidf-1000_følges | 0 to 730 days | mean | nan | False | 0.00 | 55498 | 0.00 |
| **280** | tfidf-1000_følt | 0 to 730 days | mean | nan | False | 0.00 | 80291 | 0.00 |
| **281** | tfidf-1000_følte | 0 to 730 days | mean | nan | False | 0.00 | 77671 | 0.00 |
| **282** | tfidf-1000_gange | 0 to 730 days | mean | nan | False | 0.01 | 177019 | 0.00 |

| | Feature name | Lookbehind period | Resolve multiple | Fallback strategy | Static | Mean | N. unique | Proportion using fallback |
|---|---|---|---|---|---|---|---|---|
| **283** | tfidf-1000_gangen | 0 to 730 days | mean | nan | False | 0.00 | 68731 | 0.00 |
| **284** | tfidf-1000_ganske | 0 to 730 days | mean | nan | False | 0.00 | 57157 | 0.00 |
| **285** | tfidf-1000_gav | 0 to 730 days | mean | nan | False | 0.00 | 48384 | 0.00 |
| **286** | tfidf-1000_generelt | 0 to 730 days | mean | nan | False | 0.00 | 103167 | 0.00 |
| **287** | tfidf-1000_gik | 0 to 730 days | mean | nan | False | 0.00 | 105878 | 0.00 |
| **288** | tfidf-1000_give | 0 to 730 days | mean | nan | False | 0.00 | 112976 | 0.00 |
| **289** | tfidf-1000_gives | 0 to 730 days | mean | nan | False | 0.00 | 55607 | 0.00 |
| **290** | tfidf-1000_glad | 0 to 730 days | mean | nan | False | 0.01 | 153636 | 0.00 |
| **291** | tfidf-1000_glæde | 0 to 730 days | mean | nan | False | 0.00 | 83264 | 0.00 |
| **292** | tfidf-1000_glæder | 0 to 730 days | mean | nan | False | 0.00 | 58364 | 0.00 |
| **293** | tfidf-1000_gode | 0 to 730 days | mean | nan | False | 0.00 | 108818 | 0.00 |
| **294** | tfidf-1000_grad | 0 to 730 days | mean | nan | False | 0.00 | 105750 | 0.00 |
| **295** | tfidf-1000_grund | 0 to 730 days | mean | nan | False | 0.00 | 75729 | 0.00 |
| **296** | tfidf-1000_grundet | 0 to 730 days | mean | nan | False | 0.01 | 112341 | 0.00 |
| **297** | tfidf-1000_gruppe | 0 to 730 days | mean | nan | False | 0.01 | 61480 | 0.00 |
| **298** | tfidf-1000_grådlabil | 0 to 730 days | mean | nan | False | 0.01 | 85475 | 0.00 |
| **299** | tfidf-1000_grædende | 0 to 730 days | mean | nan | False | 0.00 | 67250 | 0.00 |
| **300** | tfidf-1000_græder | 0 to 730 days | mean | nan | False | 0.00 | 63524 | 0.00 |

| | Feature name | Lookbehind period | Resolve multiple | Fallback strategy | Static | Mean | N. unique | Proportion using fallback |
|---|---|---|---|---|---|---|---|---|
| **301** | tfidf-1000_gulvet | 0 to 730 days | mean | nan | False | 0.00 | 27150 | 0.00 |
| **302** | tfidf-1000_gået | 0 to 730 days | mean | nan | False | 0.00 | 126740 | 0.00 |
| **303** | tfidf-1000_gåtur | 0 to 730 days | mean | nan | False | 0.00 | 31142 | 0.00 |
| **304** | tfidf-1000_gøremål | 0 to 730 days | mean | nan | False | 0.00 | 52810 | 0.00 |
| **305** | tfidf-1000_hallucinationer | 0 to 730 days | mean | nan | False | 0.00 | 54265 | 0.00 |
| **306** | tfidf-1000_handle | 0 to 730 days | mean | nan | False | 0.00 | 74656 | 0.00 |
| **307** | tfidf-1000_handler | 0 to 730 days | mean | nan | False | 0.00 | 70122 | 0.00 |
| **308** | tfidf-1000_hash | 0 to 730 days | mean | nan | False | 0.00 | 29067 | 0.00 |
| **309** | tfidf-1000_haven | 0 to 730 days | mean | nan | False | 0.00 | 25409 | 0.00 |
| **310** | tfidf-1000_henblik | 0 to 730 days | mean | nan | False | 0.00 | 75502 | 0.00 |
| **311** | tfidf-1000_hente | 0 to 730 days | mean | nan | False | 0.00 | 46841 | 0.00 |
| **312** | tfidf-1000_hentet | 0 to 730 days | mean | nan | False | 0.00 | 35154 | 0.00 |
| **313** | tfidf-1000_henvende | 0 to 730 days | mean | nan | False | 0.00 | 43612 | 0.00 |
| **314** | tfidf-1000_henvender | 0 to 730 days | mean | nan | False | 0.00 | 41479 | 0.00 |
| **315** | tfidf-1000_henvises | 0 to 730 days | mean | nan | False | 0.01 | 91191 | 0.00 |
| **316** | tfidf-1000_henvisning | 0 to 730 days | mean | nan | False | 0.00 | 73888 | 0.00 |
| **317** | tfidf-1000_henvist | 0 to 730 days | mean | nan | False | 0.02 | 152921 | 0.00 |
| **318** | tfidf-1000_herfor | 0 to 730 days | mean | nan | False | 0.00 | 55247 | 0.00 |

| | Feature name | Lookbehind period | Resolve multiple | Fallback strategy | Static | Mean | N. unique | Proportion using fallback |
|---|---|---|---|---|---|---|---|---|
| **319** | tfidf-1000_herfra | 0 to 730 days | mean | nan | False | 0.00 | 77104 | 0.00 |
| **320** | tfidf-1000_herom | 0 to 730 days | mean | nan | False | 0.00 | 49655 | 0.00 |
| **321** | tfidf-1000_hertil | 0 to 730 days | mean | nan | False | 0.00 | 79549 | 0.00 |
| **322** | tfidf-1000_herunder | 0 to 730 days | mean | nan | False | 0.00 | 62860 | 0.00 |
| **323** | tfidf-1000_hinanden | 0 to 730 days | mean | nan | False | 0.00 | 63392 | 0.00 |
| **324** | tfidf-1000_hjem | 0 to 730 days | mean | nan | False | 0.01 | 132727 | 0.00 |
| **325** | tfidf-1000_hjemme | 0 to 730 days | mean | nan | False | 0.01 | 125705 | 0.00 |
| **326** | tfidf-1000_hjemmebesøg | 0 to 730 days | mean | nan | False | 0.00 | 36745 | 0.00 |
| **327** | tfidf-1000_hjemmet | 0 to 730 days | mean | nan | False | 0.00 | 98804 | 0.00 |
| **328** | tfidf-1000_hjertebanken | 0 to 730 days | mean | nan | False | 0.00 | 77772 | 0.00 |
| **329** | tfidf-1000_hjulpet | 0 to 730 days | mean | nan | False | 0.00 | 68394 | 0.00 |
| **330** | tfidf-1000_hjælp | 0 to 730 days | mean | nan | False | 0.01 | 142067 | 0.00 |
| **331** | tfidf-1000_hjælpe | 0 to 730 days | mean | nan | False | 0.00 | 110321 | 0.00 |
| **332** | tfidf-1000_hjælper | 0 to 730 days | mean | nan | False | 0.00 | 99656 | 0.00 |
| **333** | tfidf-1000_holde | 0 to 730 days | mean | nan | False | 0.01 | 141547 | 0.00 |
| **334** | tfidf-1000_holdt | 0 to 730 days | mean | nan | False | 0.00 | 66324 | 0.00 |
| **335** | tfidf-1000_hovedet | 0 to 730 days | mean | nan | False | 0.00 | 97942 | 0.00 |
| **336** | tfidf-1000_hovedpine | 0 to 730 days | mean | nan | False | 0.00 | 72763 | 0.00 |

| | Feature name | Lookbehind period | Resolve multiple | Fallback strategy | Static | Mean | N. unique | Proportion using fallback |
|---|---|---|---|---|---|---|---|---|
| **337** | tfidf-1000_humør | 0 to 730 days | mean | nan | False | 0.00 | 104296 | 0.00 |
| **338** | tfidf-1000_humøret | 0 to 730 days | mean | nan | False | 0.00 | 84326 | 0.00 |
| **339** | tfidf-1000_hurtig | 0 to 730 days | mean | nan | False | 0.00 | 47697 | 0.00 |
| **340** | tfidf-1000_hurtigt | 0 to 730 days | mean | nan | False | 0.00 | 109468 | 0.00 |
| **341** | tfidf-1000_hus | 0 to 730 days | mean | nan | False | 0.00 | 48445 | 0.00 |
| **342** | tfidf-1000_huset | 0 to 730 days | mean | nan | False | 0.00 | 52406 | 0.00 |
| **343** | tfidf-1000_huske | 0 to 730 days | mean | nan | False | 0.00 | 99058 | 0.00 |
| **344** | tfidf-1000_husker | 0 to 730 days | mean | nan | False | 0.00 | 61964 | 0.00 |
| **345** | tfidf-1000_hustru | 0 to 730 days | mean | nan | False | 0.00 | 19564 | 0.00 |
| **346** | tfidf-1000_hverdag | 0 to 730 days | mean | nan | False | 0.00 | 61244 | 0.00 |
| **347** | tfidf-1000_hverdagen | 0 to 730 days | mean | nan | False | 0.00 | 81192 | 0.00 |
| **348** | tfidf-1000_hverken | 0 to 730 days | mean | nan | False | 0.00 | 57631 | 0.00 |
| **349** | tfidf-1000_hvile | 0 to 730 days | mean | nan | False | 0.00 | 35980 | 0.00 |
| **350** | tfidf-1000_hvilket | 0 to 730 days | mean | nan | False | 0.01 | 175950 | 0.00 |
| **351** | tfidf-1000_hvilket pt | 0 to 730 days | mean | nan | False | 0.00 | 55646 | 0.00 |
| **352** | tfidf-1000_hvorvidt | 0 to 730 days | mean | nan | False | 0.00 | 68038 | 0.00 |
| **353** | tfidf-1000_håb | 0 to 730 days | mean | nan | False | 0.00 | 55258 | 0.00 |
| **354** | tfidf-1000_håber | 0 to 730 days | mean | nan | False | 0.00 | 59668 | 0.00 |

| | Feature name | Lookbehind period | Resolve multiple | Fallback strategy | Static | Mean | N. unique | Proportion using fallback |
|---|---|---|---|---|---|---|---|---|
| **355** | tfidf-1000_hånd | 0 to 730 days | mean | nan | False | 0.00 | 45039 | 0.00 |
| **356** | tfidf-1000_håndtere | 0 to 730 days | mean | nan | False | 0.00 | 56927 | 0.00 |
| **357** | tfidf-1000_hårdt | 0 to 730 days | mean | nan | False | 0.00 | 70932 | 0.00 |
| **358** | tfidf-1000_hænder | 0 to 730 days | mean | nan | False | 0.00 | 44641 | 0.00 |
| **359** | tfidf-1000_høj | 0 to 730 days | mean | nan | False | 0.00 | 57709 | 0.00 |
| **360** | tfidf-1000_højt | 0 to 730 days | mean | nan | False | 0.00 | 57520 | 0.00 |
| **361** | tfidf-1000_høre | 0 to 730 days | mean | nan | False | 0.00 | 94754 | 0.00 |
| **362** | tfidf-1000_idag | 0 to 730 days | mean | nan | False | 0.00 | 84198 | 0.00 |
| **363** | tfidf-1000_idet | 0 to 730 days | mean | nan | False | 0.01 | 127692 | 0.00 |
| **364** | tfidf-1000_ifht | 0 to 730 days | mean | nan | False | 0.00 | 27835 | 0.00 |
| **365** | tfidf-1000_ifm | 0 to 730 days | mean | nan | False | 0.00 | 56131 | 0.00 |
| **366** | tfidf-1000_ift | 0 to 730 days | mean | nan | False | 0.01 | 106538 | 0.00 |
| **367** | tfidf-1000_ifølge | 0 to 730 days | mean | nan | False | 0.00 | 50101 | 0.00 |
| **368** | tfidf-1000_igang | 0 to 730 days | mean | nan | False | 0.00 | 80039 | 0.00 |
| **369** | tfidf-1000_igår | 0 to 730 days | mean | nan | False | 0.00 | 51834 | 0.00 |
| **370** | tfidf-1000_imorgen | 0 to 730 days | mean | nan | False | 0.00 | 46648 | 0.00 |
| **371** | tfidf-1000_imødekommende | 0 to 730 days | mean | nan | False | 0.00 | 68566 | 0.00 |
| **372** | tfidf-1000_inde | 0 to 730 days | mean | nan | False | 0.00 | 74032 | 0.00 |

| | Feature name | Lookbehind period | Resolve multiple | Fallback strategy | Static | Mean | N. unique | Proportion using fallback |
|---|---|---|---|---|---|---|---|---|
| **373** | tfidf-1000_inden | 0 to 730 days | mean | nan | False | 0.01 | 134670 | 0.00 |
| **374** | tfidf-1000_indenfor | 0 to 730 days | mean | nan | False | 0.00 | 63068 | 0.00 |
| **375** | tfidf-1000_indforstået | 0 to 730 days | mean | nan | False | 0.00 | 53055 | 0.00 |
| **376** | tfidf-1000_indgå | 0 to 730 days | mean | nan | False | 0.00 | 53969 | 0.00 |
| **377** | tfidf-1000_indimellem | 0 to 730 days | mean | nan | False | 0.00 | 78157 | 0.00 |
| **378** | tfidf-1000_indlagt | 0 to 730 days | mean | nan | False | 0.00 | 67159 | 0.00 |
| **379** | tfidf-1000_indlæggelse | 0 to 730 days | mean | nan | False | 0.00 | 57356 | 0.00 |
| **380** | tfidf-1000_indlæggelsen | 0 to 730 days | mean | nan | False | 0.00 | 38159 | 0.00 |
| **381** | tfidf-1000_indlægges | 0 to 730 days | mean | nan | False | 0.00 | 36532 | 0.00 |
| **382** | tfidf-1000_indre | 0 to 730 days | mean | nan | False | 0.00 | 68992 | 0.00 |
| **383** | tfidf-1000_indstillet | 0 to 730 days | mean | nan | False | 0.00 | 67290 | 0.00 |
| **384** | tfidf-1000_indtryk | 0 to 730 days | mean | nan | False | 0.00 | 59717 | 0.00 |
| **385** | tfidf-1000_informeres | 0 to 730 days | mean | nan | False | 0.00 | 80296 | 0.00 |
| **386** | tfidf-1000_informeret | 0 to 730 days | mean | nan | False | 0.00 | 80849 | 0.00 |
| **387** | tfidf-1000_initiativ | 0 to 730 days | mean | nan | False | 0.00 | 54170 | 0.00 |
| **388** | tfidf-1000_interesse | 0 to 730 days | mean | nan | False | 0.00 | 70157 | 0.00 |
| **389** | tfidf-1000_interesseret | 0 to 730 days | mean | nan | False | 0.00 | 78127 | 0.00 |
| **390** | tfidf-1000_irritabel | 0 to 730 days | mean | nan | False | 0.00 | 52353 | 0.00 |

| | Feature name | Lookbehind period | Resolve multiple | Fallback strategy | Static | Mean | N. unique | Proportion using fallback |
|---|---|---|---|---|---|---|---|---|
| **391** | tfidf-1000_irriteret | 0 to 730 days | mean | nan | False | 0.00 | 54377 | 0.00 |
| **392** | tfidf-1000_job | 0 to 730 days | mean | nan | False | 0.00 | 71474 | 0.00 |
| **393** | tfidf-1000_jul | 0 to 730 days | mean | nan | False | 0.00 | 48021 | 0.00 |
| **394** | tfidf-1000_kaffe | 0 to 730 days | mean | nan | False | 0.00 | 25122 | 0.00 |
| **395** | tfidf-1000_ked | 0 to 730 days | mean | nan | False | 0.01 | 130992 | 0.00 |
| **396** | tfidf-1000_kender | 0 to 730 days | mean | nan | False | 0.00 | 85197 | 0.00 |
| **397** | tfidf-1000_kendt | 0 to 730 days | mean | nan | False | 0.00 | 77065 | 0.00 |
| **398** | tfidf-1000_kg | 0 to 730 days | mean | nan | False | 0.00 | 65474 | 0.00 |
| **399** | tfidf-1000_kigger | 0 to 730 days | mean | nan | False | 0.00 | 58758 | 0.00 |
| **400** | tfidf-1000_kiosken | 0 to 730 days | mean | nan | False | 0.00 | 9919 | 0.00 |
| **401** | tfidf-1000_kl | 0 to 730 days | mean | nan | False | 0.04 | 172054 | 0.00 |
| **402** | tfidf-1000_kl 10 | 0 to 730 days | mean | nan | False | 0.00 | 41357 | 0.00 |
| **403** | tfidf-1000_kl 11 | 0 to 730 days | mean | nan | False | 0.00 | 41159 | 0.00 |
| **404** | tfidf-1000_kl 13 | 0 to 730 days | mean | nan | False | 0.00 | 40033 | 0.00 |
| **405** | tfidf-1000_kl 1300 | 0 to 730 days | mean | nan | False | 0.00 | 50131 | 0.00 |
| **406** | tfidf-1000_klager | 0 to 730 days | mean | nan | False | 0.01 | 82350 | 0.00 |
| **407** | tfidf-1000_klar | 0 to 730 days | mean | nan | False | 0.01 | 152921 | 0.00 |
| **408** | tfidf-1000_klar orienteret | 0 to 730 days | mean | nan | False | 0.00 | 83674 | 0.00 |

| | Feature name | Lookbehind period | Resolve multiple | Fallback strategy | Static | Mean | N. unique | Proportion using fallback |
|---|---|---|---|---|---|---|---|---|
| **409** | tfidf-1000_klare | 0 to 730 days | mean | nan | False | 0.00 | 102572 | 0.00 |
| **410** | tfidf-1000_klart | 0 to 730 days | mean | nan | False | 0.00 | 69306 | 0.00 |
| **411** | tfidf-1000_klinik | 0 to 730 days | mean | nan | False | 0.00 | 52587 | 0.00 |
| **412** | tfidf-1000_klinikken | 0 to 730 days | mean | nan | False | 0.00 | 47446 | 0.00 |
| **413** | tfidf-1000_kognitive | 0 to 730 days | mean | nan | False | 0.00 | 50297 | 0.00 |
| **414** | tfidf-1000_kollega | 0 to 730 days | mean | nan | False | 0.00 | 29549 | 0.00 |
| **415** | tfidf-1000_kommende | 0 to 730 days | mean | nan | False | 0.00 | 58438 | 0.00 |
| **416** | tfidf-1000_kommune | 0 to 730 days | mean | nan | False | 0.00 | 44825 | 0.00 |
| **417** | tfidf-1000_kommunen | 0 to 730 days | mean | nan | False | 0.00 | 89554 | 0.00 |
| **418** | tfidf-1000_koncentration | 0 to 730 days | mean | nan | False | 0.00 | 63467 | 0.00 |
| **419** | tfidf-1000_koncentrere | 0 to 730 days | mean | nan | False | 0.00 | 88374 | 0.00 |
| **420** | tfidf-1000_konkret | 0 to 730 days | mean | nan | False | 0.00 | 51752 | 0.00 |
| **421** | tfidf-1000_konkrete | 0 to 730 days | mean | nan | False | 0.00 | 60232 | 0.00 |
| **422** | tfidf-1000_konstant | 0 to 730 days | mean | nan | False | 0.00 | 82192 | 0.00 |
| **423** | tfidf-1000_kontakt | 0 to 730 days | mean | nan | False | 0.02 | 196878 | 0.00 |
| **424** | tfidf-1000_kontakt pt | 0 to 730 days | mean | nan | False | 0.00 | 66958 | 0.00 |
| **425** | tfidf-1000_kontakte | 0 to 730 days | mean | nan | False | 0.00 | 99932 | 0.00 |
| **426** | tfidf-1000_kontakten | 0 to 730 days | mean | nan | False | 0.01 | 118112 | 0.00 |

|  | Feature name | Lookbehind period | Resolve multiple | Fallback strategy | Static | Mean | N. unique | Proportion using fallback |
|---|---|---|---|---|---|---|---|---|
| **427** | tfidf-1000_kontakter | 0 to 730 days | mean | nan | False | 0.00 | 100226 | 0.00 |
| **428** | tfidf-1000_kontaktes | 0 to 730 days | mean | nan | False | 0.00 | 44995 | 0.00 |
| **429** | tfidf-1000_kontaktet | 0 to 730 days | mean | nan | False | 0.00 | 66348 | 0.00 |
| **430** | tfidf-1000_kontaktperson | 0 to 730 days | mean | nan | False | 0.00 | 54842 | 0.00 |
| **431** | tfidf-1000_kontoret | 0 to 730 days | mean | nan | False | 0.00 | 21732 | 0.00 |
| **432** | tfidf-1000_kontrol | 0 to 730 days | mean | nan | False | 0.00 | 71973 | 0.00 |
| **433** | tfidf-1000_kort | 0 to 730 days | mean | nan | False | 0.00 | 115531 | 0.00 |
| **434** | tfidf-1000_kortvarigt | 0 to 730 days | mean | nan | False | 0.00 | 42098 | 0.00 |
| **435** | tfidf-1000_kp | 0 to 730 days | mean | nan | False | 0.00 | 20034 | 0.00 |
| **436** | tfidf-1000_kr | 0 to 730 days | mean | nan | False | 0.00 | 25922 | 0.00 |
| **437** | tfidf-1000_krav | 0 to 730 days | mean | nan | False | 0.00 | 58244 | 0.00 |
| **438** | tfidf-1000_kroppen | 0 to 730 days | mean | nan | False | 0.00 | 87267 | 0.00 |
| **439** | tfidf-1000_kunnet | 0 to 730 days | mean | nan | False | 0.00 | 80121 | 0.00 |
| **440** | tfidf-1000_kvalme | 0 to 730 days | mean | nan | False | 0.00 | 57245 | 0.00 |
| **441** | tfidf-1000_kvinde | 0 to 730 days | mean | nan | False | 0.00 | 63120 | 0.00 |
| **442** | tfidf-1000_kæreste | 0 to 730 days | mean | nan | False | 0.01 | 104894 | 0.00 |
| **443** | tfidf-1000_kæresten | 0 to 730 days | mean | nan | False | 0.01 | 62832 | 0.00 |
| **444** | tfidf-1000_købe | 0 to 730 days | mean | nan | False | 0.00 | 47669 | 0.00 |

| | Feature name | Lookbehind period | Resolve multiple | Fallback strategy | Static | Mean | N. unique | Proportion using fallback |
|---|---|---|---|---|---|---|---|---|
| **445** | tfidf-1000_køkkenet | 0 to 730 days | mean | nan | False | 0.00 | 28587 | 0.00 |
| **446** | tfidf-1000_køre | 0 to 730 days | mean | nan | False | 0.00 | 71119 | 0.00 |
| **447** | tfidf-1000_kører | 0 to 730 days | mean | nan | False | 0.00 | 75627 | 0.00 |
| **448** | tfidf-1000_lade | 0 to 730 days | mean | nan | False | 0.00 | 72380 | 0.00 |
| **449** | tfidf-1000_lagt | 0 to 730 days | mean | nan | False | 0.00 | 71215 | 0.00 |
| **450** | tfidf-1000_lang | 0 to 730 days | mean | nan | False | 0.00 | 95526 | 0.00 |
| **451** | tfidf-1000_lang tid | 0 to 730 days | mean | nan | False | 0.00 | 67861 | 0.00 |
| **452** | tfidf-1000_langt | 0 to 730 days | mean | nan | False | 0.00 | 70308 | 0.00 |
| **453** | tfidf-1000_latenstid | 0 to 730 days | mean | nan | False | 0.00 | 79031 | 0.00 |
| **454** | tfidf-1000_laver | 0 to 730 days | mean | nan | False | 0.00 | 96507 | 0.00 |
| **455** | tfidf-1000_laves | 0 to 730 days | mean | nan | False | 0.00 | 65096 | 0.00 |
| **456** | tfidf-1000_lejlighed | 0 to 730 days | mean | nan | False | 0.01 | 90345 | 0.00 |
| **457** | tfidf-1000_lejligheden | 0 to 730 days | mean | nan | False | 0.00 | 43419 | 0.00 |
| **458** | tfidf-1000_let | 0 to 730 days | mean | nan | False | 0.01 | 139443 | 0.00 |
| **459** | tfidf-1000_lettere | 0 to 730 days | mean | nan | False | 0.00 | 102802 | 0.00 |
| **460** | tfidf-1000_leve | 0 to 730 days | mean | nan | False | 0.00 | 73976 | 0.00 |
| **461** | tfidf-1000_lide | 0 to 730 days | mean | nan | False | 0.00 | 70864 | 0.00 |
| **462** | tfidf-1000_lidelse | 0 to 730 days | mean | nan | False | 0.00 | 43249 | 0.00 |

| | Feature name | Lookbehind period | Resolve multiple | Fallback strategy | Static | Mean | N. unique | Proportion using fallback |
|---|---|---|---|---|---|---|---|---|
| **463** | tfidf-1000_ligeledes | 0 to 730 days | mean | nan | False | 0.00 | 106560 | 0.00 |
| **464** | tfidf-1000_liv | 0 to 730 days | mean | nan | False | 0.00 | 119782 | 0.00 |
| **465** | tfidf-1000_livet | 0 to 730 days | mean | nan | False | 0.00 | 89172 | 0.00 |
| **466** | tfidf-1000_lov | 0 to 730 days | mean | nan | False | 0.00 | 51514 | 0.00 |
| **467** | tfidf-1000_lukket | 0 to 730 days | mean | nan | False | 0.00 | 38202 | 0.00 |
| **468** | tfidf-1000_lyder | 0 to 730 days | mean | nan | False | 0.00 | 47612 | 0.00 |
| **469** | tfidf-1000_lyst | 0 to 730 days | mean | nan | False | 0.00 | 136427 | 0.00 |
| **470** | tfidf-1000_læge | 0 to 730 days | mean | nan | False | 0.02 | 178503 | 0.00 |
| **471** | tfidf-1000_lægen | 0 to 730 days | mean | nan | False | 0.00 | 45437 | 0.00 |
| **472** | tfidf-1000_lægesamtale | 0 to 730 days | mean | nan | False | 0.00 | 53118 | 0.00 |
| **473** | tfidf-1000_lægge | 0 to 730 days | mean | nan | False | 0.00 | 53434 | 0.00 |
| **474** | tfidf-1000_lægger | 0 to 730 days | mean | nan | False | 0.00 | 48087 | 0.00 |
| **475** | tfidf-1000_længe | 0 to 730 days | mean | nan | False | 0.00 | 86837 | 0.00 |
| **476** | tfidf-1000_læse | 0 to 730 days | mean | nan | False | 0.00 | 80458 | 0.00 |
| **477** | tfidf-1000_løbet | 0 to 730 days | mean | nan | False | 0.00 | 120684 | 0.00 |
| **478** | tfidf-1000_løbet dagen | 0 to 730 days | mean | nan | False | 0.00 | 58285 | 0.00 |
| **479** | tfidf-1000_lørdag | 0 to 730 days | mean | nan | False | 0.00 | 36257 | 0.00 |
| **480** | tfidf-1000_mad | 0 to 730 days | mean | nan | False | 0.00 | 84761 | 0.00 |

| | Feature name | Lookbehind period | Resolve multiple | Fallback strategy | Static | Mean | N. unique | Proportion using fallback |
|---|---|---|---|---|---|---|---|---|
| **481** | tfidf-1000_magter | 0 to 730 days | mean | nan | False | 0.00 | 63698 | 0.00 |
| **482** | tfidf-1000_mandag | 0 to 730 days | mean | nan | False | 0.01 | 95590 | 0.00 |
| **483** | tfidf-1000_manglende | 0 to 730 days | mean | nan | False | 0.00 | 106246 | 0.00 |
| **484** | tfidf-1000_mangler | 0 to 730 days | mean | nan | False | 0.00 | 73749 | 0.00 |
| **485** | tfidf-1000_mareridt | 0 to 730 days | mean | nan | False | 0.00 | 53218 | 0.00 |
| **486** | tfidf-1000_maven | 0 to 730 days | mean | nan | False | 0.00 | 53692 | 0.00 |
| **487** | tfidf-1000_mdr | 0 to 730 days | mean | nan | False | 0.00 | 90583 | 0.00 |
| **488** | tfidf-1000_medicin | 0 to 730 days | mean | nan | False | 0.01 | 141798 | 0.00 |
| **489** | tfidf-1000_medicinen | 0 to 730 days | mean | nan | False | 0.00 | 82153 | 0.00 |
| **490** | tfidf-1000_medicinsk | 0 to 730 days | mean | nan | False | 0.00 | 88966 | 0.00 |
| **491** | tfidf-1000_medicinsk behandling | 0 to 730 days | mean | nan | False | 0.00 | 69425 | 0.00 |
| **492** | tfidf-1000_medicinske | 0 to 730 days | mean | nan | False | 0.00 | 60744 | 0.00 |
| **493** | tfidf-1000_medicinske behandling | 0 to 730 days | mean | nan | False | 0.00 | 51743 | 0.00 |
| **494** | tfidf-1000_medpatienter | 0 to 730 days | mean | nan | False | 0.00 | 21432 | 0.00 |
| **495** | tfidf-1000_medpt | 0 to 730 days | mean | nan | False | 0.00 | 26864 | 0.00 |
| **496** | tfidf-1000_mener | 0 to 730 days | mean | nan | False | 0.01 | 147513 | 0.00 |
| **497** | tfidf-1000_mening | 0 to 730 days | mean | nan | False | 0.00 | 56591 | 0.00 |
| **498** | tfidf-1000_mennesker | 0 to 730 days | mean | nan | False | 0.01 | 127228 | 0.00 |

|  | Feature name | Lookbehind period | Resolve multiple | Fallback strategy | Static | Mean | N. unique | Proportion using fallback |
|---|---|---|---|---|---|---|---|---|
| **499** | tfidf-1000_mentor | 0 to 730 days | mean | nan | False | 0.00 | 35115 | 0.00 |
| **500** | tfidf-1000_meste | 0 to 730 days | mean | nan | False | 0.00 | 86849 | 0.00 |
| **501** | tfidf-1000_mette | 0 to 730 days | mean | nan | False | 0.00 | 28226 | 0.00 |
| **502** | tfidf-1000_mg | 0 to 730 days | mean | nan | False | 0.01 | 106364 | 0.00 |
| **503** | tfidf-1000_mhp | 0 to 730 days | mean | nan | False | 0.01 | 139605 | 0.00 |
| **504** | tfidf-1000_middag | 0 to 730 days | mean | nan | False | 0.00 | 36663 | 0.00 |
| **505** | tfidf-1000_miljøet | 0 to 730 days | mean | nan | False | 0.00 | 29007 | 0.00 |
| **506** | tfidf-1000_mimik | 0 to 730 days | mean | nan | False | 0.00 | 76154 | 0.00 |
| **507** | tfidf-1000_minutter | 0 to 730 days | mean | nan | False | 0.00 | 39459 | 0.00 |
| **508** | tfidf-1000_misbrug | 0 to 730 days | mean | nan | False | 0.00 | 47962 | 0.00 |
| **509** | tfidf-1000_mistanke | 0 to 730 days | mean | nan | False | 0.00 | 77167 | 0.00 |
| **510** | tfidf-1000_mm | 0 to 730 days | mean | nan | False | 0.00 | 61443 | 0.00 |
| **511** | tfidf-1000_moderen | 0 to 730 days | mean | nan | False | 0.00 | 77738 | 0.00 |
| **512** | tfidf-1000_modtage | 0 to 730 days | mean | nan | False | 0.00 | 38761 | 0.00 |
| **513** | tfidf-1000_modtagelsen | 0 to 730 days | mean | nan | False | 0.00 | 30052 | 0.00 |
| **514** | tfidf-1000_modtager | 0 to 730 days | mean | nan | False | 0.00 | 44136 | 0.00 |
| **515** | tfidf-1000_modtaget | 0 to 730 days | mean | nan | False | 0.00 | 52355 | 0.00 |
| **516** | tfidf-1000_mor | 0 to 730 days | mean | nan | False | 0.01 | 141350 | 0.00 |

| | Feature name | Lookbehind period | Resolve multiple | Fallback strategy | Static | Mean | N. unique | Proportion using fallback |
|---|---|---|---|---|---|---|---|---|
| **517** | tfidf-1000_moren | 0 to 730 days | mean | nan | False | 0.00 | 37332 | 0.00 |
| **518** | tfidf-1000_morgen | 0 to 730 days | mean | nan | False | 0.01 | 106039 | 0.00 |
| **519** | tfidf-1000_morgenen | 0 to 730 days | mean | nan | False | 0.00 | 79049 | 0.00 |
| **520** | tfidf-1000_morgenmad | 0 to 730 days | mean | nan | False | 0.00 | 34619 | 0.00 |
| **521** | tfidf-1000_morgenstunden | 0 to 730 days | mean | nan | False | 0.00 | 20512 | 0.00 |
| **522** | tfidf-1000_motiveret | 0 to 730 days | mean | nan | False | 0.00 | 74563 | 0.00 |
| **523** | tfidf-1000_motorisk | 0 to 730 days | mean | nan | False | 0.00 | 45437 | 0.00 |
| **524** | tfidf-1000_mulighed | 0 to 730 days | mean | nan | False | 0.00 | 102971 | 0.00 |
| **525** | tfidf-1000_muligheden | 0 to 730 days | mean | nan | False | 0.00 | 64539 | 0.00 |
| **526** | tfidf-1000_muligt | 0 to 730 days | mean | nan | False | 0.00 | 97888 | 0.00 |
| **527** | tfidf-1000_muligvis | 0 to 730 days | mean | nan | False | 0.00 | 62992 | 0.00 |
| **528** | tfidf-1000_musik | 0 to 730 days | mean | nan | False | 0.00 | 41577 | 0.00 |
| **529** | tfidf-1000_måde | 0 to 730 days | mean | nan | False | 0.00 | 114206 | 0.00 |
| **530** | tfidf-1000_måltider | 0 to 730 days | mean | nan | False | 0.00 | 30300 | 0.00 |
| **531** | tfidf-1000_måned | 0 to 730 days | mean | nan | False | 0.00 | 95129 | 0.00 |
| **532** | tfidf-1000_måneder | 0 to 730 days | mean | nan | False | 0.00 | 95104 | 0.00 |
| **533** | tfidf-1000_mærke | 0 to 730 days | mean | nan | False | 0.00 | 113036 | 0.00 |
| **534** | tfidf-1000_mærker | 0 to 730 days | mean | nan | False | 0.00 | 80374 | 0.00 |

| | Feature name | Lookbehind period | Resolve multiple | Fallback strategy | Static | Mean | N. unique | Proportion using fallback |
|---|---|---|---|---|---|---|---|---|
| **535** | tfidf-1000_møde | 0 to 730 days | mean | nan | False | 0.01 | 115349 | 0.00 |
| **536** | tfidf-1000_møder | 0 to 730 days | mean | nan | False | 0.01 | 107157 | 0.00 |
| **537** | tfidf-1000_mødet | 0 to 730 days | mean | nan | False | 0.00 | 42644 | 0.00 |
| **538** | tfidf-1000_mødt | 0 to 730 days | mean | nan | False | 0.01 | 49630 | 0.00 |
| **539** | tfidf-1000_nada | 0 to 730 days | mean | nan | False | 0.00 | 19896 | 0.00 |
| **540** | tfidf-1000_nat | 0 to 730 days | mean | nan | False | 0.00 | 84885 | 0.00 |
| **541** | tfidf-1000_natten | 0 to 730 days | mean | nan | False | 0.00 | 117971 | 0.00 |
| **542** | tfidf-1000_nattesøvn | 0 to 730 days | mean | nan | False | 0.00 | 56171 | 0.00 |
| **543** | tfidf-1000_nede | 0 to 730 days | mean | nan | False | 0.00 | 40915 | 0.00 |
| **544** | tfidf-1000_nedsat | 0 to 730 days | mean | nan | False | 0.01 | 136288 | 0.00 |
| **545** | tfidf-1000_negative | 0 to 730 days | mean | nan | False | 0.00 | 74419 | 0.00 |
| **546** | tfidf-1000_nemt | 0 to 730 days | mean | nan | False | 0.00 | 56729 | 0.00 |
| **547** | tfidf-1000_nervøs | 0 to 730 days | mean | nan | False | 0.00 | 85391 | 0.00 |
| **548** | tfidf-1000_neutral | 0 to 730 days | mean | nan | False | 0.00 | 54005 | 0.00 |
| **549** | tfidf-1000_neutralt | 0 to 730 days | mean | nan | False | 0.01 | 110993 | 0.00 |
| **550** | tfidf-1000_neutralt stemningsleje | 0 to 730 days | mean | nan | False | 0.00 | 65602 | 0.00 |
| **551** | tfidf-1000_nogenlunde | 0 to 730 days | mean | nan | False | 0.00 | 55172 | 0.00 |
| **552** | tfidf-1000_normal | 0 to 730 days | mean | nan | False | 0.00 | 86845 | 0.00 |

| | Feature name | Lookbehind period | Resolve multiple | Fallback strategy | Static | Mean | N. unique | Proportion using fallback |
|---|---|---|---|---|---|---|---|---|
| **553** | tfidf-1000_normalt | 0 to 730 days | mean | nan | False | 0.00 | 94714 | 0.00 |
| **554** | tfidf-1000_notat | 0 to 730 days | mean | nan | False | 0.00 | 61334 | 0.00 |
| **555** | tfidf-1000_nuværende | 0 to 730 days | mean | nan | False | 0.00 | 111330 | 0.00 |
| **556** | tfidf-1000_nærmere | 0 to 730 days | mean | nan | False | 0.00 | 87347 | 0.00 |
| **557** | tfidf-1000_nævner | 0 to 730 days | mean | nan | False | 0.00 | 69903 | 0.00 |
| **558** | tfidf-1000_nødt | 0 to 730 days | mean | nan | False | 0.00 | 57751 | 0.00 |
| **559** | tfidf-1000_nødvendigt | 0 to 730 days | mean | nan | False | 0.00 | 46292 | 0.00 |
| **560** | tfidf-1000_obs | 0 to 730 days | mean | nan | False | 0.00 | 61663 | 0.00 |
| **561** | tfidf-1000_observeres | 0 to 730 days | mean | nan | False | 0.00 | 27527 | 0.00 |
| **562** | tfidf-1000_observeret | 0 to 730 days | mean | nan | False | 0.00 | 26321 | 0.00 |
| **563** | tfidf-1000_ocd | 0 to 730 days | mean | nan | False | 0.01 | 40693 | 0.00 |
| **564** | tfidf-1000_ofte | 0 to 730 days | mean | nan | False | 0.01 | 154701 | 0.00 |
| **565** | tfidf-1000_ok | 0 to 730 days | mean | nan | False | 0.00 | 92193 | 0.00 |
| **566** | tfidf-1000_olanzapin | 0 to 730 days | mean | nan | False | 0.00 | 7491 | 0.00 |
| **567** | tfidf-1000_ondt | 0 to 730 days | mean | nan | False | 0.00 | 77196 | 0.00 |
| **568** | tfidf-1000_onsdag | 0 to 730 days | mean | nan | False | 0.00 | 73730 | 0.00 |
| **569** | tfidf-1000_opfordres | 0 to 730 days | mean | nan | False | 0.00 | 71431 | 0.00 |
| **570** | tfidf-1000_opfordret | 0 to 730 days | mean | nan | False | 0.00 | 63415 | 0.00 |

| | Feature name | Lookbehind period | Resolve multiple | Fallback strategy | Static | Mean | N. unique | Proportion using fallback |
|---|---|---|---|---|---|---|---|---|
| **571** | tfidf-1000_opfølgning | 0 to 730 days | mean | nan | False | 0.00 | 77373 | 0.00 |
| **572** | tfidf-1000_opgaver | 0 to 730 days | mean | nan | False | 0.00 | 74666 | 0.00 |
| **573** | tfidf-1000_opgivende | 0 to 730 days | mean | nan | False | 0.00 | 52554 | 0.00 |
| **574** | tfidf-1000_opholder | 0 to 730 days | mean | nan | False | 0.00 | 46218 | 0.00 |
| **575** | tfidf-1000_opholdt | 0 to 730 days | mean | nan | False | 0.00 | 40337 | 0.00 |
| **576** | tfidf-1000_opholdt stue | 0 to 730 days | mean | nan | False | 0.00 | 18487 | 0.00 |
| **577** | tfidf-1000_opleve | 0 to 730 days | mean | nan | False | 0.00 | 59101 | 0.00 |
| **578** | tfidf-1000_oplevede | 0 to 730 days | mean | nan | False | 0.00 | 88961 | 0.00 |
| **579** | tfidf-1000_oplevelse | 0 to 730 days | mean | nan | False | 0.00 | 84597 | 0.00 |
| **580** | tfidf-1000_oplevelser | 0 to 730 days | mean | nan | False | 0.00 | 82015 | 0.00 |
| **581** | tfidf-1000_oplever | 0 to 730 days | mean | nan | False | 0.01 | 181746 | 0.00 |
| **582** | tfidf-1000_opleves | 0 to 730 days | mean | nan | False | 0.00 | 57373 | 0.00 |
| **583** | tfidf-1000_oplevet | 0 to 730 days | mean | nan | False | 0.00 | 116813 | 0.00 |
| **584** | tfidf-1000_oplyser | 0 to 730 days | mean | nan | False | 0.01 | 105078 | 0.00 |
| **585** | tfidf-1000_opmærksom | 0 to 730 days | mean | nan | False | 0.00 | 84323 | 0.00 |
| **586** | tfidf-1000_opmærksomhed | 0 to 730 days | mean | nan | False | 0.00 | 48932 | 0.00 |
| **587** | tfidf-1000_oppe | 0 to 730 days | mean | nan | False | 0.00 | 51859 | 0.00 |
| **588** | tfidf-1000_opstart | 0 to 730 days | mean | nan | False | 0.01 | 105018 | 0.00 |

| | Feature name | Lookbehind period | Resolve multiple | Fallback strategy | Static | Mean | N. unique | Proportion using fallback |
|---|---|---|---|---|---|---|---|---|
| **589** | tfidf-1000_optaget | 0 to 730 days | mean | nan | False | 0.00 | 70461 | 0.00 |
| **590** | tfidf-1000_ord | 0 to 730 days | mean | nan | False | 0.00 | 68142 | 0.00 |
| **591** | tfidf-1000_orienteres | 0 to 730 days | mean | nan | False | 0.00 | 54439 | 0.00 |
| **592** | tfidf-1000_orienteret | 0 to 730 days | mean | nan | False | 0.01 | 130887 | 0.00 |
| **593** | tfidf-1000_orienteret tid | 0 to 730 days | mean | nan | False | 0.00 | 42002 | 0.00 |
| **594** | tfidf-1000_orlov | 0 to 730 days | mean | nan | False | 0.00 | 15855 | 0.00 |
| **595** | tfidf-1000_osv | 0 to 730 days | mean | nan | False | 0.00 | 50159 | 0.00 |
| **596** | tfidf-1000_ovenstående | 0 to 730 days | mean | nan | False | 0.00 | 50708 | 0.00 |
| **597** | tfidf-1000_overblik | 0 to 730 days | mean | nan | False | 0.00 | 49704 | 0.00 |
| **598** | tfidf-1000_overfor | 0 to 730 days | mean | nan | False | 0.00 | 120893 | 0.00 |
| **599** | tfidf-1000_overlæge | 0 to 730 days | mean | nan | False | 0.00 | 63323 | 0.00 |
| **600** | tfidf-1000_overskud | 0 to 730 days | mean | nan | False | 0.00 | 96936 | 0.00 |
| **601** | tfidf-1000_overskue | 0 to 730 days | mean | nan | False | 0.00 | 96340 | 0.00 |
| **602** | tfidf-1000_oxapax | 0 to 730 days | mean | nan | False | 0.00 | 19527 | 0.00 |
| **603** | tfidf-1000_par | 0 to 730 days | mean | nan | False | 0.00 | 125776 | 0.00 |
| **604** | tfidf-1000_par gange | 0 to 730 days | mean | nan | False | 0.00 | 50689 | 0.00 |
| **605** | tfidf-1000_paranoid | 0 to 730 days | mean | nan | False | 0.00 | 20882 | 0.00 |
| **606** | tfidf-1000_paranoide | 0 to 730 days | mean | nan | False | 0.00 | 19883 | 0.00 |

| | Feature name | Lookbehind period | Resolve multiple | Fallback strategy | Static | Mean | N. unique | Proportion using fallback |
|---|---|---|---|---|---|---|---|---|
| **607** | tfidf-1000_passe | 0 to 730 days | mean | nan | False | 0.00 | 71894 | 0.00 |
| **608** | tfidf-1000_passer | 0 to 730 days | mean | nan | False | 0.00 | 58038 | 0.00 |
| **609** | tfidf-1000_patient | 0 to 730 days | mean | nan | False | 0.00 | 29202 | 0.00 |
| **610** | tfidf-1000_patienten | 0 to 730 days | mean | nan | False | 0.02 | 97084 | 0.00 |
| **611** | tfidf-1000_patientens | 0 to 730 days | mean | nan | False | 0.00 | 46882 | 0.00 |
| **612** | tfidf-1000_patienter | 0 to 730 days | mean | nan | False | 0.00 | 24418 | 0.00 |
| **613** | tfidf-1000_penge | 0 to 730 days | mean | nan | False | 0.00 | 50700 | 0.00 |
| **614** | tfidf-1000_periode | 0 to 730 days | mean | nan | False | 0.00 | 111528 | 0.00 |
| **615** | tfidf-1000_perioder | 0 to 730 days | mean | nan | False | 0.00 | 95392 | 0.00 |
| **616** | tfidf-1000_person | 0 to 730 days | mean | nan | False | 0.00 | 61849 | 0.00 |
| **617** | tfidf-1000_personale | 0 to 730 days | mean | nan | False | 0.00 | 35553 | 0.00 |
| **618** | tfidf-1000_personalet | 0 to 730 days | mean | nan | False | 0.00 | 40994 | 0.00 |
| **619** | tfidf-1000_personer | 0 to 730 days | mean | nan | False | 0.00 | 44306 | 0.00 |
| **620** | tfidf-1000_personlighedsforstyrrelse | 0 to 730 days | mean | nan | False | 0.00 | 46830 | 0.00 |
| **621** | tfidf-1000_pga | 0 to 730 days | mean | nan | False | 0.01 | 132448 | 0.00 |
| **622** | tfidf-1000_plads | 0 to 730 days | mean | nan | False | 0.00 | 58856 | 0.00 |
| **623** | tfidf-1000_plaget | 0 to 730 days | mean | nan | False | 0.00 | 80474 | 0.00 |
| **624** | tfidf-1000_plan | 0 to 730 days | mean | nan | False | 0.00 | 93451 | 0.00 |

| | Feature name | Lookbehind period | Resolve multiple | Fallback strategy | Static | Mean | N. unique | Proportion using fallback |
|---|---|---|---|---|---|---|---|---|
| **625** | tfidf-1000_planen | 0 to 730 days | mean | nan | False | 0.00 | 58387 | 0.00 |
| **626** | tfidf-1000_planer | 0 to 730 days | mean | nan | False | 0.00 | 99679 | 0.00 |
| **627** | tfidf-1000_planlagt | 0 to 730 days | mean | nan | False | 0.00 | 75273 | 0.00 |
| **628** | tfidf-1000_plejer | 0 to 730 days | mean | nan | False | 0.00 | 65894 | 0.00 |
| **629** | tfidf-1000_pludselig | 0 to 730 days | mean | nan | False | 0.00 | 58769 | 0.00 |
| **630** | tfidf-1000_pn | 0 to 730 days | mean | nan | False | 0.00 | 49929 | 0.00 |
| **631** | tfidf-1000_pn medicin | 0 to 730 days | mean | nan | False | 0.00 | 21408 | 0.00 |
| **632** | tfidf-1000_politiet | 0 to 730 days | mean | nan | False | 0.00 | 28155 | 0.00 |
| **633** | tfidf-1000_positiv | 0 to 730 days | mean | nan | False | 0.00 | 57975 | 0.00 |
| **634** | tfidf-1000_pr | 0 to 730 days | mean | nan | False | 0.00 | 75513 | 0.00 |
| **635** | tfidf-1000_praktik | 0 to 730 days | mean | nan | False | 0.00 | 42265 | 0.00 |
| **636** | tfidf-1000_praktiske | 0 to 730 days | mean | nan | False | 0.00 | 52258 | 0.00 |
| **637** | tfidf-1000_presset | 0 to 730 days | mean | nan | False | 0.00 | 88335 | 0.00 |
| **638** | tfidf-1000_primært | 0 to 730 days | mean | nan | False | 0.00 | 76579 | 0.00 |
| **639** | tfidf-1000_problem | 0 to 730 days | mean | nan | False | 0.00 | 78782 | 0.00 |
| **640** | tfidf-1000_problemer | 0 to 730 days | mean | nan | False | 0.01 | 156668 | 0.00 |
| **641** | tfidf-1000_præget | 0 to 730 days | mean | nan | False | 0.00 | 96066 | 0.00 |
| **642** | tfidf-1000_prøve | 0 to 730 days | mean | nan | False | 0.00 | 78237 | 0.00 |

| | Feature name | Lookbehind period | Resolve multiple | Fallback strategy | Static | Mean | N. unique | Proportion using fallback |
|---|---|---|---|---|---|---|---|---|
| **643** | tfidf-1000_prøver | 0 to 730 days | mean | nan | False | 0.00 | 77146 | 0.00 |
| **644** | tfidf-1000_prøvet | 0 to 730 days | mean | nan | False | 0.00 | 54812 | 0.00 |
| **645** | tfidf-1000_psykiatrien | 0 to 730 days | mean | nan | False | 0.00 | 31435 | 0.00 |
| **646** | tfidf-1000_psykiatrisk | 0 to 730 days | mean | nan | False | 0.00 | 66562 | 0.00 |
| **647** | tfidf-1000_psykisk | 0 to 730 days | mean | nan | False | 0.01 | 119307 | 0.00 |
| **648** | tfidf-1000_psykiske | 0 to 730 days | mean | nan | False | 0.00 | 91052 | 0.00 |
| **649** | tfidf-1000_psykiske tilstand | 0 to 730 days | mean | nan | False | 0.00 | 55104 | 0.00 |
| **650** | tfidf-1000_psykolog | 0 to 730 days | mean | nan | False | 0.01 | 112656 | 0.00 |
| **651** | tfidf-1000_psykomotorisk | 0 to 730 days | mean | nan | False | 0.00 | 90635 | 0.00 |
| **652** | tfidf-1000_psykomotorisk tempo | 0 to 730 days | mean | nan | False | 0.00 | 61784 | 0.00 |
| **653** | tfidf-1000_psykose | 0 to 730 days | mean | nan | False | 0.00 | 28352 | 0.00 |
| **654** | tfidf-1000_psykotisk | 0 to 730 days | mean | nan | False | 0.00 | 102404 | 0.00 |
| **655** | tfidf-1000_psykotiske | 0 to 730 days | mean | nan | False | 0.00 | 96182 | 0.00 |
| **656** | tfidf-1000_psykotiske symptomer | 0 to 730 days | mean | nan | False | 0.00 | 80501 | 0.00 |
| **657** | tfidf-1000_pt | 0 to 730 days | mean | nan | False | 0.09 | 206157 | 0.00 |
| **658** | tfidf-1000_pt angiver | 0 to 730 days | mean | nan | False | 0.00 | 70350 | 0.00 |
| **659** | tfidf-1000_pt beskriver | 0 to 730 days | mean | nan | False | 0.00 | 97986 | 0.00 |
| **660** | tfidf-1000_pt fortæller | 0 to 730 days | mean | nan | False | 0.01 | 122382 | 0.00 |

| | Feature name | Lookbehind period | Resolve multiple | Fallback strategy | Static | Mean | N. unique | Proportion using fallback |
|---|---|---|---|---|---|---|---|---|
| **661** | tfidf-1000_pt fremstår | 0 to 730 days | mean | nan | False | 0.00 | 67749 | 0.00 |
| **662** | tfidf-1000_pt fremtræder | 0 to 730 days | mean | nan | False | 0.00 | 35337 | 0.00 |
| **663** | tfidf-1000_pt fået | 0 to 730 days | mean | nan | False | 0.00 | 76822 | 0.00 |
| **664** | tfidf-1000_pt føler | 0 to 730 days | mean | nan | False | 0.00 | 74946 | 0.00 |
| **665** | tfidf-1000_pt glad | 0 to 730 days | mean | nan | False | 0.00 | 36934 | 0.00 |
| **666** | tfidf-1000_pt mener | 0 to 730 days | mean | nan | False | 0.00 | 51100 | 0.00 |
| **667** | tfidf-1000_pt møder | 0 to 730 days | mean | nan | False | 0.01 | 51017 | 0.00 |
| **668** | tfidf-1000_pt opholdt | 0 to 730 days | mean | nan | False | 0.00 | 14159 | 0.00 |
| **669** | tfidf-1000_pt oplever | 0 to 730 days | mean | nan | False | 0.00 | 88426 | 0.00 |
| **670** | tfidf-1000_pt oplyser | 0 to 730 days | mean | nan | False | 0.00 | 51279 | 0.00 |
| **671** | tfidf-1000_pt pt | 0 to 730 days | mean | nan | False | 0.00 | 59083 | 0.00 |
| **672** | tfidf-1000_pt svært | 0 to 730 days | mean | nan | False | 0.00 | 79156 | 0.00 |
| **673** | tfidf-1000_pt tager | 0 to 730 days | mean | nan | False | 0.00 | 40138 | 0.00 |
| **674** | tfidf-1000_pt taget | 0 to 730 days | mean | nan | False | 0.00 | 35823 | 0.00 |
| **675** | tfidf-1000_pt udtryk | 0 to 730 days | mean | nan | False | 0.00 | 45811 | 0.00 |
| **676** | tfidf-1000_pt ut | 0 to 730 days | mean | nan | False | 0.00 | 47797 | 0.00 |
| **677** | tfidf-1000_pt venlig | 0 to 730 days | mean | nan | False | 0.00 | 16736 | 0.00 |
| **678** | tfidf-1000_pt virker | 0 to 730 days | mean | nan | False | 0.00 | 50404 | 0.00 |

| | Feature name | Lookbehind period | Resolve multiple | Fallback strategy | Static | Mean | N. unique | Proportion using fallback |
|---|---|---|---|---|---|---|---|---|
| **679** | tfidf-1000_pt ønsker | 0 to 730 days | mean | nan | False | 0.00 | 63007 | 0.00 |
| **680** | tfidf-1000_pts | 0 to 730 days | mean | nan | False | 0.01 | 148267 | 0.00 |
| **681** | tfidf-1000_ptsd | 0 to 730 days | mean | nan | False | 0.00 | 43194 | 0.00 |
| **682** | tfidf-1000_ptt | 0 to 730 days | mean | nan | False | 0.00 | 22414 | 0.00 |
| **683** | tfidf-1000_pårørende | 0 to 730 days | mean | nan | False | 0.00 | 44669 | 0.00 |
| **684** | tfidf-1000_påvirket | 0 to 730 days | mean | nan | False | 0.00 | 100985 | 0.00 |
| **685** | tfidf-1000_quetiapin | 0 to 730 days | mean | nan | False | 0.00 | 29080 | 0.00 |
| **686** | tfidf-1000_rask | 0 to 730 days | mean | nan | False | 0.00 | 54575 | 0.00 |
| **687** | tfidf-1000_regi | 0 to 730 days | mean | nan | False | 0.00 | 41541 | 0.00 |
| **688** | tfidf-1000_relevant | 0 to 730 days | mean | nan | False | 0.01 | 132793 | 0.00 |
| **689** | tfidf-1000_rent | 0 to 730 days | mean | nan | False | 0.00 | 55051 | 0.00 |
| **690** | tfidf-1000_resten | 0 to 730 days | mean | nan | False | 0.00 | 49268 | 0.00 |
| **691** | tfidf-1000_retur | 0 to 730 days | mean | nan | False | 0.00 | 35269 | 0.00 |
| **692** | tfidf-1000_rigtig | 0 to 730 days | mean | nan | False | 0.01 | 132546 | 0.00 |
| **693** | tfidf-1000_rigtigt | 0 to 730 days | mean | nan | False | 0.00 | 63247 | 0.00 |
| **694** | tfidf-1000_ringe | 0 to 730 days | mean | nan | False | 0.00 | 90872 | 0.00 |
| **695** | tfidf-1000_ringer | 0 to 730 days | mean | nan | False | 0.00 | 98333 | 0.00 |
| **696** | tfidf-1000_ringet | 0 to 730 days | mean | nan | False | 0.00 | 59182 | 0.00 |

| | Feature name | Lookbehind period | Resolve multiple | Fallback strategy | Static | Mean | N. unique | Proportion using fallback |
|---|---|---|---|---|---|---|---|---|
| **697** | tfidf-1000_rolig | 0 to 730 days | mean | nan | False | 0.00 | 87006 | 0.00 |
| **698** | tfidf-1000_roligt | 0 to 730 days | mean | nan | False | 0.00 | 48421 | 0.00 |
| **699** | tfidf-1000_ryge | 0 to 730 days | mean | nan | False | 0.00 | 31616 | 0.00 |
| **700** | tfidf-1000_ryger | 0 to 730 days | mean | nan | False | 0.00 | 30009 | 0.00 |
| **701** | tfidf-1000_rygning | 0 to 730 days | mean | nan | False | 0.00 | 11483 | 0.00 |
| **702** | tfidf-1000_sagde | 0 to 730 days | mean | nan | False | 0.00 | 45617 | 0.00 |
| **703** | tfidf-1000_sagsbehandler | 0 to 730 days | mean | nan | False | 0.00 | 74629 | 0.00 |
| **704** | tfidf-1000_samarbejde | 0 to 730 days | mean | nan | False | 0.00 | 49919 | 0.00 |
| **705** | tfidf-1000_samlet | 0 to 730 days | mean | nan | False | 0.01 | 116870 | 0.00 |
| **706** | tfidf-1000_samlet relevant | 0 to 730 days | mean | nan | False | 0.00 | 45568 | 0.00 |
| **707** | tfidf-1000_samt | 0 to 730 days | mean | nan | False | 0.01 | 174436 | 0.00 |
| **708** | tfidf-1000_samtale | 0 to 730 days | mean | nan | False | 0.04 | 196690 | 0.00 |
| **709** | tfidf-1000_samtale pt | 0 to 730 days | mean | nan | False | 0.00 | 67004 | 0.00 |
| **710** | tfidf-1000_samtale ut | 0 to 730 days | mean | nan | False | 0.00 | 47815 | 0.00 |
| **711** | tfidf-1000_samtalen | 0 to 730 days | mean | nan | False | 0.02 | 184673 | 0.00 |
| **712** | tfidf-1000_samtalen pt | 0 to 730 days | mean | nan | False | 0.00 | 54296 | 0.00 |
| **713** | tfidf-1000_samtaler | 0 to 730 days | mean | nan | False | 0.01 | 118801 | 0.00 |
| **714** | tfidf-1000_samtidig | 0 to 730 days | mean | nan | False | 0.00 | 104842 | 0.00 |

| | Feature name | Lookbehind period | Resolve multiple | Fallback strategy | Static | Mean | N. unique | Proportion using fallback |
|---|---|---|---|---|---|---|---|---|
| **715** | tfidf-1000_samvær | 0 to 730 days | mean | nan | False | 0.00 | 56273 | 0.00 |
| **716** | tfidf-1000_sat | 0 to 730 days | mean | nan | False | 0.00 | 79502 | 0.00 |
| **717** | tfidf-1000_selvmord | 0 to 730 days | mean | nan | False | 0.00 | 58931 | 0.00 |
| **718** | tfidf-1000_selvmordstanker | 0 to 730 days | mean | nan | False | 0.01 | 116438 | 0.00 |
| **719** | tfidf-1000_selvskade | 0 to 730 days | mean | nan | False | 0.00 | 30276 | 0.00 |
| **720** | tfidf-1000_selvskadende | 0 to 730 days | mean | nan | False | 0.00 | 40449 | 0.00 |
| **721** | tfidf-1000_sendes | 0 to 730 days | mean | nan | False | 0.00 | 47931 | 0.00 |
| **722** | tfidf-1000_sendt | 0 to 730 days | mean | nan | False | 0.00 | 69395 | 0.00 |
| **723** | tfidf-1000_senest | 0 to 730 days | mean | nan | False | 0.00 | 42831 | 0.00 |
| **724** | tfidf-1000_seneste | 0 to 730 days | mean | nan | False | 0.00 | 97140 | 0.00 |
| **725** | tfidf-1000_seng | 0 to 730 days | mean | nan | False | 0.00 | 75857 | 0.00 |
| **726** | tfidf-1000_sengen | 0 to 730 days | mean | nan | False | 0.00 | 63000 | 0.00 |
| **727** | tfidf-1000_seroquel | 0 to 730 days | mean | nan | False | 0.00 | 38282 | 0.00 |
| **728** | tfidf-1000_sertralin | 0 to 730 days | mean | nan | False | 0.00 | 39829 | 0.00 |
| **729** | tfidf-1000_sidde | 0 to 730 days | mean | nan | False | 0.00 | 78095 | 0.00 |
| **730** | tfidf-1000_sidder | 0 to 730 days | mean | nan | False | 0.00 | 110123 | 0.00 |
| **731** | tfidf-1000_siddet | 0 to 730 days | mean | nan | False | 0.00 | 36895 | 0.00 |
| **732** | tfidf-1000_side | 0 to 730 days | mean | nan | False | 0.00 | 87181 | 0.00 |

| | Feature name | Lookbehind period | Resolve multiple | Fallback strategy | Static | Mean | N. unique | Proportion using fallback |
|---|---|---|---|---|---|---|---|---|
| **733** | tfidf-1000_sidst | 0 to 730 days | mean | nan | False | 0.01 | 125209 | 0.00 |
| **734** | tfidf-1000_sidste | 0 to 730 days | mean | nan | False | 0.01 | 182382 | 0.00 |
| **735** | tfidf-1000_sidste samtale | 0 to 730 days | mean | nan | False | 0.00 | 73070 | 0.00 |
| **736** | tfidf-1000_sidste uge | 0 to 730 days | mean | nan | False | 0.00 | 79864 | 0.00 |
| **737** | tfidf-1000_sikker | 0 to 730 days | mean | nan | False | 0.00 | 52090 | 0.00 |
| **738** | tfidf-1000_situation | 0 to 730 days | mean | nan | False | 0.00 | 114743 | 0.00 |
| **739** | tfidf-1000_situationen | 0 to 730 days | mean | nan | False | 0.00 | 80648 | 0.00 |
| **740** | tfidf-1000_situationer | 0 to 730 days | mean | nan | False | 0.00 | 91113 | 0.00 |
| **741** | tfidf-1000_skade | 0 to 730 days | mean | nan | False | 0.00 | 56722 | 0.00 |
| **742** | tfidf-1000_sket | 0 to 730 days | mean | nan | False | 0.00 | 73295 | 0.00 |
| **743** | tfidf-1000_skidt | 0 to 730 days | mean | nan | False | 0.00 | 69236 | 0.00 |
| **744** | tfidf-1000_skizofreni | 0 to 730 days | mean | nan | False | 0.00 | 11119 | 0.00 |
| **745** | tfidf-1000_skole | 0 to 730 days | mean | nan | False | 0.00 | 76432 | 0.00 |
| **746** | tfidf-1000_skolen | 0 to 730 days | mean | nan | False | 0.00 | 78293 | 0.00 |
| **747** | tfidf-1000_skrevet | 0 to 730 days | mean | nan | False | 0.00 | 61765 | 0.00 |
| **748** | tfidf-1000_skrive | 0 to 730 days | mean | nan | False | 0.00 | 61245 | 0.00 |
| **749** | tfidf-1000_skyld | 0 to 730 days | mean | nan | False | 0.00 | 53092 | 0.00 |
| **750** | tfidf-1000_skyldes | 0 to 730 days | mean | nan | False | 0.00 | 75289 | 0.00 |

| | Feature name | Lookbehind period | Resolve multiple | Fallback strategy | Static | Mean | N. unique | Proportion using fallback |
|---|---|---|---|---|---|---|---|---|
| **751** | tfidf-1000_slet | 0 to 730 days | mean | nan | False | 0.00 | 78387 | 0.00 |
| **752** | tfidf-1000_slå | 0 to 730 days | mean | nan | False | 0.00 | 47268 | 0.00 |
| **753** | tfidf-1000_smerter | 0 to 730 days | mean | nan | False | 0.01 | 78098 | 0.00 |
| **754** | tfidf-1000_smil | 0 to 730 days | mean | nan | False | 0.00 | 51994 | 0.00 |
| **755** | tfidf-1000_smilende | 0 to 730 days | mean | nan | False | 0.00 | 77742 | 0.00 |
| **756** | tfidf-1000_smiler | 0 to 730 days | mean | nan | False | 0.00 | 39551 | 0.00 |
| **757** | tfidf-1000_sms | 0 to 730 days | mean | nan | False | 0.00 | 51074 | 0.00 |
| **758** | tfidf-1000_smule | 0 to 730 days | mean | nan | False | 0.00 | 66177 | 0.00 |
| **759** | tfidf-1000_snak | 0 to 730 days | mean | nan | False | 0.00 | 58666 | 0.00 |
| **760** | tfidf-1000_snakke | 0 to 730 days | mean | nan | False | 0.00 | 73545 | 0.00 |
| **761** | tfidf-1000_snakker | 0 to 730 days | mean | nan | False | 0.00 | 73313 | 0.00 |
| **762** | tfidf-1000_snakket | 0 to 730 days | mean | nan | False | 0.00 | 47455 | 0.00 |
| **763** | tfidf-1000_social | 0 to 730 days | mean | nan | False | 0.00 | 92390 | 0.00 |
| **764** | tfidf-1000_sociale | 0 to 730 days | mean | nan | False | 0.00 | 105907 | 0.00 |
| **765** | tfidf-1000_socialt | 0 to 730 days | mean | nan | False | 0.00 | 70908 | 0.00 |
| **766** | tfidf-1000_somatisk | 0 to 730 days | mean | nan | False | 0.00 | 50072 | 0.00 |
| **767** | tfidf-1000_somatiske | 0 to 730 days | mean | nan | False | 0.00 | 44731 | 0.00 |
| **768** | tfidf-1000_sov | 0 to 730 days | mean | nan | False | 0.00 | 36775 | 0.00 |

| | Feature name | Lookbehind period | Resolve multiple | Fallback strategy | Static | Mean | N. unique | Proportion using fallback |
|---|---|---|---|---|---|---|---|---|
| **769** | tfidf-1000_sove | 0 to 730 days | mean | nan | False | 0.00 | 110815 | 0.00 |
| **770** | tfidf-1000_sover | 0 to 730 days | mean | nan | False | 0.00 | 129456 | 0.00 |
| **771** | tfidf-1000_sovet | 0 to 730 days | mean | nan | False | 0.00 | 81404 | 0.00 |
| **772** | tfidf-1000_sparsom | 0 to 730 days | mean | nan | False | 0.00 | 39652 | 0.00 |
| **773** | tfidf-1000_specielt | 0 to 730 days | mean | nan | False | 0.00 | 66153 | 0.00 |
| **774** | tfidf-1000_spil | 0 to 730 days | mean | nan | False | 0.00 | 33410 | 0.00 |
| **775** | tfidf-1000_spille | 0 to 730 days | mean | nan | False | 0.00 | 38410 | 0.00 |
| **776** | tfidf-1000_spiller | 0 to 730 days | mean | nan | False | 0.00 | 38735 | 0.00 |
| **777** | tfidf-1000_spillet | 0 to 730 days | mean | nan | False | 0.00 | 22916 | 0.00 |
| **778** | tfidf-1000_spise | 0 to 730 days | mean | nan | False | 0.00 | 81851 | 0.00 |
| **779** | tfidf-1000_spiser | 0 to 730 days | mean | nan | False | 0.00 | 81405 | 0.00 |
| **780** | tfidf-1000_spist | 0 to 730 days | mean | nan | False | 0.00 | 48432 | 0.00 |
| **781** | tfidf-1000_spontant | 0 to 730 days | mean | nan | False | 0.00 | 41437 | 0.00 |
| **782** | tfidf-1000_spurgt | 0 to 730 days | mean | nan | False | 0.00 | 55404 | 0.00 |
| **783** | tfidf-1000_spørge | 0 to 730 days | mean | nan | False | 0.00 | 47932 | 0.00 |
| **784** | tfidf-1000_spørger | 0 to 730 days | mean | nan | False | 0.00 | 102443 | 0.00 |
| **785** | tfidf-1000_spørges | 0 to 730 days | mean | nan | False | 0.00 | 49941 | 0.00 |
| **786** | tfidf-1000_spørgsmål | 0 to 730 days | mean | nan | False | 0.00 | 107775 | 0.00 |

| | Feature name | Lookbehind period | Resolve multiple | Fallback strategy | Static | Mean | N. unique | Proportion using fallback |
|---|---|---|---|---|---|---|---|---|
| **787** | tfidf-1000_stabil | 0 to 730 days | mean | nan | False | 0.00 | 47398 | 0.00 |
| **788** | tfidf-1000_stand | 0 to 730 days | mean | nan | False | 0.00 | 78553 | 0.00 |
| **789** | tfidf-1000_start | 0 to 730 days | mean | nan | False | 0.00 | 53963 | 0.00 |
| **790** | tfidf-1000_starte | 0 to 730 days | mean | nan | False | 0.00 | 97506 | 0.00 |
| **791** | tfidf-1000_starten | 0 to 730 days | mean | nan | False | 0.00 | 78502 | 0.00 |
| **792** | tfidf-1000_starter | 0 to 730 days | mean | nan | False | 0.00 | 77485 | 0.00 |
| **793** | tfidf-1000_startet | 0 to 730 days | mean | nan | False | 0.00 | 77096 | 0.00 |
| **794** | tfidf-1000_sted | 0 to 730 days | mean | nan | False | 0.00 | 102945 | 0.00 |
| **795** | tfidf-1000_stede | 0 to 730 days | mean | nan | False | 0.00 | 80618 | 0.00 |
| **796** | tfidf-1000_steder | 0 to 730 days | mean | nan | False | 0.00 | 58104 | 0.00 |
| **797** | tfidf-1000_stedet | 0 to 730 days | mean | nan | False | 0.00 | 87920 | 0.00 |
| **798** | tfidf-1000_stemme | 0 to 730 days | mean | nan | False | 0.00 | 35249 | 0.00 |
| **799** | tfidf-1000_stemmehøring | 0 to 730 days | mean | nan | False | 0.00 | 21804 | 0.00 |
| **800** | tfidf-1000_stemmer | 0 to 730 days | mean | nan | False | 0.00 | 37070 | 0.00 |
| **801** | tfidf-1000_stemmerne | 0 to 730 days | mean | nan | False | 0.00 | 13021 | 0.00 |
| **802** | tfidf-1000_stemningsleje | 0 to 730 days | mean | nan | False | 0.01 | 134836 | 0.00 |
| **803** | tfidf-1000_stemningslejet | 0 to 730 days | mean | nan | False | 0.01 | 101018 | 0.00 |
| **804** | tfidf-1000_stille | 0 to 730 days | mean | nan | False | 0.00 | 109949 | 0.00 |

| | Feature name | Lookbehind period | Resolve multiple | Fallback strategy | Static | Mean | N. unique | Proportion using fallback |
|---|---|---|---|---|---|---|---|---|
| **805** | tfidf-1000_stille rolig | 0 to 730 days | mean | nan | False | 0.00 | 28668 | 0.00 |
| **806** | tfidf-1000_stilling | 0 to 730 days | mean | nan | False | 0.00 | 56511 | 0.00 |
| **807** | tfidf-1000_stod | 0 to 730 days | mean | nan | False | 0.00 | 34473 | 0.00 |
| **808** | tfidf-1000_stoffer | 0 to 730 days | mean | nan | False | 0.00 | 28101 | 0.00 |
| **809** | tfidf-1000_stoppe | 0 to 730 days | mean | nan | False | 0.00 | 61329 | 0.00 |
| **810** | tfidf-1000_stresset | 0 to 730 days | mean | nan | False | 0.00 | 70977 | 0.00 |
| **811** | tfidf-1000_struktur | 0 to 730 days | mean | nan | False | 0.00 | 68903 | 0.00 |
| **812** | tfidf-1000_stue | 0 to 730 days | mean | nan | False | 0.00 | 36008 | 0.00 |
| **813** | tfidf-1000_stuen | 0 to 730 days | mean | nan | False | 0.00 | 45122 | 0.00 |
| **814** | tfidf-1000_styr | 0 to 730 days | mean | nan | False | 0.00 | 58556 | 0.00 |
| **815** | tfidf-1000_stå | 0 to 730 days | mean | nan | False | 0.00 | 86444 | 0.00 |
| **816** | tfidf-1000_står | 0 to 730 days | mean | nan | False | 0.00 | 103481 | 0.00 |
| **817** | tfidf-1000_større | 0 to 730 days | mean | nan | False | 0.00 | 74458 | 0.00 |
| **818** | tfidf-1000_støtte | 0 to 730 days | mean | nan | False | 0.00 | 111503 | 0.00 |
| **819** | tfidf-1000_suicidal | 0 to 730 days | mean | nan | False | 0.00 | 51200 | 0.00 |
| **820** | tfidf-1000_suicidale | 0 to 730 days | mean | nan | False | 0.00 | 48095 | 0.00 |
| **821** | tfidf-1000_suicidaltanker | 0 to 730 days | mean | nan | False | 0.00 | 44539 | 0.00 |
| **822** | tfidf-1000_suicidaltruet | 0 to 730 days | mean | nan | False | 0.00 | 42769 | 0.00 |

| | Feature name | Lookbehind period | Resolve multiple | Fallback strategy | Static | Mean | N. unique | Proportion using fallback |
|---|---|---|---|---|---|---|---|---|
| **823** | tfidf-1000_svar | 0 to 730 days | mean | nan | False | 0.00 | 78540 | 0.00 |
| **824** | tfidf-1000_svare | 0 to 730 days | mean | nan | False | 0.00 | 54509 | 0.00 |
| **825** | tfidf-1000_svarer | 0 to 730 days | mean | nan | False | 0.01 | 121861 | 0.00 |
| **826** | tfidf-1000_svarer relevant | 0 to 730 days | mean | nan | False | 0.00 | 52205 | 0.00 |
| **827** | tfidf-1000_svingende | 0 to 730 days | mean | nan | False | 0.00 | 69694 | 0.00 |
| **828** | tfidf-1000_svinger | 0 to 730 days | mean | nan | False | 0.00 | 52563 | 0.00 |
| **829** | tfidf-1000_svær | 0 to 730 days | mean | nan | False | 0.00 | 94429 | 0.00 |
| **830** | tfidf-1000_svære | 0 to 730 days | mean | nan | False | 0.00 | 73698 | 0.00 |
| **831** | tfidf-1000_svært | 0 to 730 days | mean | nan | False | 0.02 | 197894 | 0.00 |
| **832** | tfidf-1000_syg | 0 to 730 days | mean | nan | False | 0.00 | 80326 | 0.00 |
| **833** | tfidf-1000_sygdom | 0 to 730 days | mean | nan | False | 0.00 | 70854 | 0.00 |
| **834** | tfidf-1000_sygemeldt | 0 to 730 days | mean | nan | False | 0.00 | 77797 | 0.00 |
| **835** | tfidf-1000_sygeplejerske | 0 to 730 days | mean | nan | False | 0.00 | 50257 | 0.00 |
| **836** | tfidf-1000_symptomer | 0 to 730 days | mean | nan | False | 0.01 | 174068 | 0.00 |
| **837** | tfidf-1000_såfremt | 0 to 730 days | mean | nan | False | 0.00 | 70866 | 0.00 |
| **838** | tfidf-1000_særligt | 0 to 730 days | mean | nan | False | 0.00 | 80125 | 0.00 |
| **839** | tfidf-1000_sætte | 0 to 730 days | mean | nan | False | 0.00 | 81138 | 0.00 |
| **840** | tfidf-1000_sætter | 0 to 730 days | mean | nan | False | 0.00 | 54159 | 0.00 |

| | Feature name | Lookbehind period | Resolve multiple | Fallback strategy | Static | Mean | N. unique | Proportion using fallback |
|---|---|---|---|---|---|---|---|---|
| **841** | tfidf-1000_søge | 0 to 730 days | mean | nan | False | 0.00 | 64489 | 0.00 |
| **842** | tfidf-1000_søn | 0 to 730 days | mean | nan | False | 0.00 | 53874 | 0.00 |
| **843** | tfidf-1000_søndag | 0 to 730 days | mean | nan | False | 0.00 | 39001 | 0.00 |
| **844** | tfidf-1000_sønnen | 0 to 730 days | mean | nan | False | 0.00 | 33839 | 0.00 |
| **845** | tfidf-1000_søster | 0 to 730 days | mean | nan | False | 0.00 | 58037 | 0.00 |
| **846** | tfidf-1000_søvn | 0 to 730 days | mean | nan | False | 0.00 | 125891 | 0.00 |
| **847** | tfidf-1000_søvnen | 0 to 730 days | mean | nan | False | 0.00 | 46065 | 0.00 |
| **848** | tfidf-1000_tabt | 0 to 730 days | mean | nan | False | 0.00 | 54368 | 0.00 |
| **849** | tfidf-1000_tager | 0 to 730 days | mean | nan | False | 0.01 | 158343 | 0.00 |
| **850** | tfidf-1000_tages | 0 to 730 days | mean | nan | False | 0.00 | 66000 | 0.00 |
| **851** | tfidf-1000_taget | 0 to 730 days | mean | nan | False | 0.01 | 150998 | 0.00 |
| **852** | tfidf-1000_tale | 0 to 730 days | mean | nan | False | 0.01 | 155890 | 0.00 |
| **853** | tfidf-1000_talende | 0 to 730 days | mean | nan | False | 0.00 | 47543 | 0.00 |
| **854** | tfidf-1000_talepres | 0 to 730 days | mean | nan | False | 0.00 | 20662 | 0.00 |
| **855** | tfidf-1000_taler | 0 to 730 days | mean | nan | False | 0.01 | 159764 | 0.00 |
| **856** | tfidf-1000_taler pt | 0 to 730 days | mean | nan | False | 0.00 | 35553 | 0.00 |
| **857** | tfidf-1000_tales | 0 to 730 days | mean | nan | False | 0.00 | 51040 | 0.00 |
| **858** | tfidf-1000_talt | 0 to 730 days | mean | nan | False | 0.00 | 113763 | 0.00 |

| | Feature name | Lookbehind period | Resolve multiple | Fallback strategy | Static | Mean | N. unique | Proportion using fallback |
|---|---|---|---|---|---|---|---|---|
| **859** | tfidf-1000_tanke | 0 to 730 days | mean | nan | False | 0.00 | 54284 | 0.00 |
| **860** | tfidf-1000_tankegang | 0 to 730 days | mean | nan | False | 0.00 | 38155 | 0.00 |
| **861** | tfidf-1000_tankemylder | 0 to 730 days | mean | nan | False | 0.00 | 62942 | 0.00 |
| **862** | tfidf-1000_tanker | 0 to 730 days | mean | nan | False | 0.01 | 169611 | 0.00 |
| **863** | tfidf-1000_tankerne | 0 to 730 days | mean | nan | False | 0.00 | 92001 | 0.00 |
| **864** | tfidf-1000_tbl | 0 to 730 days | mean | nan | False | 0.00 | 46288 | 0.00 |
| **865** | tfidf-1000_tegn | 0 to 730 days | mean | nan | False | 0.00 | 82189 | 0.00 |
| **866** | tfidf-1000_telefon | 0 to 730 days | mean | nan | False | 0.00 | 53304 | 0.00 |
| **867** | tfidf-1000_telefonen | 0 to 730 days | mean | nan | False | 0.00 | 38658 | 0.00 |
| **868** | tfidf-1000_telefonisk | 0 to 730 days | mean | nan | False | 0.00 | 65357 | 0.00 |
| **869** | tfidf-1000_tempo | 0 to 730 days | mean | nan | False | 0.00 | 82854 | 0.00 |
| **870** | tfidf-1000_tendens | 0 to 730 days | mean | nan | False | 0.00 | 102835 | 0.00 |
| **871** | tfidf-1000_thomas | 0 to 730 days | mean | nan | False | 0.00 | 9238 | 0.00 |
| **872** | tfidf-1000_tid | 0 to 730 days | mean | nan | False | 0.03 | 197541 | 0.00 |
| **873** | tfidf-1000_tid sted | 0 to 730 days | mean | nan | False | 0.00 | 47762 | 0.00 |
| **874** | tfidf-1000_tiden | 0 to 730 days | mean | nan | False | 0.01 | 156986 | 0.00 |
| **875** | tfidf-1000_tider | 0 to 730 days | mean | nan | False | 0.00 | 71277 | 0.00 |
| **876** | tfidf-1000_tidl | 0 to 730 days | mean | nan | False | 0.00 | 74080 | 0.00 |

| | Feature name | Lookbehind period | Resolve multiple | Fallback strategy | Static | Mean | N. unique | Proportion using fallback |
|---|---|---|---|---|---|---|---|---|
| **877** | tfidf-1000_tidligt | 0 to 730 days | mean | nan | False | 0.00 | 45527 | 0.00 |
| **878** | tfidf-1000_tidspunkt | 0 to 730 days | mean | nan | False | 0.00 | 104732 | 0.00 |
| **879** | tfidf-1000_tilbagemelding | 0 to 730 days | mean | nan | False | 0.00 | 54221 | 0.00 |
| **880** | tfidf-1000_tilbud | 0 to 730 days | mean | nan | False | 0.00 | 67373 | 0.00 |
| **881** | tfidf-1000_tilbudt | 0 to 730 days | mean | nan | False | 0.00 | 63307 | 0.00 |
| **882** | tfidf-1000_tilbydes | 0 to 730 days | mean | nan | False | 0.00 | 69701 | 0.00 |
| **883** | tfidf-1000_tilfreds | 0 to 730 days | mean | nan | False | 0.00 | 48240 | 0.00 |
| **884** | tfidf-1000_tilknyttet | 0 to 730 days | mean | nan | False | 0.00 | 61666 | 0.00 |
| **885** | tfidf-1000_tilstand | 0 to 730 days | mean | nan | False | 0.01 | 124847 | 0.00 |
| **886** | tfidf-1000_tilstanden | 0 to 730 days | mean | nan | False | 0.00 | 63129 | 0.00 |
| **887** | tfidf-1000_tilstede | 0 to 730 days | mean | nan | False | 0.01 | 83364 | 0.00 |
| **888** | tfidf-1000_tilstede pt | 0 to 730 days | mean | nan | False | 0.00 | 31786 | 0.00 |
| **889** | tfidf-1000_tilsyn | 0 to 730 days | mean | nan | False | 0.00 | 30651 | 0.00 |
| **890** | tfidf-1000_tiltagende | 0 to 730 days | mean | nan | False | 0.00 | 96328 | 0.00 |
| **891** | tfidf-1000_time | 0 to 730 days | mean | nan | False | 0.00 | 86248 | 0.00 |
| **892** | tfidf-1000_timer | 0 to 730 days | mean | nan | False | 0.01 | 136318 | 0.00 |
| **893** | tfidf-1000_ting | 0 to 730 days | mean | nan | False | 0.01 | 166802 | 0.00 |
| **894** | tfidf-1000_tingene | 0 to 730 days | mean | nan | False | 0.00 | 97765 | 0.00 |

| | Feature name | Lookbehind period | Resolve multiple | Fallback strategy | Static | Mean | N. unique | Proportion using fallback |
|---|---|---|---|---|---|---|---|---|
| **895** | tfidf-1000_tirsdag | 0 to 730 days | mean | nan | False | 0.00 | 73049 | 0.00 |
| **896** | tfidf-1000_tlf | 0 to 730 days | mean | nan | False | 0.00 | 57907 | 0.00 |
| **897** | tfidf-1000_tog | 0 to 730 days | mean | nan | False | 0.00 | 85239 | 0.00 |
| **898** | tfidf-1000_torsdag | 0 to 730 days | mean | nan | False | 0.00 | 76781 | 0.00 |
| **899** | tfidf-1000_trist | 0 to 730 days | mean | nan | False | 0.01 | 126520 | 0.00 |
| **900** | tfidf-1000_tristhed | 0 to 730 days | mean | nan | False | 0.00 | 70437 | 0.00 |
| **901** | tfidf-1000_trods | 0 to 730 days | mean | nan | False | 0.00 | 90735 | 0.00 |
| **902** | tfidf-1000_tror | 0 to 730 days | mean | nan | False | 0.00 | 88458 | 0.00 |
| **903** | tfidf-1000_truende | 0 to 730 days | mean | nan | False | 0.00 | 16786 | 0.00 |
| **904** | tfidf-1000_tryg | 0 to 730 days | mean | nan | False | 0.00 | 55857 | 0.00 |
| **905** | tfidf-1000_trække | 0 to 730 days | mean | nan | False | 0.00 | 53455 | 0.00 |
| **906** | tfidf-1000_trækker | 0 to 730 days | mean | nan | False | 0.00 | 52145 | 0.00 |
| **907** | tfidf-1000_træt | 0 to 730 days | mean | nan | False | 0.01 | 142516 | 0.00 |
| **908** | tfidf-1000_træthed | 0 to 730 days | mean | nan | False | 0.00 | 68946 | 0.00 |
| **909** | tfidf-1000_tur | 0 to 730 days | mean | nan | False | 0.00 | 79489 | 0.00 |
| **910** | tfidf-1000_ture | 0 to 730 days | mean | nan | False | 0.00 | 41322 | 0.00 |
| **911** | tfidf-1000_tv | 0 to 730 days | mean | nan | False | 0.00 | 67651 | 0.00 |
| **912** | tfidf-1000_tvivl | 0 to 730 days | mean | nan | False | 0.00 | 82806 | 0.00 |

| | Feature name | Lookbehind period | Resolve multiple | Fallback strategy | Static | Mean | N. unique | Proportion using fallback |
|---|---|---|---|---|---|---|---|---|
| **913** | tfidf-1000_tydeligt | 0 to 730 days | mean | nan | False | 0.00 | 85355 | 0.00 |
| **914** | tfidf-1000_tænke | 0 to 730 days | mean | nan | False | 0.00 | 107906 | 0.00 |
| **915** | tfidf-1000_tænker | 0 to 730 days | mean | nan | False | 0.01 | 136520 | 0.00 |
| **916** | tfidf-1000_tænkt | 0 to 730 days | mean | nan | False | 0.00 | 68983 | 0.00 |
| **917** | tfidf-1000_tøj | 0 to 730 days | mean | nan | False | 0.00 | 53799 | 0.00 |
| **918** | tfidf-1000_uddannelse | 0 to 730 days | mean | nan | False | 0.00 | 62007 | 0.00 |
| **919** | tfidf-1000_udenfor | 0 to 730 days | mean | nan | False | 0.00 | 65549 | 0.00 |
| **920** | tfidf-1000_udgang | 0 to 730 days | mean | nan | False | 0.00 | 24916 | 0.00 |
| **921** | tfidf-1000_udleveret | 0 to 730 days | mean | nan | False | 0.00 | 58109 | 0.00 |
| **922** | tfidf-1000_udredning | 0 to 730 days | mean | nan | False | 0.01 | 96902 | 0.00 |
| **923** | tfidf-1000_udskrevet | 0 to 730 days | mean | nan | False | 0.00 | 37830 | 0.00 |
| **924** | tfidf-1000_udskrivelse | 0 to 730 days | mean | nan | False | 0.00 | 35143 | 0.00 |
| **925** | tfidf-1000_udskrives | 0 to 730 days | mean | nan | False | 0.00 | 34043 | 0.00 |
| **926** | tfidf-1000_udtalt | 0 to 730 days | mean | nan | False | 0.00 | 53984 | 0.00 |
| **927** | tfidf-1000_udtryk | 0 to 730 days | mean | nan | False | 0.01 | 127075 | 0.00 |
| **928** | tfidf-1000_udtrykker | 0 to 730 days | mean | nan | False | 0.00 | 66049 | 0.00 |
| **929** | tfidf-1000_uge | 0 to 730 days | mean | nan | False | 0.01 | 157299 | 0.00 |
| **930** | tfidf-1000_ugen | 0 to 730 days | mean | nan | False | 0.00 | 108658 | 0.00 |

| | Feature name | Lookbehind period | Resolve multiple | Fallback strategy | Static | Mean | N. unique | Proportion using fallback |
|---|---|---|---|---|---|---|---|---|
| **931** | tfidf-1000_ugentligt | 0 to 730 days | mean | nan | False | 0.00 | 54469 | 0.00 |
| **932** | tfidf-1000_uger | 0 to 730 days | mean | nan | False | 0.01 | 124424 | 0.00 |
| **933** | tfidf-1000_umiddelbart | 0 to 730 days | mean | nan | False | 0.00 | 96351 | 0.00 |
| **934** | tfidf-1000_undersøgelse | 0 to 730 days | mean | nan | False | 0.00 | 51924 | 0.00 |
| **935** | tfidf-1000_undertegnede | 0 to 730 days | mean | nan | False | 0.01 | 71368 | 0.00 |
| **936** | tfidf-1000_undgå | 0 to 730 days | mean | nan | False | 0.00 | 68728 | 0.00 |
| **937** | tfidf-1000_upåfaldende | 0 to 730 days | mean | nan | False | 0.00 | 54110 | 0.00 |
| **938** | tfidf-1000_uro | 0 to 730 days | mean | nan | False | 0.00 | 111593 | 0.00 |
| **939** | tfidf-1000_urolig | 0 to 730 days | mean | nan | False | 0.00 | 75231 | 0.00 |
| **940** | tfidf-1000_usikker | 0 to 730 days | mean | nan | False | 0.00 | 72867 | 0.00 |
| **941** | tfidf-1000_ut | 0 to 730 days | mean | nan | False | 0.02 | 169421 | 0.00 |
| **942** | tfidf-1000_ut pt | 0 to 730 days | mean | nan | False | 0.00 | 38766 | 0.00 |
| **943** | tfidf-1000_ut spørger | 0 to 730 days | mean | nan | False | 0.00 | 33602 | 0.00 |
| **944** | tfidf-1000_uændret | 0 to 730 days | mean | nan | False | 0.00 | 67076 | 0.00 |
| **945** | tfidf-1000_vagt | 0 to 730 days | mean | nan | False | 0.00 | 32530 | 0.00 |
| **946** | tfidf-1000_vagten | 0 to 730 days | mean | nan | False | 0.00 | 31065 | 0.00 |
| **947** | tfidf-1000_vand | 0 to 730 days | mean | nan | False | 0.00 | 30272 | 0.00 |
| **948** | tfidf-1000_vanlig | 0 to 730 days | mean | nan | False | 0.00 | 31735 | 0.00 |

| | Feature name | Lookbehind period | Resolve multiple | Fallback strategy | Static | Mean | N. unique | Proportion using fallback |
|---|---|---|---|---|---|---|---|---|
| **949** | tfidf-1000_vanligt | 0 to 730 days | mean | nan | False | 0.00 | 46007 | 0.00 |
| **950** | tfidf-1000_vanskeligheder | 0 to 730 days | mean | nan | False | 0.00 | 91204 | 0.00 |
| **951** | tfidf-1000_vanskeligt | 0 to 730 days | mean | nan | False | 0.00 | 65657 | 0.00 |
| **952** | tfidf-1000_vedr | 0 to 730 days | mean | nan | False | 0.00 | 96729 | 0.00 |
| **953** | tfidf-1000_vej | 0 to 730 days | mean | nan | False | 0.00 | 68177 | 0.00 |
| **954** | tfidf-1000_velbefindende | 0 to 730 days | mean | nan | False | 0.00 | 41378 | 0.00 |
| **955** | tfidf-1000_ven | 0 to 730 days | mean | nan | False | 0.00 | 45986 | 0.00 |
| **956** | tfidf-1000_vender | 0 to 730 days | mean | nan | False | 0.00 | 48141 | 0.00 |
| **957** | tfidf-1000_veninde | 0 to 730 days | mean | nan | False | 0.00 | 73304 | 0.00 |
| **958** | tfidf-1000_veninder | 0 to 730 days | mean | nan | False | 0.00 | 56091 | 0.00 |
| **959** | tfidf-1000_venlig | 0 to 730 days | mean | nan | False | 0.00 | 84961 | 0.00 |
| **960** | tfidf-1000_venlig imødekommende | 0 to 730 days | mean | nan | False | 0.00 | 43627 | 0.00 |
| **961** | tfidf-1000_venlig kontakten | 0 to 730 days | mean | nan | False | 0.00 | 31172 | 0.00 |
| **962** | tfidf-1000_venner | 0 to 730 days | mean | nan | False | 0.00 | 111603 | 0.00 |
| **963** | tfidf-1000_vente | 0 to 730 days | mean | nan | False | 0.00 | 53951 | 0.00 |
| **964** | tfidf-1000_vide | 0 to 730 days | mean | nan | False | 0.00 | 88166 | 0.00 |
| **965** | tfidf-1000_vigtigt | 0 to 730 days | mean | nan | False | 0.00 | 70635 | 0.00 |
| **966** | tfidf-1000_virker | 0 to 730 days | mean | nan | False | 0.01 | 147213 | 0.00 |

| | Feature name | Lookbehind period | Resolve multiple | Fallback strategy | Static | Mean | N. unique | Proportion using fallback |
|---|---|---|---|---|---|---|---|---|
| **967** | tfidf-1000_virket | 0 to 730 days | mean | nan | False | 0.00 | 22866 | 0.00 |
| **968** | tfidf-1000_virkning | 0 to 730 days | mean | nan | False | 0.00 | 49317 | 0.00 |
| **969** | tfidf-1000_viser | 0 to 730 days | mean | nan | False | 0.00 | 89229 | 0.00 |
| **970** | tfidf-1000_vko | 0 to 730 days | mean | nan | False | 0.00 | 46797 | 0.00 |
| **971** | tfidf-1000_voldsomt | 0 to 730 days | mean | nan | False | 0.00 | 58506 | 0.00 |
| **972** | tfidf-1000_vrangforestillinger | 0 to 730 days | mean | nan | False | 0.00 | 54625 | 0.00 |
| **973** | tfidf-1000_vred | 0 to 730 days | mean | nan | False | 0.00 | 85862 | 0.00 |
| **974** | tfidf-1000_vrede | 0 to 730 days | mean | nan | False | 0.00 | 62858 | 0.00 |
| **975** | tfidf-1000_vurdere | 0 to 730 days | mean | nan | False | 0.00 | 55254 | 0.00 |
| **976** | tfidf-1000_vurderer | 0 to 730 days | mean | nan | False | 0.00 | 53385 | 0.00 |
| **977** | tfidf-1000_vurderes | 0 to 730 days | mean | nan | False | 0.01 | 135726 | 0.00 |
| **978** | tfidf-1000_vurderet | 0 to 730 days | mean | nan | False | 0.00 | 47184 | 0.00 |
| **979** | tfidf-1000_vurdering | 0 to 730 days | mean | nan | False | 0.01 | 109340 | 0.00 |
| **980** | tfidf-1000_vågen | 0 to 730 days | mean | nan | False | 0.00 | 114514 | 0.00 |
| **981** | tfidf-1000_vågen klar | 0 to 730 days | mean | nan | False | 0.00 | 92641 | 0.00 |
| **982** | tfidf-1000_vågner | 0 to 730 days | mean | nan | False | 0.00 | 79611 | 0.00 |
| **983** | tfidf-1000_vægt | 0 to 730 days | mean | nan | False | 0.00 | 48849 | 0.00 |
| **984** | tfidf-1000_væk | 0 to 730 days | mean | nan | False | 0.00 | 101320 | 0.00 |

| | Feature name | Lookbehind period | Resolve multiple | Fallback strategy | Static | Mean | N. unique | Proportion using fallback |
|---|---|---|---|---|---|---|---|---|
| **985** | tfidf-1000_vælger | 0 to 730 days | mean | nan | False | 0.00 | 41610 | 0.00 |
| **986** | tfidf-1000_værelse | 0 to 730 days | mean | nan | False | 0.00 | 38679 | 0.00 |
| **987** | tfidf-1000_værelset | 0 to 730 days | mean | nan | False | 0.00 | 26757 | 0.00 |
| **988** | tfidf-1000_værre | 0 to 730 days | mean | nan | False | 0.00 | 63350 | 0.00 |
| **989** | tfidf-1000_weekend | 0 to 730 days | mean | nan | False | 0.00 | 53132 | 0.00 |
| **990** | tfidf-1000_weekenden | 0 to 730 days | mean | nan | False | 0.00 | 74468 | 0.00 |
| **991** | tfidf-1000_yderligere | 0 to 730 days | mean | nan | False | 0.00 | 105191 | 0.00 |
| **992** | tfidf-1000_åben | 0 to 730 days | mean | nan | False | 0.00 | 61562 | 0.00 |
| **993** | tfidf-1000_årig | 0 to 730 days | mean | nan | False | 0.00 | 55017 | 0.00 |
| **994** | tfidf-1000_æf | 0 to 730 days | mean | nan | False | 0.00 | 28049 | 0.00 |
| **995** | tfidf-1000_ægtefælle | 0 to 730 days | mean | nan | False | 0.00 | 38593 | 0.00 |
| **996** | tfidf-1000_ægtefællen | 0 to 730 days | mean | nan | False | 0.00 | 29427 | 0.00 |
| **997** | tfidf-1000_ændret | 0 to 730 days | mean | nan | False | 0.00 | 50739 | 0.00 |
| **998** | tfidf-1000_øget | 0 to 730 days | mean | nan | False | 0.00 | 110053 | 0.00 |
| **999** | tfidf-1000_øjenkontakt | 0 to 730 days | mean | nan | False | 0.01 | 99618 | 0.00 |
| **1000** | tfidf-1000_ønske | 0 to 730 days | mean | nan | False | 0.00 | 113805 | 0.00 |
| **1001** | tfidf-1000_ønskede | 0 to 730 days | mean | nan | False | 0.00 | 40550 | 0.00 |
| **1002** | tfidf-1000_ønsker | 0 to 730 days | mean | nan | False | 0.01 | 158660 | 0.00 |

| | Feature name | Lookbehind period | Resolve multiple | Fallback strategy | Static | Mean | N. unique | Proportion using fallback |
|---|---|---|---|---|---|---|---|---|
| **1003** | tfidf-1000_ønsket | 0 to 730 days | mean | nan | False | 0.00 | 48562 | 0.00 |
| **1004** | antidepressives | 0 to 365 days | count | 0 | False | 3.62 | 378 | 0.88 |
| **1005** | antidepressives | 0 to 183 days | count | 0 | False | 1.95 | 263 | 0.92 |
| **1006** | antidepressives | 0 to 730 days | count | 0 | False | 6.21 | 616 | 0.84 |
| **1007** | antipsychotics | 0 to 730 days | count | 0 | False | 12.87 | 1152 | 0.85 |
| **1008** | antipsychotics | 0 to 365 days | count | 0 | False | 7.57 | 804 | 0.88 |
| **1009** | antipsychotics | 0 to 183 days | count | 0 | False | 4.01 | 594 | 0.92 |
| **1010** | benzodiazepine_related_sleeping_agents | 0 to 365 days | boolean | 0 | False | 0.08 | 2 | 0.92 |
| **1011** | benzodiazepine_related_sleeping_agents | 0 to 183 days | boolean | 0 | False | 0.05 | 2 | 0.95 |
| **1012** | benzodiazepine_related_sleeping_agents | 0 to 730 days | boolean | 0 | False | 0.11 | 2 | 0.89 |
| **1013** | benzodiazepines | 0 to 730 days | boolean | 0 | False | 0.11 | 2 | 0.89 |
| **1014** | benzodiazepines | 0 to 183 days | boolean | 0 | False | 0.05 | 2 | 0.95 |
| **1015** | benzodiazepines | 0 to 365 days | boolean | 0 | False | 0.08 | 2 | 0.92 |
| **1016** | broeset_violence_checklis4 | 0 to 730 days | latest | nan | False | 0.11 | 8 | 0.00 |
| **1017** | broeset_violence_checklist | 0 to 183 days | latest | nan | False | 0.12 | 8 | 0.00 |
| **1018** | broeset_violence_checklist | 0 to 365 days | latest | nan | False | 0.11 | 8 | 0.00 |
| **1019** | clozapine | 0 to 365 days | boolean | 0 | False | 0.00 | 2 | 1.00 |
| **1020** | clozapine | 0 to 183 days | boolean | 0 | False | 0.00 | 2 | 1.00 |

| | Feature name | Lookbehind period | Resolve multiple | Fallback strategy | Static | Mean | N. unique | Proportion using fallback |
|---|---|---|---|---|---|---|---|---|
| **1021** | clozapine | 0 to 730 days | boolean | 0 | False | 0.00 | 2 | 1.00 |
| **1022** | f0_disorders | 0 to 183 days | boolean | 0 | False | 0.01 | 2 | 0.99 |
| **1023** | f0_disorders | 0 to 365 days | boolean | 0 | False | 0.01 | 2 | 0.99 |
| **1024** | f0_disorders | 0 to 730 days | boolean | 0 | False | 0.02 | 2 | 0.98 |
| **1025** | f1_disorders | 0 to 730 days | boolean | 0 | False | 0.07 | 2 | 0.93 |
| **1026** | f1_disorders | 0 to 183 days | boolean | 0 | False | 0.03 | 2 | 0.97 |
| **1027** | f1_disorders | 0 to 365 days | boolean | 0 | False | 0.05 | 2 | 0.95 |
| **1028** | f2_disorders | 0 to 183 days | boolean | 0 | False | 0.02 | 2 | 0.98 |
| **1029** | f2_disorders | 0 to 365 days | boolean | 0 | False | 0.03 | 2 | 0.97 |
| **1030** | f2_disorders | 0 to 730 days | boolean | 0 | False | 0.03 | 2 | 0.97 |
| **1031** | f3_disorders | 0 to 183 days | boolean | 0 | False | 0.18 | 2 | 0.82 |
| **1032** | f3_disorders | 0 to 730 days | boolean | 0 | False | 0.35 | 2 | 0.65 |
| **1033** | f3_disorders | 0 to 365 days | boolean | 0 | False | 0.27 | 2 | 0.73 |
| **1034** | f4_disorders | 0 to 730 days | boolean | 0 | False | 0.36 | 2 | 0.64 |
| **1035** | f4_disorders | 0 to 365 days | boolean | 0 | False | 0.28 | 2 | 0.72 |
| **1036** | f4_disorders | 0 to 183 days | boolean | 0 | False | 0.18 | 2 | 0.82 |
| **1037** | f5_disorders | 0 to 365 days | boolean | 0 | False | 0.05 | 2 | 0.95 |
| **1038** | f5_disorders | 0 to 183 days | boolean | 0 | False | 0.03 | 2 | 0.97 |

| | Feature name | Lookbehind period | Resolve multiple | Fallback strategy | Static | Mean | N. unique | Proportion using fallback |
|---|---|---|---|---|---|---|---|---|
| **1039** | f5_disorders | 0 to 730 days | boolean | 0 | False | 0.06 | 2 | 0.94 |
| **1040** | f6_disorders | 0 to 730 days | boolean | 0 | False | 0.19 | 2 | 0.81 |
| **1041** | f6_disorders | 0 to 365 days | boolean | 0 | False | 0.15 | 2 | 0.85 |
| **1042** | f6_disorders | 0 to 183 days | boolean | 0 | False | 0.10 | 2 | 0.90 |
| **1043** | f7_disorders | 0 to 365 days | boolean | 0 | False | 0.01 | 2 | 0.99 |
| **1044** | f7_disorders | 0 to 730 days | boolean | 0 | False | 0.01 | 2 | 0.99 |
| **1045** | f7_disorders | 0 to 183 days | boolean | 0 | False | 0.01 | 2 | 0.99 |
| **1046** | f8_disorders | 0 to 730 days | boolean | 0 | False | 0.04 | 2 | 0.96 |
| **1047** | f8_disorders | 0 to 365 days | boolean | 0 | False | 0.03 | 2 | 0.97 |
| **1048** | f8_disorders | 0 to 183 days | boolean | 0 | False | 0.02 | 2 | 0.98 |
| **1049** | f9_disorders | 0 to 365 days | boolean | 0 | False | 0.13 | 2 | 0.87 |
| **1050** | f9_disorders | 0 to 183 days | boolean | 0 | False | 0.08 | 2 | 0.92 |
| **1051** | f9_disorders | 0 to 730 days | boolean | 0 | False | 0.17 | 2 | 0.83 |
| **1052** | first_gen_antipsychotics | 0 to 365 days | boolean | 0 | False | 0.03 | 2 | 0.97 |
| **1053** | first_gen_antipsychotics | 0 to 730 days | boolean | 0 | False | 0.04 | 2 | 0.96 |
| **1054** | first_gen_antipsychotics | 0 to 183 days | boolean | 0 | False | 0.02 | 2 | 0.98 |
| **1055** | hamilton_d17 | 0 to 365 days | latest | nan | False | 17.80 | 44 | 0.00 |
| **1056** | hamilton_d17 | 0 to 183 days | latest | nan | False | 17.87 | 44 | 0.00 |

| | Feature name | Lookbehind period | Resolve multiple | Fallback strategy | Static | Mean | N. unique | Proportion using fallback |
|---|---|---|---|---|---|---|---|---|
| **1057** | hamilton_d17 | 0 to 730 days | latest | nan | False | 17.78 | 44 | 0.00 |
| **1058** | lamotrigine | 0 to 365 days | boolean | 0 | False | 0.02 | 2 | 0.98 |
| **1059** | lamotrigine | 0 to 730 days | boolean | 0 | False | 0.03 | 2 | 0.97 |
| **1060** | lamotrigine | 0 to 183 days | boolean | 0 | False | 0.01 | 2 | 0.99 |
| **1061** | lithium | 0 to 730 days | boolean | 0 | False | 0.01 | 2 | 0.99 |
| **1062** | lithium | 0 to 183 days | boolean | 0 | False | 0.00 | 2 | 1.00 |
| **1063** | lithium | 0 to 365 days | boolean | 0 | False | 0.01 | 2 | 0.99 |
| **1064** | physical_visits_to_psychiatry | 0 to 730 days | count | 0 | False | 31.65 | 444 | 0.01 |
| **1065** | physical_visits_to_psychiatry | 0 to 365 days | count | 0 | False | 23.87 | 275 | 0.02 |
| **1066** | physical_visits_to_psychiatry | 0 to 183 days | count | 0 | False | 15.92 | 150 | 0.04 |
| **1067** | physical_visits_to_somatic | 0 to 730 days | count | 0 | False | 3.79 | 124 | 0.40 |
| **1068** | physical_visits_to_somatic | 0 to 365 days | count | 0 | False | 2.03 | 82 | 0.54 |
| **1069** | physical_visits_to_somatic | 0 to 183 days | count | 0 | False | 1.04 | 63 | 0.67 |
| **1070** | pregabaline | 0 to 730 days | boolean | 0 | False | 0.03 | 2 | 0.97 |
| **1071** | pregabaline | 0 to 183 days | boolean | 0 | False | 0.02 | 2 | 0.98 |
| **1072** | pregabaline | 0 to 365 days | boolean | 0 | False | 0.02 | 2 | 0.98 |
| **1073** | second_gen_antipsychotics | 0 to 183 days | boolean | 0 | False | 0.08 | 2 | 0.92 |
| **1074** | second_gen_antipsychotics | 0 to 730 days | boolean | 0 | False | 0.14 | 2 | 0.86 |

| | Feature name | Lookbehind period | Resolve multiple | Fallback strategy | Static | Mean | N. unique | Proportion using fallback |
|---|---|---|---|---|---|---|---|---|
| **1075** | second_gen_antipsychotics | 0 to 365 days | boolean | 0 | False | 0.11 | 2 | 0.89 |
| **1076** | selected_nassa | 0 to 730 days | boolean | 0 | False | 0.04 | 2 | 0.96 |
| **1077** | selected_nassa | 0 to 183 days | boolean | 0 | False | 0.02 | 2 | 0.98 |
| **1078** | selected_nassa | 0 to 365 days | boolean | 0 | False | 0.03 | 2 | 0.97 |
| **1079** | sex_female | 0 | N/A | nan | True | 0.65 | 2 | 0.00 |
| **1080** | snri | 0 to 183 days | boolean | 0 | False | 0.03 | 2 | 0.97 |
| **1081** | snri | 0 to 730 days | boolean | 0 | False | 0.07 | 2 | 0.93 |
| **1082** | snri | 0 to 365 days | boolean | 0 | False | 0.05 | 2 | 0.95 |
| **1083** | ssri | 0 to 730 days | boolean | 0 | False | 0.10 | 2 | 0.90 |
| **1084** | ssri | 0 to 365 days | boolean | 0 | False | 0.07 | 2 | 0.93 |
| **1085** | ssri | 0 to 183 days | boolean | 0 | False | 0.05 | 2 | 0.95 |
| **1086** | tca | 0 to 183 days | boolean | 0 | False | 0.02 | 2 | 0.98 |
| **1087** | tca | 0 to 365 days | boolean | 0 | False | 0.03 | 2 | 0.97 |
| **1088** | tca | 0 to 730 days | boolean | 0 | False | 0.04 | 2 | 0.96 |
| **1089** | valproate | 0 to 730 days | boolean | 0 | False | 0.01 | 2 | 0.99 |
| **1090** | valproate | 0 to 365 days | boolean | 0 | False | 0.01 | 2 | 0.99 |
| **1091** | valproate | 0 to 183 days | boolean | 0 | False | 0.00 | 2 | 1.00 |
